# Multicentre electrophysiology reveals the limits of auditory prediction responses as biomarkers of chronic tinnitus

**DOI:** 10.64898/2026.11.25.26368835

**Authors:** Nathan Shields, Elva Arulchelvan, Feifan Chen, Anusha Yasoda-Mohan, Payam S. Shabestari, Shagun Ajmera, Milena Engelke, Namitha Jain, Emilia Kaniewska, Tobias Kleinjung, Hongyu Lu, Jan Ost, Lauren Ralston, Sarah Rambour, Susanne Staudinger, Beat Tödtli, Stephan M. Wolpert, Julia Campbell, Christopher R. Cederroth, Dirk De Ridder, Fatima T. Husain, Berthold Langguth, Patrick Neff, Winfried Schlee, Sven Vanneste

## Abstract

Identifying reliable neural biomarkers of chronic tinnitus remains challenging because ageing and hearing loss independently influence auditory cortical activity. Here, we investigated auditory expectation processing in 440 adults with (n = 226) and without (n = 214) chronic tinnitus using electroencephalography during a local-global auditory paradigm across six centres. Although several electrophysiological group differences emerged in unadjusted analyses, most were substantially attenuated after adjustment for demographic and audiological factors. Two clusters remained significant within the unexpected-omission-versus-sound contrast (44–100 ms and 356–476 ms), but neither survived correction across eight contrasts. Direct comparisons between expected and unexpected omissions revealed no significant group differences. Exploratory classification using selected EEG features achieved a cross-validated area under the receiver operating characteristic curve of 0.624, increasing to 0.689 when combined with demographic and audiological predictors. However, this improvement was not reproduced in held-out validation, and EEG features showed limited predictive value for tinnitus burden or loudness. These findings highlight a fundamental distinction between electrophysiological group differences and clinically informative individual-level biomarkers, underscoring the importance of controlling for confounding factors and establishing independent predictive validity in tinnitus research.

## Introduction

Tinnitus affects approximately 10–15% of adults^1–5^. In its chronic subjective form, it involves the persistent perception of sound without a corresponding external source. Despite its prevalence and clinical burden, its neural basis remains only partly understood, and objective physiological characterisation remains challenging^1,6^. Tinnitus becomes more common with age and often co-occurs with hearing loss^2,6–8^. Both ageing and hearing loss can alter cortical responses to sound^2,6–8^. Differences in cortical auditory responses between people with and without tinnitus may therefore reflect, in part, differences in age and hearing sensitivity. It remains unclear which features of the cortical response to sound are associated with tinnitus after accounting for these factors.

Predictive-processing accounts propose that auditory perception depends on how the brain weighs incoming sensory evidence against prior expectations^9–14^. When auditory input becomes less reliable, greater reliance on prior expectations may favour the perception of sound in the absence of an external source^15,16^. If this altered weighting extends to external sounds, the influence of sequence context on the response to an identical auditory event may differ between people with and without tinnitus. The local-global auditory paradigm provides a way to test this possibility by varying which event listeners can expect at the end of a sound sequence^17^. Two comparisons are particularly informative. Comparing an unexpectedly omitted sound with the sound normally presented at that moment captures the combined effects of violating an expectation and withholding acoustic input. Comparing the same omission across contexts in which it is expected or unexpected holds the acoustic input constant and tests how the response to silence depends on what was anticipated.

Previous EEG studies suggest that tinnitus is associated with differences in how listeners process changes in sound^14,18^. Sedley and colleagues found that unexpected increases in the intensity of tinnitus-like sounds elicited larger mismatch responses than unexpected decreases in participants with tinnitus^18^. Age- and hearing-matched controls showed no comparable asymmetry. Mohan and colleagues used an auditory oddball paradigm that included omissions and reported larger P300 responses in participants with tinnitus and little or no hearing loss^14^. Across a broader range of hearing thresholds, two questions remain. Do people with and without tinnitus differ in their cortical responses to an omitted versus a presented sound? Do they also differ when the same omission is expected in one context and unexpected in another? Even when an EEG response differs on average between people with and without tinnitus, individual responses may overlap substantially across the two groups. A separate question is whether the same response relates to how loud or distressing tinnitus is for an individual^19,20^. Previous work illustrates this distinction by showing that auditory physiological differences associated with tinnitus and sound sensitivity do not necessarily track tinnitus-related distress^21^. Here, we investigated auditory expectation processing in 440 adults with chronic subjective tinnitus (n = 226) or without tinnitus (n = 214), using 64-channel electroencephalography during a local-global auditory oddball paradigm across six centres. Our primary aim was to determine whether group differences in responses to auditory deviance and sound omission persisted after accounting for site, age, sex and hearing thresholds. In exploratory analyses, we assessed how well selected EEG features distinguished participants with and without tinnitus, both alone and in combination with age, sex and hearing thresholds. Among participants with tinnitus, we also examined whether these features predicted tinnitus-related distress, subjective loudness and psychophysically matched loudness.

## Methods

### Study design and registration

This study formed part of the Tinnitus Detection (TIDE) project, a prospective, multicentre case-control study conducted between September 2024 and October 2025 at seven centres in Europe and the United States: the University of Texas at Austin, University of Illinois Urbana-Champaign, Trinity College Dublin, Brai3n Clinic in Ghent, University Hospital Zurich, University Clinic Tübingen and University Hospital Regensburg. Trinity College Dublin coordinated the study. The consortium protocol has been published^22^, and the study was registered on ClinicalTrials.gov (NCT06520865).

The present analysis examined EEG responses recorded during the local-global auditory oddball paradigm. Resting-state and gap-in-noise recordings acquired within the broader TIDE project are reported separately. Following the quality-control procedures described below, the analysable oddball cohort included participants from six centres.

The principal objective was to examine differences in auditory deviance- and omission-related responses between participants with and without chronic subjective tinnitus, accounting for site, age, sex and hearing thresholds. Classification of tinnitus status and prediction of continuous clinical outcomes were exploratory analyses not specified in the original protocol. The observational study is reported with reference to STROBE guidelines, and prediction modelling is described with reference to TRIPOD+AI.

### Ethics and sample-size rationale

The study received approval from the relevant ethics committee or institutional review board at each participating centre: University of Texas at Austin (#STUDY00004651), University of Illinois Urbana-Champaign (#IRB24-1117), Trinity College Dublin (#SPREC022020-07), Brai3n Clinic (#6497-B3002024000114), University Hospital Zurich (#KEK-ZH 2025-01839), University Clinic Tübingen (#079/2024BO2) and University Hospital Regensburg (#23-3580-101). All participants provided written informed consent before undergoing study procedures in accordance with the Declaration of Helsinki.

No formal a priori power calculation was performed. The consortium recruitment target was approximately 560 participants (aiming for 40 tinnitus cases and 40 controls per centre), reflecting recruitment capacity within the study period. The final sample was determined by task completion, EEG quality criteria, and complete covariate availability (Figure 1). Predictive modelling was exploratory and evaluated within the available complete-case cohort.

**Figure 1.**
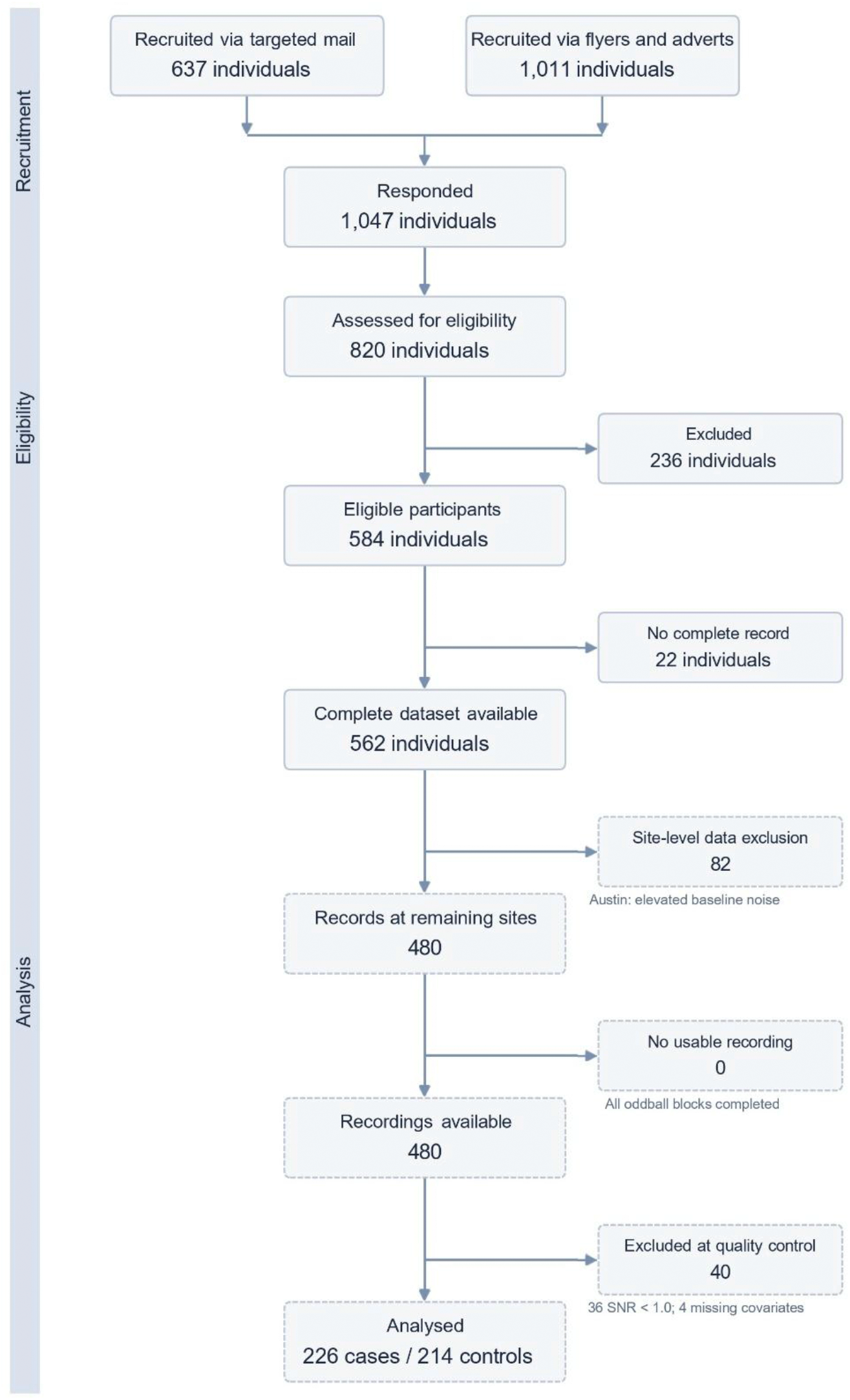
Flow diagram of participant enrolment, screening, quality control, and analysis. Participant progression through the prospective multicentre study. Left vertical bars denote recruitment, eligibility assessment, and analysis stages. Boxes indicate participant counts at each phase, including recruitment via targeted mailing (*n* = 637) and public advertisements (*n* = 1,011), eligibility screening (*n* = 820), exclusions prior to protocol testing (*n* = 236), eligible participants completing baseline procedures (*n* = 584), and records established in the consortium database (*n* = 562; 282 cases, 280 controls). Note that pre-screening response counts were not tracked at UZH, such that responder totals (*n* = 1,047) represent the sum of the other six centres and do not subtract directly into eligibility assessments. Oddball protocol analysis drew directly on participants completing all three task blocks, followed by site-level exclusion of the Austin centre (*n* = 82) due to elevated sensory baseline noise and individual exclusions for sub-threshold signal-to-noise ratio or missing covariates (*n* = 40). The final complete-case analysable cohort comprised 440 individuals (226 cases with chronic subjective tinnitus and 214 healthy controls).

### Participant recruitment and eligibility

Participants were recruited through institutional emails, community flyers and media announcements at participating centres. Recruitment aimed to achieve group-level frequency matching for age, sex and audiometric hearing thresholds. Matching was not fully achieved; demographic and audiological differences were therefore addressed through covariate adjustment, as described in the statistical analysis section. Participant screening, eligibility assessment and subsequent exclusions are summarised in Figure 1.

General eligibility criteria were age 18–70 years, capacity to understand the study procedures and provide informed consent, and fulfilment of the prespecified cognitive-function and sound-tolerance criteria. The instruments and thresholds used to operationalise these criteria are described in the clinical and audiometric assessment section below.

Otological and audiological exclusions comprised objective or pulsatile tinnitus as the primary complaint; otosclerosis; vestibular schwannoma; middle- or inner-ear disorders associated with fluctuating hearing; acute ear, nose or throat infection; Ménière’s disease or endolymphatic hydrops; vestibular migraine; severe hearing impairment; and single-sided deafness.

Additional exclusions comprised epilepsy, central nervous system neoplasms, encephalitis or other major intracranial pathology; a history of major neurodegenerative disease or traumatic brain injury with persistent focal deficits; active drug, medication or alcohol abuse within the preceding 12 weeks; implanted electronic devices, including cardiac pacemakers and cochlear implants; and pregnancy.

### Reference standard

The reference standard for tinnitus status was based on a structured clinical interview. Participants in the tinnitus group were required to report chronic, constant, subjective, non-pulsatile primary tinnitus lasting at least six months. Interviews were conducted by an audiologist at the US sites and by a physician at the European sites. Tinnitus characteristics were documented using the study questionnaire battery, as described in the clinical and audiometric assessment section.

Case and control status was established through standardised self-report during the eligibility interview, before EEG acquisition. Consequently, index-test results were unavailable to the assessors when the reference standard was established. Controls required no experience of tinnitus lasting longer than two minutes on any single occasion. No intervention occurred between the reference-standard assessment and the EEG recording.

### Clinical and audiometric assessment

Participants underwent clinical, questionnaire-based and audiometric assessment at each centre. Questionnaires were administered electronically through REDCap or on paper, except at Zürich, where responses were collected using UniPark and subsequently recoded for integration into the consortium database.

Cognitive function was assessed using the Montreal Cognitive Assessment (MoCA), with a minimum eligibility score of ≥26; assessments were administered only by certified researchers. Tinnitus history and characteristics were assessed using the European School for Interdisciplinary Tinnitus Research Survey Questionnaire (ESIT-SQ)^23^. All participants were intended to complete Part A, whereas Part B was administered to participants with tinnitus only. However, controls from Zürich did not complete the ESIT-SQ (n = 36 missing). Tinnitus-related burden was assessed using the 25-item Tinnitus Handicap Inventory (THI; range 0–100)^24^, which was the principal burden measure used in the present analyses. The Tinnitus Functional Index (TFI) was also collected as part of the broader consortium assessment^25,26^. Both the THI and TFI were administered to participants with tinnitus only. Subjective tinnitus loudness was measured using a visual analogue scale (VAS; range 0–100).

Sound hypersensitivity was assessed using the Hyperacusis Questionnaire (HQ)^27,28^, with a maximum eligibility score of ≤27. At Zürich, a German version of the HQ with three response options per item was administered, whereas the other six centres used a four-option version. Consequently, no participant at Zürich was excluded on the basis of the HQ criterion. Anxiety and depression were assessed using the Hospital Anxiety and Depression Scale (HADS)^29^, which was collected in both groups. A HADS score >14 was initially used as an exclusion criterion at several sites but was modified during recruitment. HADS anxiety and depression subscale scores were included as additional covariates in secondary analyses. Perceived stress was assessed using the 30-item Perceived Stress Questionnaire (PSQ-30)^30^. There were fifteen missing entries on the PSQ score. Further details of the consortium assessment battery are provided in the published protocol^22^. Audiometry was performed in sound-treated environments using calibrated clinical audiometers. Bilateral air-conduction thresholds were measured at 0.5, 1, 2, 3, 4, 6, 8, 10 and 12.5 kHz, with additional testing up to 16 kHz where available. Bone-conduction testing and masking were performed when indicated. Hearing sensitivity was summarised using the pure-tone average across air-conduction thresholds at 0.5, 1, 2 and 4 kHz (PTA4), calculated bilaterally for covariate adjustment and group comparisons. Consistent with the study protocol, participants with severe hearing loss, defined as a PTA4 ≥71 dB HL in the better-hearing ear, were excluded from the study. When a participant did not respond at the maximum available output, the threshold was recorded as that output plus 5 dB. This convention did not affect any threshold contributing to PTA4.

Participants with tinnitus additionally completed repeated tinnitus pitch- and loudness-matching assessments. Matched loudness was recorded in decibels sensation level (dB SL) or hearing level (dB HL), depending on the centre. To harmonise measurements across centres, the hearing threshold (*T*) at the individually matched tinnitus frequency (*f*_tinnitus_) was estimated from the audiogram by linear interpolation in log_2_-transformed frequency space:

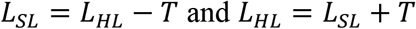

Conversions were performed separately per ear and averaged to derive participant-level mean loudness estimates in dB SL and dB HL, with mean loudness in dB HL serving as the primary measure. THI scores, subjective VAS loudness and psychophysically matched loudness were analysed separately to distinguish tinnitus-related burden, perceived loudness and the level of an external sound matched to the tinnitus percept.

### EEG acquisition and stimulus delivery

Continuous EEG was recorded across seven clinical centres using 64-channel acquisition systems with site-specific cap configurations, recording references, online filters, and sampling rates (Supplementary Table 1). Acoustic stimuli were delivered through insert earphones (EARtone GOLD 3A or Etymotic ER2), with calibration performed using sound-level meters or artificial ear simulators (Supplementary Table 1). Before recording, presentation levels were individually calibrated using a two-stage behavioural procedure. Participants first rated the loudness of four 500-Hz tones presented at varying levels on a nine-point scale ranging from “very soft” to “very uncomfortable”, allowing a comfortable tone presentation level to be established. They then rated a white-noise burst presented after four tones at the selected level, using a nine-point scale ranging from “very soft” to “very loud”. The noise level was adjusted to match the perceived loudness of the tones. Participants maintained visual fixation on a central cross throughout auditory testing. No adverse events related to the recordings were reported. The broader TIDE protocol also included resting-state and gap-in-noise recordings, which are reported separately in companion manuscripts: cGPIAS analyses in 451 participants by Lu et al. and resting-state normative connectivity modelling in 544 participants by Shabestari et al.

### Local-global auditory paradigm and contrasts

The task comprised three protocols presenting five-event sequences: five consecutive 500-Hz pure tones (*xxxxx*), four tones followed by a white-noise burst (*xxxxY*), or four tones followed by omission of the fifth event (*xxxx*_). Stimuli lasted 50 ms, including 7-ms linear onset and offset ramps, with a 300-ms stimulus onset asynchrony (250-ms offset-to-onset interval) and a randomly jittered inter-trial interval of 1,300–1,500 ms (Figure 2A). Site-specific acoustic calibration equipment and PsychoPy versions are listed in Supplementary Table 1.

**Figure 2.**
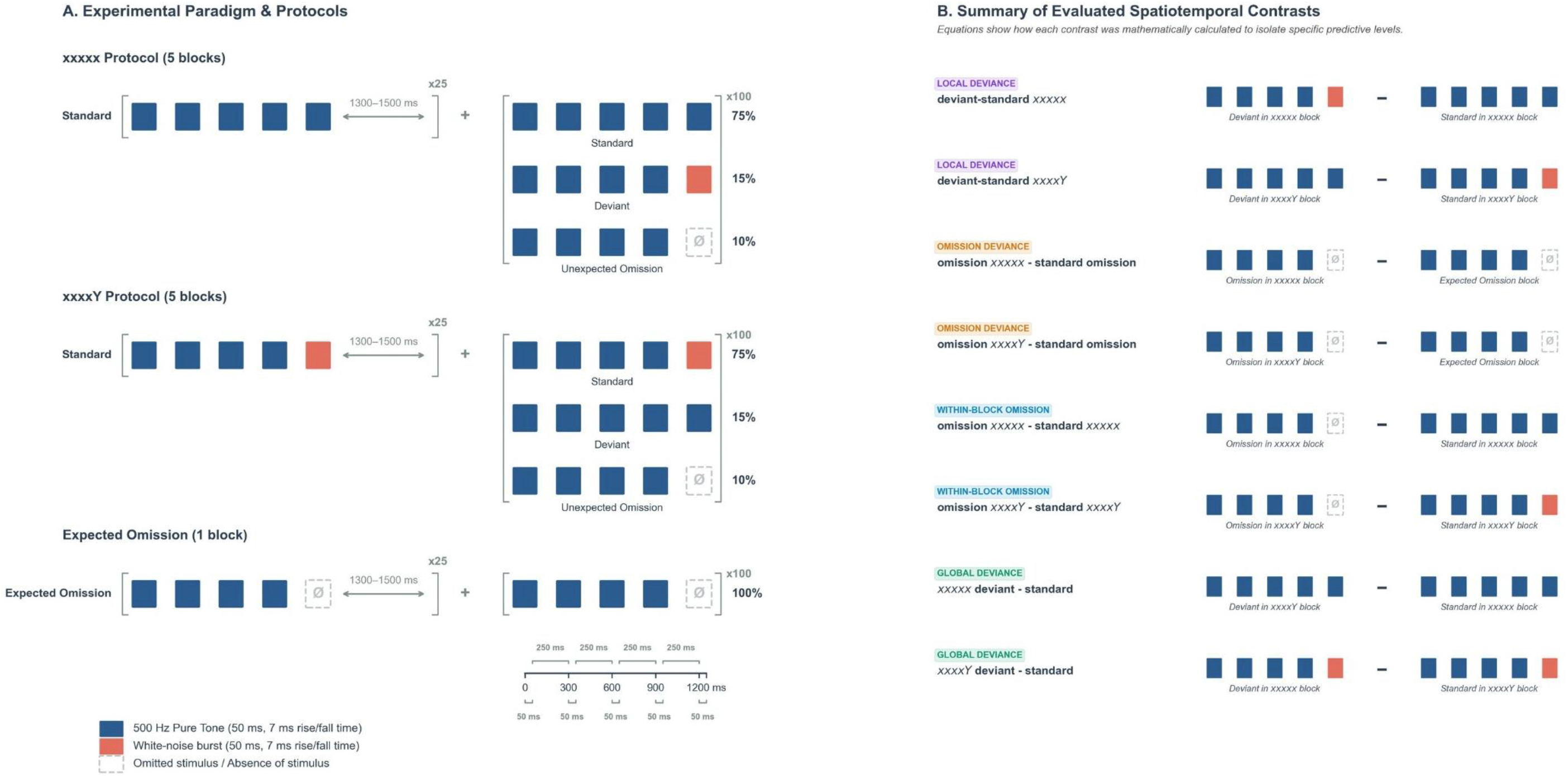
Hierarchical Auditory Local-Global Oddball Paradigm and Evaluated Spatiotemporal Contrasts. **(A) Experimental Paradigm & Protocols.** The experimental framework consists of three distinct protocols designed to separate prediction error processing at different levels of the auditory hierarchy. **Top**: The *xxxxx* Protocol. Each trial is a five-element sequence comprising 500 Hz pure tones (50 ms duration, 7 ms linear rise/fall ramps; represented in blue). In training blocks (left), a global regularity is established via 25 consecutive standard (*xxxxx*) sequences. In subsequent test blocks (right), 100 pseudo-randomised trials are presented containing 75% Standards (*xxxxx*), 15% Deviants (*xxxxY*; where the fifth element is a white-noise burst, represented in orange), and 10% Unexpected Omissions (*xxxx_*; where the expected fifth tone is omitted, represented in dashed grey). **Middle**: The *xxxxY* Protocol. The structure mirrors the *xxxxx* protocol, but utilises the *xxxxY* sequence as the standard (establishing standard white-noise bursts at the fifth position), while *xxxxx* sequences serve as the deviant. **Bottom**: The Expected Omission Protocol. A baseline block consisting of 100% predictable omission trials (*xxxx*_) to establish expected omission responses. Sequences are separated by a randomised, jittered inter-trial interval (1300-1500 ms). **Timeline & Legend**: Successive elements within each five-element sequence are separated by a standard inter-stimulus interval of 250 ms, with each individual pure-tone or white-noise stimulus lasting 50 ms (total sequence duration = 1250 ms). **(B) Summary of Evaluated Spatiotemporal Contrasts.** Mathematical subtraction equations illustrating the eight spatiotemporal contrasts used to isolate distinct hierarchical levels of auditory prediction error. **Local Deviance** (*deviant-standard xxxxx* and *deviant-standard xxxxY*) evaluate immediate stimulus-level prediction violations within blocks; **Omission Deviance** (*omission xxxxx-standard omission* and *omission xxxxY-standard omission*) evaluate pure top-down omission responses relative to expected baseline omissions; **Within-Block Omissions** (*omission xxxxx-standard xxxxx* and *omission xxxxY-standard xxxxY*) evaluate within-block expectation violations; and **Global Deviance** (*xxxxx deviant-standard* and *xxxxY deviant-standard*) evaluate contextual, sequence-level prediction violations using physically identical terminal elements presented in opposing global contexts.

The *xxxxx* and *xxxxY* protocols each comprised five blocks of 125 trials. Each block began with 25 repetitions of the protocol-specific standard, followed by 100 test trials comprising 75 standards, 15 deviants and 10 unexpected omissions. Only the 75 test-phase standards in each block were eligible for inclusion in the standard ERP, subject to artefact rejection. Standard and deviant sequence identities were reversed between the two protocols, whereas omissions were unexpected in both. A third protocol comprised 125 sequences with predictable omission of the fifth event. Protocol and test-trial order were pseudorandomised, yielding 1,375 trials overall. Participants could take self-paced breaks between blocks, with fatigue and comfort monitored between protocols (Figure 2A).

Eight ERP contrasts were evaluated across four families (Figure 2B). Within-block deviance contrasts compared deviant with standard responses separately within each sound-present protocol, reflecting differences in terminal-event identity and sequence probability. Global contextual contrasts compared physically identical sequences across protocols in which they served as deviants or standards, assessing responses associated with global sequence context while holding sequence identity constant. Omission deviance contrasts compared unexpected omissions in each sound-present protocol with predictable omissions in the expected-omission protocol, examining omission-related activity across different expectation and block contexts. Within-block omission contrasts compared unexpected omissions with sound-present standards within each protocol, capturing omission-related differences that also incorporate the response to the presented terminal sound in the standard condition.

All temporal measurements were referenced to sequence onset; where relevant, latencies were additionally expressed relative to the fifth event position at 1,200 ms.

### EEG preprocessing and physical montage alignment

EEG preprocessing was performed using MNE-Python (v1.12.1), AutoReject (v0.4.3) and mne-icalabel (v0.8.1). Recordings acquired above 1,000 Hz were downsampled to 1,000 Hz before all datasets were resampled to 250 Hz and zero-phase band-pass filtered (1–40 Hz); site-specific line-noise frequencies (50 or 60 Hz) were notch-filtered. Electrode locations were mapped to manufacturer-specific international 10–05 coordinates (Supplementary Table 1). Illinois digitised electrode coordinates were co-registered to the standard 10–05 template using an affine 3D transformation based on periauricular and nasion fiducials. Signals were average-referenced, and candidate bad channels were identified using Local Outlier Factor (threshold = 3.0), visually inspected by an analyst blinded to diagnostic group, and reconstructed using spherical-spline interpolation. Continuous data were inspected for gross non-physiological movement and mechanical artefacts, which were manually annotated and excluded.

Physiological artefacts were addressed using Extended Infomax independent component analysis (ICA). Components identified by ICLabel as ocular/saccadic artefacts with probability >0.70 were removed, and the cleaned signals reconstructed. Data were epoched from −0.5 to 1.8 s relative to sequence onset and baseline-corrected using the −0.5 to 0 s pre-sequence interval. Epochs overlapping manually annotated artefacts were rejected. Before epoch-level correction, trigger counts were verified separately for each protocol against nominal trial totals, with a permitted deviation of 10% (approximately 500 standards, 75 deviants and 50 unexpected omissions per five-block protocol, and 125 trials for the expected-omission protocol); recordings outside this tolerance were excluded. Remaining epochs were processed using AutoReject, which estimated participant- and sensor-specific peak-to-peak thresholds to interpolate transient sensor artefacts and reject corrupted epochs.

Subject-level condition ERPs and subtraction contrasts were computed in each site’s native montage. For group-level spatiotemporal permutation testing, datasets were restricted to the physical intersection of 52 scalp electrodes shared across recording caps (Supplementary Table 1); non-native channels were not synthesised or interpolated.

### Signal-to-noise ratio quality control and cohort attrition

Sensory signal-to-noise ratio was quantified from the unconditioned evoked response elicited by sequence onset:

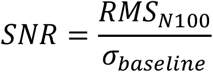

where RMS*_N_*_100_ denotes the root-mean-square amplitude across 90–110 ms post-sequence onset across a nine-channel frontocentral region of interest (FC1, FCz, FC2, C1, Cz, C2, CP1, CPz, CP2) and *σ_baseline_* denotes the standard deviation across the pre-stimulus baseline (−100 to 0 ms). To ensure a consistent minimum level of electrophysiological signal quality across centres, datasets were retained only when SNR was ≥1.0. This threshold was applied before group-level analyses and without reference to tinnitus status. The resulting participant flow and centre-specific exclusions are summarised in Figure 1 and Supplementary Figures S1 and S2.

### Spatiotemporal cluster-based permutation analysis and source localisation

Before group-level testing, sensor-space ERP waveforms from 0.0 to 1.8 s after sequence onset were harmonised across the six retained centres using mean-only NeuroComBat (v0.2.12)^31^. Harmonisation was fitted jointly across cases and controls to maximise the data available for estimating centre effects, while preserving variation associated with age, sex and PTA4. Tinnitus status was not included in the harmonisation model, so diagnostic group membership did not directly inform estimation of the harmonisation parameters^32^. At each electrode and time point, we estimated the tinnitus-control difference using the following ordinary least-squares model, with mean-centred age and PTA4 and dummy-coded sex.

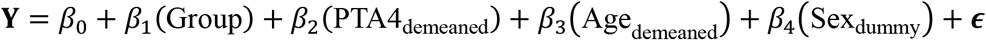

Accordingly, group coefficients represent differences conditional on the included covariates and the harmonisation procedure. The residual degrees of freedom were df_resid_ = *N* − 5 (435 for N = 440; 434 for N = 439). Candidate clusters were formed at two-tailed uncorrected p < 0.05 across ≥2 spatially adjacent channels (Delaunay triangulation). Cluster voltage polarity (positive or negative) is defined relative to the sign of the covariate-adjusted group difference wave (Tinnitus − Control). A positive cluster indicates that event-related potential amplitudes in the tinnitus group are significantly more positive (or less negative) than in the control group within that spatiotemporal window, whereas a negative cluster indicates significantly more negative (or less positive) amplitudes in the tinnitus group.

The eight contrasts were designed to investigate distinct, hierarchically organised aspects of auditory predictive processing within the local-global oddball paradigm, including local deviance detection, omission-related processing and global sequence violations (Figure 2B). Previous ERP studies have demonstrated that auditory responses to omissions depend on the predictability of the omitted stimulus^33^, while tinnitus-related abnormalities have also been associated with altered responses to violations of auditory intensity predictions^18^. These findings support the investigation of distinct predictive processing mechanisms rather than treating all auditory deviance responses as a single neurophysiological phenomenon^17^. Statistical inference was performed separately for each contrast using spatiotemporal cluster-based permutation testing, with 10,000 Freedman-Lane permutations restricted within recording centre^34^. This nonparametric approach addresses the multiple-comparisons problem arising from mass-univariate testing across the 52 common EEG sensors and time points, controlling the family-wise error rate (FWER) within each contrast. Positive and negative clusters were incorporated into the same permutation-based null distribution, with the maximum absolute cluster statistic used to control the FWER across the spatiotemporal search space^35^. The reduced model included age, sex and PTA4, while the full model additionally included tinnitus status.

To assess multiplicity across the eight investigated ERP contrasts, we additionally applied a conservative Bonferroni correction to the contrast-specific cluster-level p-values. This correction supplemented the spatiotemporal permutation procedure, which controlled the family-wise error rate across electrodes, time points and cluster polarities within each contrast. Experiment-wide statistical significance was evaluated at a Bonferroni-adjusted threshold of p < 0.05.

Exploratory cortical source estimates were computed using dynamic statistical parametric mapping (dSPM) on the fsaverage template with a three-layer boundary-element forward model, loose orientation constraint (0.2), depth weighting (0.8) and regularisation *λ*^2^ = 1/9. Noise covariance was estimated using an ad-hoc diagonal noise covariance matrix computed from sensor geometries. Source estimates represented un-thresholded group differences in mean source amplitude averaged over the corresponding empirical sensor-cluster time windows; no source-space statistical inference was performed.

### ERP feature extraction, classification and statistical analysis

Electrophysiological features were extracted across the empirical cluster windows: Integrated ERP Amplitude: Signed trapezoidal numerical integration of the channel-averaged waveform across the cluster ROI:

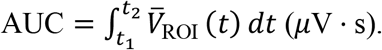

Peak-to-Peak Amplitude: Voltage range across the ROI-averaged waveform:

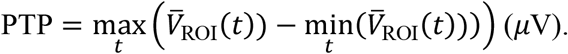

Electrodes and time windows for ERP feature extraction were selected using group-level contrasts from the full cohort before cross-validation. Consequently, participants in the held-out folds contributed to the initial feature-selection process, meaning that standard cross-validation alone cannot eliminate potential circularity. The primary objective of these analyses was therefore exploratory: to assess the incremental predictive information provided by task-evoked ERP features beyond demographic and audiological baselines (age, sex and PTA4).

To assess the potential influence of feature-selection circularity, we performed a stratified random 85:15 discovery-test split, maintaining approximately equal proportions of participants with tinnitus and controls in each subset (Figure 5C–D). NeuroComBat was fitted using the discovery subset, and the resulting parameters were applied to the test subset without refitting. Cluster-permutation testing and ERP feature selection were restricted to the discovery subset; the selected electrodes and time windows were then applied unchanged to the test subset. Models were trained in the discovery subset and evaluated in the held-out test subset. Given the relatively small test sample, this analysis was treated as an internal sensitivity assessment rather than definitive validation. The full-cohort analysis remained primary, with its performance estimates interpreted as exploratory.

Using hierarchical logistic regression, we evaluated EEG features alone and in combination with age, sex and PTA4 in 439 participants with complete data. We compared the age, sex and PTA4 baseline model with the EEG-augmented model using the same participants and identical stratified ten-fold cross-validation splits. Within each fold, we fitted ComBat to the training data and applied the fitted transformation to the held-out participants. We estimated 95% confidence intervals for model performance and for the difference between models using 1,000 bootstrap resamples of the paired out-of-fold predictions (random seed 42).

Probabilistic calibration (Brier score, calibration slope and intercept) and discrimination across sex, age and hearing-threshold subgroups (Sex, Age <50 vs ≥50, PTA4 <15 vs ≥15 dB HL) were evaluated following TRIPOD+AI guidelines (Supplementary Table 6). Multi-centre parameter stability was examined using leave-one-site-out (LOSO) sensitivity analyses, holding out each centre in turn and cross-validating across the remaining centres to assess sensitivity to cohort composition, distinct from external validation on an unseen centre. False discovery rate (FDR) was controlled across candidate predictors within each hierarchical model step using the Benjamini-Hochberg procedure (*p*_FDR_ < 0.05).

### Prediction of Continuous Audiological and Clinical Phenotypes

Hierarchical linear regression and regularised Elastic Net models (nested 5-fold inner tuning, 10-fold outer evaluation) predicted continuous outcomes within the tinnitus cohort. Univariable analyses used all participants with available data for the relevant measure. Multivariable models were restricted to complete cases across the predictors included in each model, yielding n = 217 for PTA4, THI and subjective VAS loudness, and n = 171 for psychophysically matched loudness in dB HL. Listwise deletion was applied within each multivariable model. Models were evaluated against a training-set-mean benchmark using out-of-fold cross-validated *R*^2^, MAE, RMSE, and Pearson’s *r*.

## Results

### Demographic and Clinical Cohort Profile

Across seven centres, 820 individuals were assessed for eligibility, 584 met study criteria, and 562 had complete task datasets (282 with tinnitus and 280 controls; Figure 1; Supplementary Table 2). Quality assessment excluded the Austin cohort (n = 82) because of elevated baseline noise and absent sensory-evoked responses (Supplementary Figures S1–S2). A further 40 participants were excluded across the remaining six centres: 36 for sensory signal-to-noise ratios below 1.0, 2 for missing clinical covariates, and 2 for corrupted task triggers. The final analysis included 440 participants from six centres: 226 with chronic subjective tinnitus and 214 controls. In ERP-only analyses, sample size depended on contrast trial availability (*n* = 440 for the *omission xxxxY-standard xxxxY* contrast; *n* = 439 for the *xxxxx deviant-standard* contrast due to insufficient trial counts for one tinnitus participant), while complete-case covariate-adjusted classification models included 439 participants due to missing biological sex data for one tinnitus participant.

Participants with tinnitus were older than controls (49.38 ± 14.34 versus 44.31 ± 15.31 years; two-tailed Welch’s *t*(431) = 3.59, *p* < .001), had higher bilateral pure-tone average thresholds (14.79 ± 10.88 versus 10.98 ± 8.26 dB HL; two-tailed Welch’s *t*(424) = 4.15, *p* < .001), and included a higher proportion of males (tinnitus: 119 males, 106 females [*n* = 225; 1 missing] vs. controls: 90 males, 124 females [*n* = 214]; *χ*^2^(1) = 4.74, *p* = .030; Figure 3A–C). Clinical symptom distributions are shown in Figure 3D–F. Subsequent analyses accounted for acquisition-site variation, age, sex and hearing thresholds, as appropriate to each analytical model.

**Figure 3.**
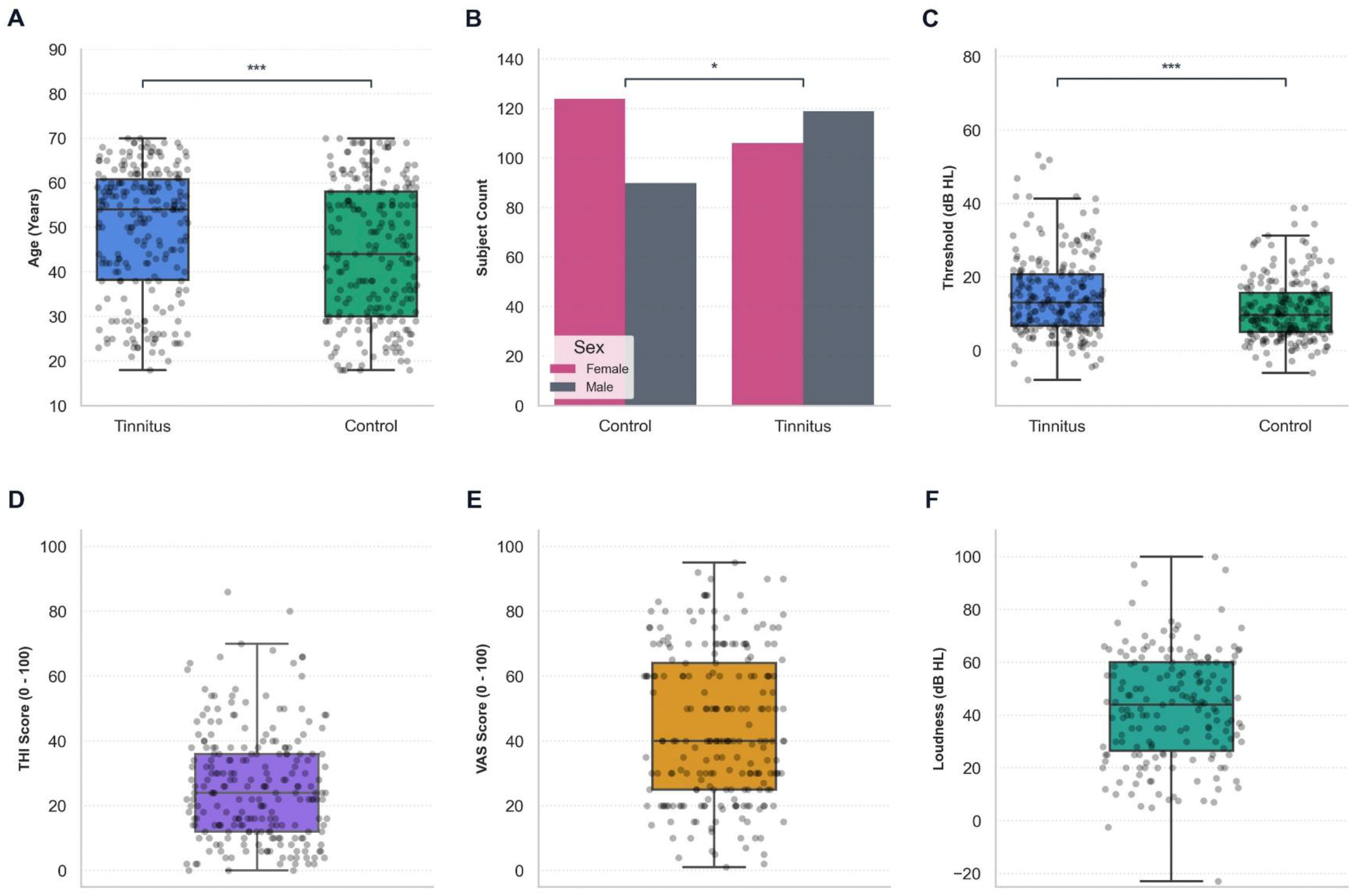
Baseline Cohort Demographics and Clinical Distributions. **(A–C)** Group comparisons between participants with chronic tinnitus (n = 226, blue) and healthy controls (n = 214, green): **(A)** chronological age showing an older tinnitus cohort (two-tailed Welch’s t(431) = 3.59, P < .001); **(B)** biological sex distribution showing a higher proportion of males in the tinnitus group (χ^2^(1) = 4.74, two-sided P = .030; females in magenta, males in grey); and **(C)** bilateral pure-tone average hearing thresholds across 0.5, 1, 2, and 4 kHz (PTA4; two-tailed Welch’s t(424) = 4.15, P < .001). **(D–F)** Clinical symptom distributions within the tinnitus cohort: **(D)** tinnitus distress measured by the Tinnitus Handicap Inventory (THI, purple; n = 226); **(E)** subjective tinnitus loudness on a 0–100 visual analogue scale (VAS, orange; n = 218); and **(F)** psychophysically matched tinnitus loudness (dB HL, teal; n = 179). Boxplot lines indicate median, boxes depict the interquartile range (IQR; 25th–75th percentiles), and whiskers represent 1.5 × IQR; scatter points depict individual participant values. Significance: ∗ P < .05, ***P < .001.

### Demographic and audiological adjustment substantially attenuates electrophysiological group differences

We first examined whether electrophysiological differences between participants with and without chronic tinnitus persisted after accounting for demographic and audiological variation. Unadjusted spatiotemporal analyses identified group differences across several auditory deviance and omission contrasts (Supplementary Table 3 and Supplementary Figure S3). However, these differences were substantially reduced after accounting for site, age, sex and hearing thresholds, indicating that much of the apparent electrophysiological variation was associated with these factors.

Following covariate adjustment and contrast-specific spatiotemporal family-wise error correction, only the comparison between unexpected omissions and sound-present standards in the *xxxxY* protocol yielded significant clusters (Supplementary Table 3). Relative to the expected onset of the fifth event, the tinnitus group showed a more positive omission-minus-standard response at 44–100 ms (peak t = 4.19, cluster p = 0.021) and a more negative response at 356–476 ms (peak t = −3.27, cluster p = 0.044; Figure 4).

**Figure 4.**
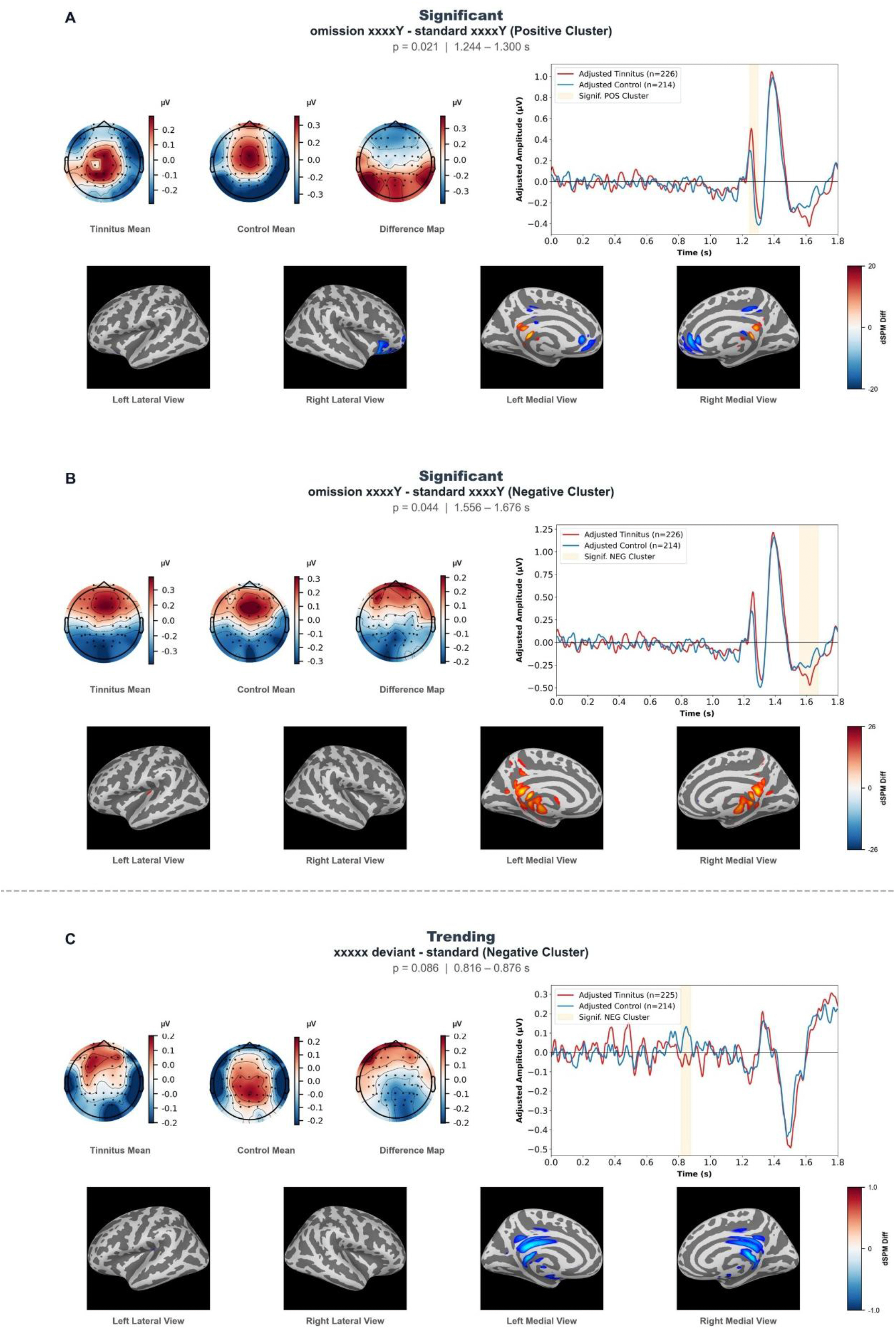
Exploratory spatiotemporal group differences in auditory omission-versus-sound responses. (A) Early positive cluster identified in the unexpected-omission-versus-sound contrast (44–100 ms after the expected terminal event; peak t = 4.19, contrast-specific cluster p = 0.021). (B) Late negative cluster identified in the same contrast (356–476 ms; peak t = −3.27, contrast-specific cluster p = 0.044). Both clusters survived spatiotemporal correction within their respective contrast but did not remain significant after additional correction across all eight ERP contrasts (Bonferroni-adjusted p = 0.168 and 0.352, respectively). Shaded temporal intervals indicate clusters identified by the contrast-specific permutation procedure and should not be interpreted as experiment-wide significant effects. Cortical source reconstructions are descriptive and exploratory. (C) Subthreshold preterminal contextual cluster (contrast-specific p = 0.086), included solely for exploratory feature evaluation.

However, neither cluster remained statistically significant after additional correction across the eight investigated ERP contrasts. Conservative Bonferroni correction yielded adjusted p-values of 0.168 and 0.352 for the early and late clusters, respectively. These findings therefore represent contrast-specific candidate effects rather than experiment-wide significant group differences. Neither comparison between unexpected and expected omissions yielded significant group differences (Figure 4A,B and Supplementary Table 3).

Exploratory template-based dSPM source estimates suggested spatial patterns compatible with activity in posterior cingulate, pregenual anterior cingulate and retrosplenial regions during the early interval (Figure 4A), and retrosplenial and parahippocampal regions during the later interval (Figure 4B). These anatomical estimates were descriptive and were not subjected to independent source-space statistical inference.

A subthreshold cluster was additionally observed in the preterminal global contextual contrast (cluster p = 0.086). Although this cluster did not reach statistical significance, it was retained together with the two omission-related clusters for exploratory evaluation of their potential individual-level predictive value.

### Candidate omission-related differences show limited individual-level discrimination

We next investigated whether the electrophysiological features identified at the group level could distinguish individuals with and without chronic tinnitus. Three candidate ERP features were evaluated using logistic regression with 10-fold cross-validation (n = 439; Figure 5; Supplementary Table 4) , with parallel classification results for the unadjusted (uncontrolled) ERP features reported in Supplementary Figure S4. Peak-to-peak amplitudes provided limited discrimination (combined ROC-AUC = 0.523, 95% CI 0.468–0.580), whereas temporally integrated ERP amplitudes achieved a ROC-AUC of 0.624 (95% CI 0.570–0.673).

**Figure 5.**
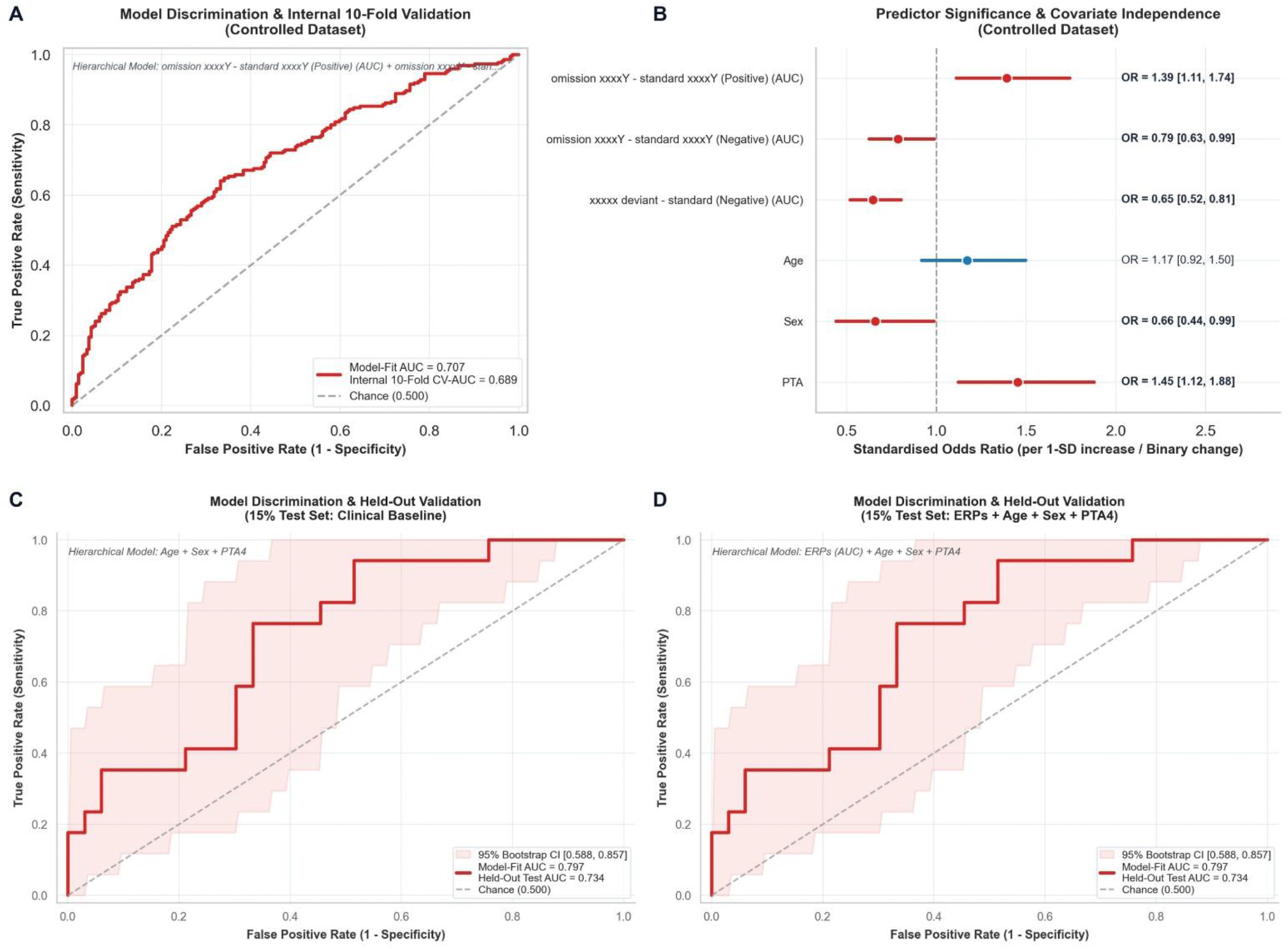
Diagnostic classification of chronic tinnitus in the primary dataset and held-out validation. **(A)** Receiver operating characteristic (ROC) curve for the integrated diagnostic classifier (three ERP AUC features + age, sex, and PTA4), showing a 10-fold cross-validated AUC of 0.689 (95% CI [0.640, 0.738]; model-fit AUC = 0.707) against chance level (dashed grey line). **(B)** Forest plot of standardised odds ratios (OR) with 95% confidence intervals from the integrated diagnostic model; statistically significant predictors (*p* < 0.05) are shown in red and non-significant predictors in blue. **(C)** Held-out validation ROC curve (15% test set) for the clinical baseline classifier (age, sex, and PTA4; model-fit AUC = 0.797, held-out test AUC = 0.734, McFadden’s Δ*R*^2^ = 0.249, *p*(Δ*R*^2^) = .001) with a shaded 95% bootstrap confidence interval (0.588–0.857). **(D)** Held-out validation ROC curve (15% test set) for the combined classifier incorporating ERP features, age, sex and PTA4 (model-fit AUC = 0.797, held-out test AUC = 0.734, McFadden’s Δ*R*^2^ = 0.234, *p*(Δ*R*^2^) = .002) with a shaded 95% bootstrap confidence interval (0.588–0.857).

The model containing age, sex and PTA4 achieved a cross-validated ROC-AUC of 0.608 (95% CI 0.551–0.662), compared with 0.689 (95% CI 0.640–0.738) after adding the three integrated ERP amplitudes. The paired difference in ROC-AUC (combined model minus baseline model) was 0.081 (95% CI 0.046-0.109). The model including the ERP features also showed better fit (Δ McFadden’s pseudo-R² = 0.056, likelihood-ratio test *p* < .001). The preterminal contextual feature (OR = 0.65, 95% CI 0.52–0.81; FDR-adjusted p < 0.001) and early positive omission-versus-sound feature (OR = 1.39, 95% CI 1.11–1.74; FDR-adjusted p = 0.016) remained associated with tinnitus status after mutual and covariate adjustment. The later negative omission-versus-sound feature did not survive FDR correction (p = 0.087). Adding anxiety and depression scores yielded a ROC-AUC of 0.716 (95% CI 0.670–0.763).

Sensitivity analyses excluding each centre in turn yielded cross-validated ROC-AUCs of 0.659–0.707 within the remaining participants (Supplementary Table 5). The preterminal contextual feature remained associated with tinnitus status across all six exclusions, and the early omission-versus-sound feature across five. These analyses assessed sensitivity to cohort composition rather than prediction in an unseen centre. The out-of-fold calibration slope was 0.90 and the intercept was −0.08. Subgroup ROC-AUCs ranged from 0.65 to 0.70 (Supplementary Table 6).

These exploratory analyses indicate modest discrimination using the selected ERP features. Because feature selection preceded cross-validation, the performance estimates describe models conditional on those features; validation incorporating feature selection within the training folds is needed to establish generalisable predictive performance.

In an internal held-out sensitivity analysis (Figure 5C–D and Supplementary Figure S5), spatiotemporal cluster-based permutation testing was restricted to the 85% discovery subset. The early positive (*p* = 0.029) and late negative (*p* = 0.049) omission-versus-sound clusters were identified at the contrast-specific significance threshold, although neither represented an experiment-wide significant finding. Models trained in the discovery subset were subsequently evaluated in the untouched 15% test subset. When evaluated in the untouched 15% held-out test subset, the clinical baseline model (*age + sex + PTA4*) achieved a held-out test ROC-AUC of 0.734 (95% CI 0.588–0.857; model-fit AUC = 0.797; Δ*R*^2^ = 0.249, *p*(Δ*R*^2^) = .001). Incorporating the three ERP AUC features alongside *age, sex and PTA4* yielded the same held-out test ROC-AUC of 0.734 (95% CI 0.588–0.857; model-fit AUC = 0.797; Δ*R*^2^ = 0.234, *p*(Δ*R*^2^) = .002), with no meaningful improvement in classification performance over demographic and audiological covariates alone. By contrast, the ERP-only model showed poor held-out discrimination (ROC-AUC = 0.394, 95% CI 0.223–0.563; model-fit AUC = 0.606; Δ*R*^2^ = 0.067, *p*(Δ*R*^2^) = .231; Supplementary Figure S5). Wide confidence intervals on the 15% test subset reflect limited precision from the smaller test sample.

### Physiological classification features do not reliably predict tinnitus burden or loudness

Finally, we examined whether the candidate ERP features captured individual differences in tinnitus-related burden, subjective loudness and psychophysically matched loudness among participants with chronic tinnitus (Figures 6 and 7; Supplementary Tables 7–14).

**Figure 6.**
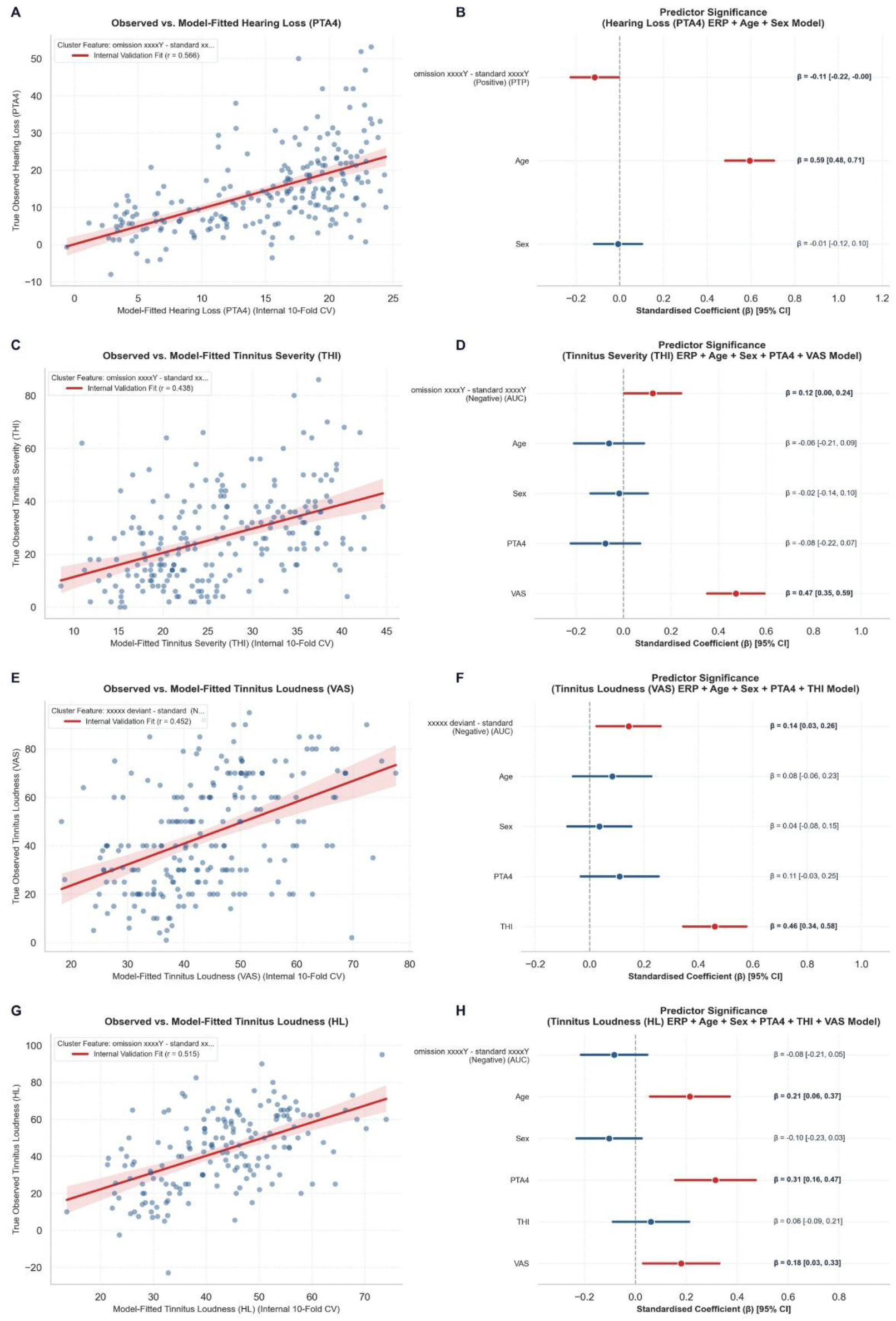
Continuous clinical phenotype predictions in the controlled dataset. Ten-fold cross-validated continuous predictions (left column; scatter points with linear fit and 95% confidence band) and corresponding standardised regression coefficients (*β* with 95% confidence intervals; right column) across clinical targets: **(A, B)** pure-tone average hearing loss (PTA4; internal validation *r* = 0.566); **(C, D)** tinnitus distress measured via the Tinnitus Handicap Inventory (THI; internal validation *r* = 0.438); **(E, F)** subjective tinnitus loudness on a visual analogue scale (VAS; internal validation *r* = 0.452); and **(G, H)** psychophysically matched tinnitus loudness (dB HL; internal validation *r* = 0.515).

**Figure 7.**
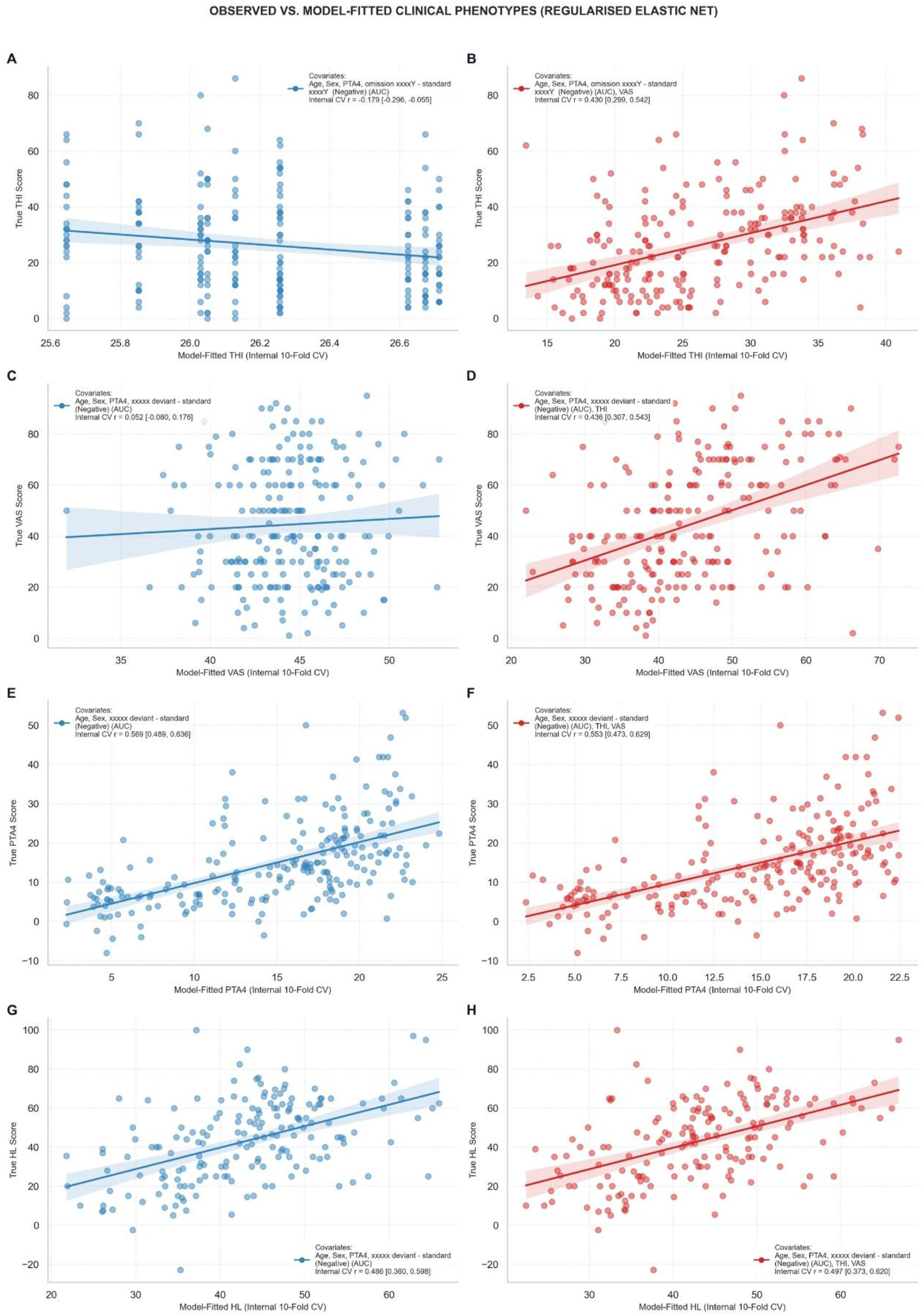
Regularised Elastic Net regression benchmarks across continuous clinical targets. Out-of-sample multivariate predictions from regularised Elastic Net models evaluated using nested cross-validation (inner 5-fold loop for penalty optimisation; outer 10-fold loop for evaluation). Scatter points depict true observed scores versus model-fitted values with regression lines and 95% confidence bands. Left column (blue; A, C, E, G) depicts the objective sensory-demographic model (age, sex, PTA4, and best individual ERP feature). Right column (red; B, D, F, H) depicts the integrated model adding subjective self-report scales. **(A, B)** Tinnitus distress (THI): objective model (internal cross-validated *r* = −.179 [95% CI: −.296, −.055]) versus integrated model adding VAS (*r* = .430 [95% CI: . 299, .542]). **(C, D)** Subjective loudness (VAS): objective model (*r* = .052 [95% CI: −.080, .176]) versus integrated model adding THI (*r* = .436 [95% CI: . 307, .543]). **(E, F)** Hearing loss (PTA4): objective model (*r* = .569 [95% CI: . 489, .636]) versus integrated model (*r* = .553 [95% CI: . 473, .629]). **(G, H)** Psychophysically matched loudness (dB HL): objective model (*r* = .486 [95% CI: . 360, .598]) versus integrated model (*r* = .497 [95% CI: . 373, .620]).

Within the tinnitus cohort, the selected ERP features showed little predictive value for symptom burden or loudness. Models combining all three integrated ERP amplitudes yielded cross-validated correlations of *r* = −0.018 for THI scores and *r* = 0.033 for subjective loudness ratings (*n* = 217). The two clinical measures were more informative about each other. Subjective loudness alone predicted THI scores with *r* = 0.438 and a mean absolute error (MAE) of 11.89 THI points (*n* = 218). A model including subjective loudness, age, sex, hearing thresholds and the late omission-versus-sound ERP feature yielded similar performance (*r* = 0.438, MAE = 11.93 points, *n* = 217). Conversely, THI scores alone predicted subjective loudness with *r* = 0.439 (*n* = 218) (Figure 6 and Supplementary Tables 9–10).

Age was the strongest individual predictor of pure-tone hearing thresholds (PTA4), with a cross-validated correlation of *r* = 0.570 and an MAE of 6.68 dB HL (*n* = 226). A model combining age, sex and the preterminal contextual ERP feature yielded *r* = 0.566 and an MAE of 6.61 dB HL (*n* = 217). ERP features alone also showed little predictive value for psychophysically matched tinnitus loudness, with cross-validated correlations near zero or negative. A model combining the late omission-versus-sound feature with age, sex, PTA4, THI scores and subjective loudness predicted matched loudness with *r* = 0.515 and an MAE of 14.26 dB HL (*n* = 171). The ERP feature showed little independent association with matched loudness in this model (standardised β = −0.084, FDR-adjusted *p* = 0.404). Full model comparisons and site-exclusion analyses are reported in Supplementary Tables 13–14 (Figures 6 and 7).

## Discussion

In this six-centre study of 440 adults, we investigated whether auditory event-related potentials distinguish individuals with and without chronic tinnitus after accounting for demographic and audiological variation. Although several group differences emerged in unadjusted analyses, most were attenuated following harmonisation and adjustment for age, sex and hearing thresholds. Two omission-versus-sound response clusters remained significant after contrast-specific spatiotemporal correction, but neither survived additional correction across the eight investigated contrasts. These findings therefore provide exploratory rather than confirmatory evidence for tinnitus-associated electrophysiological differences. Moreover, although the selected ERP features showed modest discrimination of tinnitus status in cross-validation, their incremental predictive value was not reproduced in the internal held-out test subset, and they provided little information about tinnitus burden or loudness. Together, these results highlight the distinction between candidate electrophysiological group-level associations and neural measures that provide robust, generalisable information at the individual level.

Accounting for demographic and audiological variation narrowed the set of contrasts in which we detected tinnitus-control differences. Age and hearing impairment influence cortical auditory responses and are also associated with tinnitus, making their contributions difficult to separate in case-control comparisons^36–39^. This sensitivity to adjustment may help contextualise heterogeneous findings across studies with different demographic and audiological profiles^14,18,40^. However, attenuation does not establish that age or hearing loss caused the original differences, and persistence after adjustment does not demonstrate tinnitus specificity. PTA4 provides a standardised measure of hearing sensitivity, while additional audiological measures could further clarify the contribution of peripheral auditory function. The adjusted analyses identified candidate electrophysiological differences that warrant further investigation, although none survived correction across all eight ERP contrasts. These exploratory findings provide a basis for testing whether similar response patterns can be reproduced in independent cohorts and distinguished from residual demographic and audiological influences.

In the *xxxxY* protocol, the same four tones preceded either a white-noise burst or its unexpected omission. The observed group difference could therefore reflect the response to the omission, the response to the burst, or both. Comparing expected and unexpected omissions provides a more direct test of auditory expectation because the fifth position is silent in both conditions^17,33,41–43^. Neither of the two expected-versus-unexpected omission contrasts yielded a significant group difference, leaving the contribution of expectation violation to the omission-versus-sound finding unresolved.

The temporal characteristics of the candidate omission-related responses may be compatible with different stages of auditory event processing, although their functional interpretation remains uncertain^17,44^. Exploratory source reconstruction yielded spatial patterns compatible with activity in posterior cingulate, pregenual anterior cingulate, retrosplenial and parahippocampal regions, which have previously been implicated in tinnitus-related neural activity and contextual processing^45–49^. However, these anatomical estimates were derived from template-based source modelling without individual structural MRI or formal source-space statistical inference. Moreover, the underlying sensor-level clusters did not survive correction across all eight ERP contrasts. Consequently, these spatial patterns should be considered descriptive and hypothesis-generating rather than evidence for specific cortical generators or neural mechanisms. Future studies combining electrophysiology with individual anatomical imaging and independent validation will be necessary to establish whether these regions contribute to altered auditory event processing in chronic tinnitus.

The individual-level analyses extended the central question raised in the introduction whether EEG differences associated with tinnitus remain informative when age and hearing sensitivity are considered. Given previous evidence that ageing and hearing loss alter cortical auditory responses^36–38^, evaluating these features alongside demographic and audiological measures is essential for assessing their potential contribution to individual characterisation. Selected EEG features showed modest discrimination of tinnitus status within this cohort, with a neural-only cross-validated ROC-AUC of 0.62, increasing to 0.69 when age, sex and PTA4 were included. Furthermore, including anxiety and depression showed an increase to 0.72. This pattern highlights the relevance of the demographic, audiological context as well as mood in which tinnitus occurs. Adding the three selected ERP features to age, sex and PTA4 increased cross-validated ROC-AUC from 0.608 to 0.689 in this cohort. Temporally integrated ERP features performed better than peak-to-peak measurements, potentially reflecting differences in measurement reliability, noise sensitivity or the representation of sustained activity^50,51^.

These findings should nevertheless be interpreted in light of the exploratory feature-selection procedure. Because the full cohort contributed to initial ERP feature selection, the primary cross-validated performance estimates may be optimistic and do not establish generalisable diagnostic performance. The additional 85:15 held-out sensitivity analysis restricted harmonisation and feature selection to the discovery subset. Although the same contrast-specific significant clusters were identified in this subset, the ERP-only classifier showed poor discrimination in the held-out participants (Supplementary Figure S5). The apparent improvement in discrimination following the addition of ERP features was not reproduced in the held-out test subset, where the demographic and audiological model performed equally well with or without electrophysiological predictors. Although the relatively small test sample limits the precision of this comparison, these findings suggest that the incremental predictive value observed in the primary cross-validation analysis may partly reflect feature-selection optimism. Consequently, the present results do not establish that the selected ERP features provide generalisable diagnostic information beyond readily available demographic and audiological characteristics. Site-exclusion analyses further assessed sensitivity to individual centres. Together, these results motivate further evaluation of EEG for physiological characterisation of tinnitus, rather than establishing a clinically applicable diagnostic classifier.

The clinical prediction analyses clarify which dimensions of tinnitus are captured by the selected EEG features. Previous research has shown that elevated central auditory gain in individuals with tinnitus and sound sensitivity need not track symptom severity, whereas autonomic responses to emotionally evocative sounds can predict tinnitus-related burden^21^. Our findings extend this distinction to task-evoked ERP features: measures selected to distinguish tinnitus from controls contributed little to predicting THI scores or subjective loudness. The contribution is therefore to demonstrate, within the same multicentre cohort, that physiological features associated with tinnitus presence do not necessarily provide useful estimates of its clinical expression. This motivates selecting and validating neural measures specifically for the clinical outcome of interest. THI scores and subjective loudness informed prediction of one another, consistent with their related but distinguishable clinical characteristics^46,52^. Psychophysically matched loudness showed a different predictor pattern, with greater contributions from hearing thresholds and age. Research likewise demonstrated that the relationship between loudness matching and tinnitus severity depends on hearing sensitivity and the measurement scale^53–55^. In the present study, conversion from dB SL to dB HL explicitly incorporates the hearing threshold at the matched frequency, which may partly contribute to the observed audiological association. Together, these findings support treating matched loudness, subjective loudness and tinnitus-related burden as complementary outcomes requiring separate validation of candidate physiological measures.

Future studies can build on these findings by addressing several methodological limitations. Harmonisation enabled pooled analysis, although exclusion of one centre for EEG quality reduced acquisition-system diversity, and generalisation to new recording environments remains to be tested. Although the initial analyses controlled the family-wise error rate across electrodes and time points within each contrast, neither of the two candidate omission-related clusters remained statistically significant after additional correction across all eight investigated contrasts. These findings therefore provide exploratory rather than confirmatory evidence for tinnitus-associated electrophysiological differences. Moreover, because the full cohort contributed to the initial selection of ERP features, the primary cross-validation estimates may be optimistic, as suggested by the absence of incremental predictive value in held-out validation. Although NeuroComBat was used to minimise centre-related variation, future studies should assess the robustness of these findings using alternative approaches to site adjustment, including regression models incorporating recording centre as a fixed effect without prior harmonisation. Source localisation was exploratory and relied on template-based anatomical modelling without individual MRI or formal source-space statistical inference, limiting the anatomical specificity of the observed patterns. More broadly, these methodological considerations illustrate a central challenge in biomarker development: statistical associations identified at the group level may be sensitive to confounding, analytical choices and sample characteristics, limiting their translation into reliable individual-level predictions.

This large multicentre study highlights the challenges of translating electrophysiological group-level associations into reliable individual-level biomarkers of chronic tinnitus. Although candidate differences in auditory event-related potentials emerged following adjustment for demographic and audiological factors, these effects did not survive correction across all investigated contrasts. Moreover, the apparent improvement in tinnitus classification following the addition of EEG features was not reproduced in held-out validation, and these features provided little predictive information regarding tinnitus burden or loudness. These findings illustrate a broader methodological challenge in neuroscience: the identification of neural differences between clinical and control populations does not necessarily establish their diagnostic relevance or generalisability to individual patients. Similar challenges arise in biomarker research across neurological and psychiatric conditions, where group-level neural associations may not translate into robust clinical prediction. Rather than excluding the potential value of electrophysiological biomarkers, our findings emphasise the importance of rigorous confounder adjustment, appropriate control of multiple comparisons, independent validation and outcome-specific biomarker development. Such approaches are essential for moving from descriptive neural associations towards clinically informative measures of brain function.

**Table 1.** Exploratory Classification Performance and Internal Cross-Validation Estimates.

| Model Specification | Predictors Included | In-Sample Fit | Cross-Validated Performance [95% CI] | Performance Gap (Shrinkage) | Metric |
|---|---|---|---|---|---|
| **Diagnostic Prediction Models (Tinnitus vs. Control)** |  |  |  |  |  |
| Clinical Baseline | Age + Sex + PTA4 | .627 | .608 [.551, .662] | .019 | AUC |
| ERP Only (Best) | Best Single ERP Feature | .636 | .624 [.570, .673] | .012 | AUC |
| Integrated Model (Best) | Best Combined ERPs + Age + Sex + PTA4 | .707 | .689 [.640, .738] | .017 | AUC |
| Fully Adjusted Model (Best) | Best ERPs + Age + Sex + PTA4 + HADS | .732 | .716 [.670, .763] | .016 | AUC |
| **Continuous Clinical Phenotype Prediction Models (Tinnitus Cohort)** |  |  |  |  |  |
| Hearing Loss (PTA4) | Age + Sex + PTA4 + Best ERP + Clinical Scales | *r* = .597 (*R*<sup>2</sup> = .356) | *r* = .570 | .027 | Pearson r |
| Tinnitus Distress (THI) | Age + Sex + PTA4 + VAS + Best ERP | *r* = .482 (*R*<sup>2</sup> = .232) | *r* = .438 | .044 | Pearson r |
| Subjective Loudness (VAS) | Age + Sex + PTA4 + THI + Best ERP | *r* = .513 (*R*<sup>2</sup> = .263) | *r* = .452 | .061 | Pearson r |
| Tinnitus Loudness (HL) | Age + Sex + PTA4 + THI + VAS + Best ERP | *r* = .565 (*R*<sup>2</sup> = .319) | *r* = .515 | .050 | Pearson r |

*Note. Performance estimates are based on internal cross-validation conditional on ERP features selected using the full cohort. Consequently, they may be optimistic and do not constitute independent validation of predictive performance.*

## Patient and Public Involvement

No patient or public involvement was incorporated in the design, conduct, reporting, or dissemination of this study.

## Data availability

Anonymised individual participant data (questionnaire, audiometry, EEG) may be made available on reasonable request to the corresponding author, subject to institutional approval and a data sharing agreement. A consortium-wide release is planned once the combined machine-learning analysis of all TIDE paradigms is complete; no date is yet fixed.

## Code availability

The code used for EEG preprocessing, harmonisation, spatiotemporal permutation testing, ERP feature extraction, statistical modelling and figure generation is available through the TIDE Consortium GitHub repository (https://github.com/TIDEConsortium/).

## Author Contributions

**Conceptualisation:** F.T.H., J.C., D.D.R., B.L., W.S., S.V., P.K.A.N., C.R.C., A.Y.M., T.K., and F.C. **Data Curation:** H.L., P.S.S., and N.S. **Formal Analysis:** H.L., P.S.S., and N.S. developed the analysis code and executed the statistical modelling. **Funding Acquisition:** F.T.H., J.C., D.D.R., B.L., W.S., C.R.C., and S.V. **Investigation:** H.L., M.E., S.A., E.A., E.K., F.C., N.J., J.O., S.R., L.R.R., N.S., S.M.W., and A.Y.-M. coordinated site data collection, clinical evaluations, and audiological testing. **Methodology:** N.S., A.Y.M, F.C., H.L., P.S.S., P.K.A.N., S.V. and C.R.C. **Project Administration:** S.S., S.M.W., F.T.H., J.C., D.D.R., B.L., W.S., S.V., and C.R.C. **Resources:** H.L., S.M.W., F.T.H., J.C., D.D.R., B.L., S.V., P.K.A.N., and C.R.C. **Software:** H.L., F.C., N.S., P.S.S., and P.K.A.N. **Supervision:** S.M.W., F.T.H., J.C., D.D.R., B.L., S.V., P.K.A.N., and C.R.C. **Validation:** A.Y.M., E.A., H.L., M.E., and S.S. **Visualisation:** N.S., A.Y.M, H.L., P.S.S., P.K.A.N., S.V., and C.R.C. **Writing - Original Draft:** N.S. and S.V. **Writing - Review & Editing:** All authors critically reviewed, edited, and approved the final manuscript.

## Acknowledgements

We thank all the participants who joined the study. We also thank Fabian Broecker, Chloe NicBhriartaigh, and Caroline Welch for their support in the recruitment procedures at TCD. We also thank Marlies Knipper, Lukas Rüttiger, Stefan Fink and Rodrigo Andrès Donoso-San Martin for their support in the recruitment procedures at UKT, as well as Aurore Brugeaud and Karina Gültig for their administrative support. This project was supported by the Rainwater Charitable Foundation (#18340) and the American Tinnitus Association. C.R.C. received additional funding from the Swedish Research Council (Vetenskapsrådet #2023-02326). P.S.S was also supported by the Swiss National Science Foundation (SNF Project Grant number 208164).

## Supplementary Tables

Table 1. Equipment and Device Summary Across All 7 TIDE Sites.

Table 2. Participant Inclusion and Exclusions by Centre

Table 3. Spatiotemporal Cluster Statistics by Uncontrolled and Controlled Covariates.

Table 4. Logistic Regression Model Results Characterising Diagnostic Prediction - Controlled Dataset.

Table 5. Sensitivity Analysis: Model Robustness Across Leave-One-Site-Out Folds - Controlled Dataset.

Table 6. Calibration and subgroup discrimination of the integrated tinnitus classifier

Table 7. Linear Regression Results Characterising Clinical Prediction - THI Score Models.

Table 8. Sensitivity Analysis Table - Tinnitus Severity (THI) - Linear Regression Model Robustness Across Leave-One-Site-Out Folds.

Table 9. Linear Regression Results Characterising Clinical Prediction - VAS Models.

Table 10. Sensitivity Analysis Table - Subjective Loudness (VAS) - Linear Regression Model Robustness Across Leave-One-Site-Out Folds.

Table 11. Linear Regression Results Characterising Clinical Prediction - PTA4 Models.

Table 12. Sensitivity Analysis Table - Hearing Thresholds (PTA4)- Linear Regression Model Robustness Across Leave-One-Site-Out Folds.

Table 13. Linear Regression Results Characterising Clinical Prediction - Loudness (HL) Models.

Table 14. Sensitivity Analysis Table - Tinnitus Loudness (dB HL) - Linear Regression Model Robustness Across Leave-One-Site-Out Folds.

## References

1 Vanneste, S. et al. Tinnitus. Nat Rev Dis Primers 12 (2026). 10.1038/s41572-026-00702-0

2 Biswas, R. et al. Tinnitus prevalence in Europe: a multi-country cross-sectional population study. Lancet Reg Health Eur 12, 100250 (2022). 10.1016/j.lanepe.2021.100250

3 McCormack, A., Edmondson-Jones, M., Somerset, S. & Hall, D. A systematic review of the reporting of tinnitus prevalence and severity. Hear Res 337, 70–79 (2016). 10.1016/j.heares.2016.05.009

4 Baguley, D., McFerran, D. & Hall, D. Tinnitus. Lancet 382, 1600–1607 (2013). 10.1016/S0140-6736(13)60142-7

5 Langguth, B., Kreuzer, P. M., Kleinjung, T. & De Ridder, D. Tinnitus: causes and clinical management. Lancet Neurol 12, 920–930 (2013). 10.1016/S1474-4422(13)70160-1

6 Shields, N., Arulchelvan, E., Broecker, F. & Vanneste, S. Biomarkers of Subjective Tinnitus Presence and Clinical Burden: A Systematic Review. Hearing Research, 109770 (2026). 10.1016/j.heares.2026.109770

7. Cederroth, C. R., et al. Editorial: Towards an Understanding of Tinnitus Heterogeneity. Front Aging Neurosci 11, 53 (2019). 10.3389/fnagi.2019.00053

8 Biswas, R. & Hall, D. A. Prevalence, Incidence, and Risk Factors for Tinnitus. Curr Top Behav Neurosci 51, 3–28 (2021). 10.1007/7854_2020_154

9 De Ridder, D. & Vanneste, S. The Bayesian brain in imbalance: Medial, lateral and descending pathways in tinnitus and pain: A perspective. Prog Brain Res 262, 309–334 (2021). 10.1016/bs.pbr.2020.07.012

10 De Ridder, D., Vanneste, S. & Freeman, W. The Bayesian brain: phantom percepts resolve sensory uncertainty. Neurosci Biobehav Rev 44, 4–15 (2014). 10.1016/j.neubiorev.2012.04.001

11 Hullfish, J., Sedley, W. & Vanneste, S. Prediction and perception: Insights for (and from) tinnitus. Neurosci Biobehav Rev 102, 1–12 (2019). 10.1016/j.neubiorev.2019.04.008

12 Chen, F., Yasoda-Mohan, A., Se, C. O. & Vanneste, S. Auditory illusory models as proxies to investigate bottom-up and top-down neural networks of phantom perception. Imaging Neurosci (Camb) 3 (2025). 10.1162/imag_a_00574

13 Chen, F., Yasoda-Mohan, A., Se, C. O. & Vanneste, S. Empirically Integrating the Evidence for Different Predictive Coding Components Using Auditory False Perception. Hum Brain Mapp 46, e70211 (2025). 10.1002/hbm.70211

14 Mohan, A., Luckey, A., Weisz, N. & Vanneste, S. Predisposition to domain-wide maladaptive changes in predictive coding in auditory phantom perception. Neuroimage 248, 118813 (2022). 10.1016/j.neuroimage.2021.118813

15 Sedley, W., Friston, K. J., Gander, P. E., Kumar, S. & Griffiths, T. D. An Integrative Tinnitus Model Based on Sensory Precision. Trends Neurosci 39, 799–812 (2016). 10.1016/j.tins.2016.10.004

16 Castejon, J., Chen, F., Yasoda-Mohan, A., Ó Sé, C. & Vanneste, S. Chronic pain - A maladaptive compensation to unbalanced hierarchical predictive processing. Neuroimage 297, 120711 (2024). 10.1016/j.neuroimage.2024.120711

17 Wacongne, C. et al. Evidence for a hierarchy of predictions and prediction errors in human cortex. Proc Natl Acad Sci U S A 108, 20754–20759 (2011). 10.1073/pnas.1117807108

18 Sedley, W., Alter, K., Gander, P. E., Berger, J. & Griffiths, T. D. Exposing Pathological Sensory Predictions in Tinnitus Using Auditory Intensity Deviant Evoked Responses. J Neurosci 39, 10096–10103 (2019). 10.1523/JNEUROSCI.1308-19.2019

19 De Ridder, D. et al. Tinnitus and tinnitus disorder: Genetic, neurobiological, and clinical differentiation. iScience 29, 116080 (2026). 10.1016/j.isci.2026.116080

20 De Ridder, D. et al. Tinnitus and tinnitus disorder: Theoretical and operational definitions (an international multidisciplinary proposal). Prog Brain Res 260, 1–25 (2021). 10.1016/bs.pbr.2020.12.002

21 Smith, S. S., Jahn, K. N., Sugai, J. A., Hancock, K. E. & Polley, D. B. Objective autonomic signatures of tinnitus and sound sensitivity disorders. Sci Transl Med 17, eadp1934 (2025). 10.1126/scitranslmed.adp1934

22 Vanneste, S. et al. Objective data-driven personalised approach to diagnosis of chronic tinnitus: the Tinnitus Detection (TIDE) project - protocol for the identification and validation of a biomarker for tinnitus. BMJ Open 16, e112788 (2026). 10.1136/bmjopen-2025-112788

23 Genitsaridi, E. et al. Standardised profiling for tinnitus research: The European School for Interdisciplinary Tinnitus Research Screening Questionnaire (ESIT-SQ). Hear Res 377, 353–359 (2019). 10.1016/j.heares.2019.02.017

24 Newman, C. W., Jacobson, G. P. & Spitzer, J. B. Development of the Tinnitus Handicap Inventory. Arch Otolaryngol Head Neck Surg 122, 143–148 (1996). 10.1001/archotol.1996.01890140029007

25 Henry, J. A. et al. Tinnitus Functional Index: Development, validation, outcomes research, and clinical application. Hear Res 334, 58–64 (2016). 10.1016/j.heares.2015.06.004

26 Meikle, M. B. et al. The Tinnitus Functional Index: Development of a New Clinical Measure for Chronic, Intrusive Tinnitus. Ear and Hearing 33, 153–176 (2012). 10.1097/AUD.0b013e31822f67c0

27 Khalfa, S. Psychometric normalization of a hyperacusis questionnaire. ORL J Otorhinolaryngol Relat Spec 64, 436–442 (2002). 10.1159/000067570

28 Fackrell, K., Fearnley, C., Hoare, D. J. & Sereda, M. Hyperacusis Questionnaire as a Tool for Measuring Hypersensitivity to Sound in a Tinnitus Research Population. Biomed Res Int 2015, 290425 (2015). 10.1155/2015/290425

29 Zigmond, A. S. & Snaith, R. P. The hospital anxiety and depression scale. Acta Psychiatr Scand 67, 361–370 (1983). 10.1111/j.1600-0447.1983.tb09716.x

30 Levenstein, S. Development of the Perceived Stress Questionnaire: a new tool for psychosomatic research. J Psychosom Res 37, 19–32 (1993). 10.1016/0022-3999(93)90120-5

31 Fortin, J. P. et al. Harmonization of cortical thickness measurements across scanners and sites. Neuroimage 167, 104–120 (2018). 10.1016/j.neuroimage.2017.11.024

32 Johnson, W. E., Li, C. & Rabinovic, A. Adjusting batch effects in microarray expression data using empirical Bayes methods. Biostatistics 8, 118–127 (2007). 10.1093/biostatistics/kxj037

33 Bendixen, A., Schröger, E. & Winkler, I. I heard that coming: event-related potential evidence for stimulus-driven prediction in the auditory system. J Neurosci 29, 8447–8451 (2009). 10.1523/jneurosci.1493-09.2009

34 Winkler, A. M., Ridgway, G. R., Webster, M. A., Smith, S. M. & Nichols, T. E. Permutation inference for the general linear model. Neuroimage 92, 381–397 (2014). 10.1016/j.neuroimage.2014.01.060

35 Maris, E. & Oostenveld, R. Nonparametric statistical testing of EEG- and MEG-data. J Neurosci Methods 164, 177–190 (2007). 10.1016/j.jneumeth.2007.03.024

36 Harris, K. C., Mills, J. H., He, N. J. & Dubno, J. R. Age-related differences in sensitivity to small changes in frequency assessed with cortical evoked potentials. Hear Res 243, 47–56 (2008). 10.1016/j.heares.2008.05.005

37 Oates, P. A., Kurtzberg, D. & Stapells, D. R. Effects of sensorineural hearing loss on cortical event-related potential and behavioral measures of speech-sound processing. Ear Hear 23, 399–415 (2002). 10.1097/00003446-200210000-00002

38 Tremblay, K. L., Piskosz, M. & Souza, P. Effects of age and age-related hearing loss on the neural representation of speech cues. Clin Neurophysiol 114, 1332–1343 (2003). 10.1016/s1388-2457(03)00114-7

39 Gurkan, S., Basokcu, O., Durankaya, S. M., Isler, Y. & Kirkim, G. Central Auditory Changes Associated with Age-related Hearing Loss. Clin EEG Neurosci 55, 508–517 (2024). 10.1177/15500594241243116

40 Riha, C., Guntensperger, D., Kleinjung, T. & Meyer, M. Accounting for Heterogeneity: Mixed-Effects Models in Resting-State EEG Data in a Sample of Tinnitus Sufferers. Brain Topogr 33, 413–424 (2020). 10.1007/s10548-020-00772-7

41 Cho, H. et al. Unexpected sound omissions are signaled in human posterior superior temporal gyrus: an intracranial study. Cereb Cortex 33, 8837–8848 (2023). 10.1093/cercor/bhad155

42 Hughes, H. C. et al. Responses of human auditory association cortex to the omission of an expected acoustic event. Neuroimage 13, 1073–1089 (2001). 10.1006/nimg.2001.0766

43 Braga, A. & Schonwiesner, M. Neural Substrates and Models of Omission Responses and Predictive Processes. Front Neural Circuits 16, 799581 (2022). 10.3389/fncir.2022.799581

44 Jiang, Y. et al. Constructing the hierarchy of predictive auditory sequences in the marmoset brain. Elife 11 (2022). 10.7554/eLife.74653

45 Vanneste, S., Alsalman, O. & De Ridder, D. Top-down and Bottom-up Regulated Auditory Phantom Perception. J Neurosci 39, 364–378 (2019). 10.1523/JNEUROSCI.0966-18.2018

46 Vanneste, S. et al. Switching tinnitus on or off: An initial investigation into the role of the pregenual and rostral to dorsal anterior cingulate cortices. Neuroimage 297, 120713 (2024). 10.1016/j.neuroimage.2024.120713

47 Chen, F., Fahimi Hnazaee, M., Vanneste, S. & Yasoda-Mohan, A. Effective Connectivity Network of Aberrant Prediction Error Processing in Auditory Phantom Perception. Brain Connect 14, 430–444 (2024). 10.1089/brain.2024.0013

48 Bar, M. & Aminoff, E. Cortical analysis of visual context. Neuron 38, 347–358 (2003). 10.1016/s0896-6273(03)00167-3

49 Warrington, O., Graedel, N. N., Callaghan, M. F. & Kok, P. Communication of perceptual predictions from the hippocampus to the deep layers of the parahippocampal cortex. Sci Adv 11, eads4970 (2025). 10.1126/sciadv.ads4970

50 Picton, T. W. et al. Guidelines for using human event-related potentials to study cognition: recording standards and publication criteria. Psychophysiology 37, 127–152 (2000).

51 Clayson, P. E., Baldwin, S. A. & Larson, M. J. How does noise affect amplitude and latency measurement of event-related potentials (ERPs)? A methodological critique and simulation study. Psychophysiology 50, 174–186 (2013). 10.1111/psyp.12001

52 Wallhausser-Franke, E. et al. Tinnitus: distinguishing between subjectively perceived loudness and tinnitus-related distress. PLoS One 7, e34583 (2012). 10.1371/journal.pone.0034583

53 Yakunina, N. & Nam, E. C. What Makes Tinnitus Loud? Otol Neurotol 42, 235–241 (2021). 10.1097/MAO.0000000000002932

54 Andersson, G. Tinnitus loudness matchings in relation to annoyance and grading of severity. Auris Nasus Larynx 30, 129–133 (2003). 10.1016/s0385-8146(03)00008-7

55 Vanneste, S. & De Ridder, D. Stress-Related Functional Connectivity Changes Between Auditory Cortex and Cingulate in Tinnitus. Brain Connect 5, 371–383 (2015). 10.1089/brain.2014.0255

