## Supplementary Figures for "Multicentre electrophysiology reveals the limits of auditory prediction responses as biomarkers of chronic tinnitus"

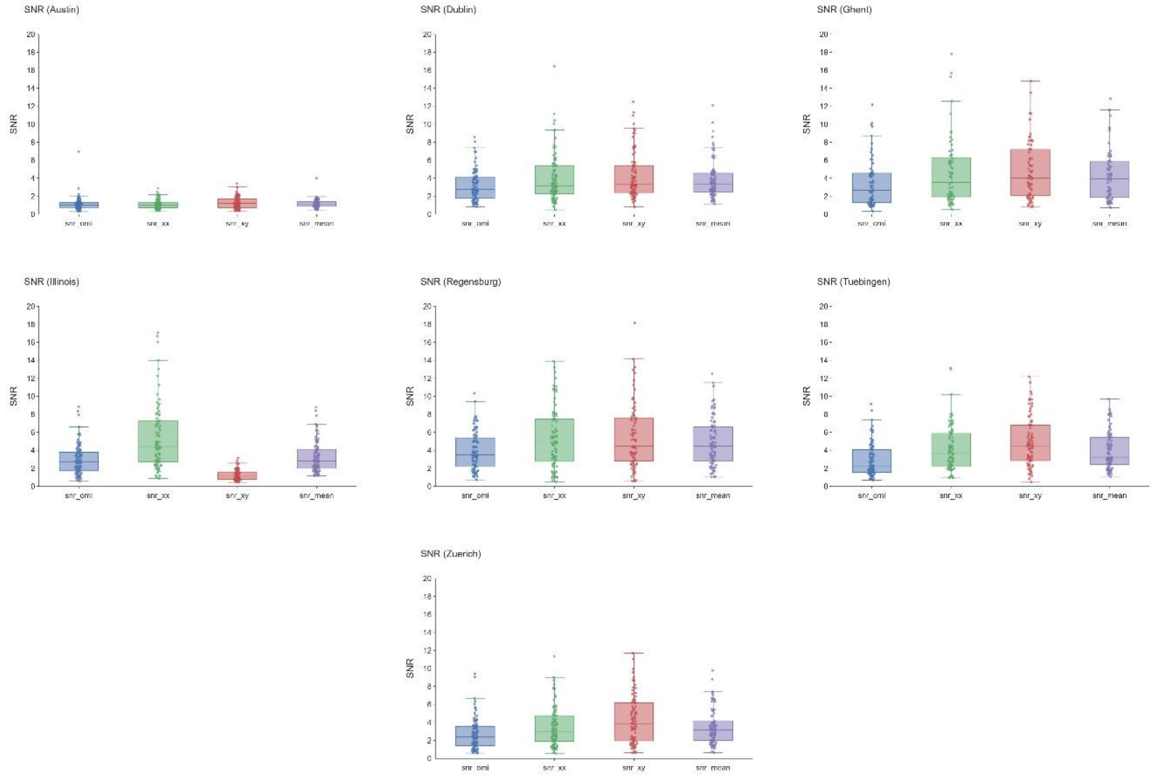


**Supplementary Figure S1. Multi-site signal-to-noise ratio distributions across clinical acquisition centres.**

Participant-level sensory signal-to-noise ratio (SNR) distributions across all seven clinical data collection sites. For each participant, SNR was calculated from the root-mean-square amplitude of the early auditory sensory evoked response (90–110 ms post-stimulus onset across a nine-channel frontocentral region of interest: FC1, FCz, FC2, C1, Cz, C2, CP1, CPz, and CP2) relative to the standard deviation of the pre-stimulus baseline interval ($-100$ to 0 ms). Subplots depict individual site cohorts: Austin, Dublin, Ghent, Illinois, Regensburg, Tübingen, and Zürich. Boxplot center lines represent the median, box edges depict the interquartile range (IQR; 25th–75th percentiles), and whiskers extend to 1.5 × IQR; overlaid scatter points show individual participant SNR values. Horizontal reference lines indicate the pre-specified objective quality retention threshold (SNR ≥ 1.0). High baseline impedance noise systematically depressed SNR across the Austin cohort ($n = 82$), leading to site-level exclusion, whereas individual recordings falling below threshold were excluded across the remaining six sites ($n = 36$).

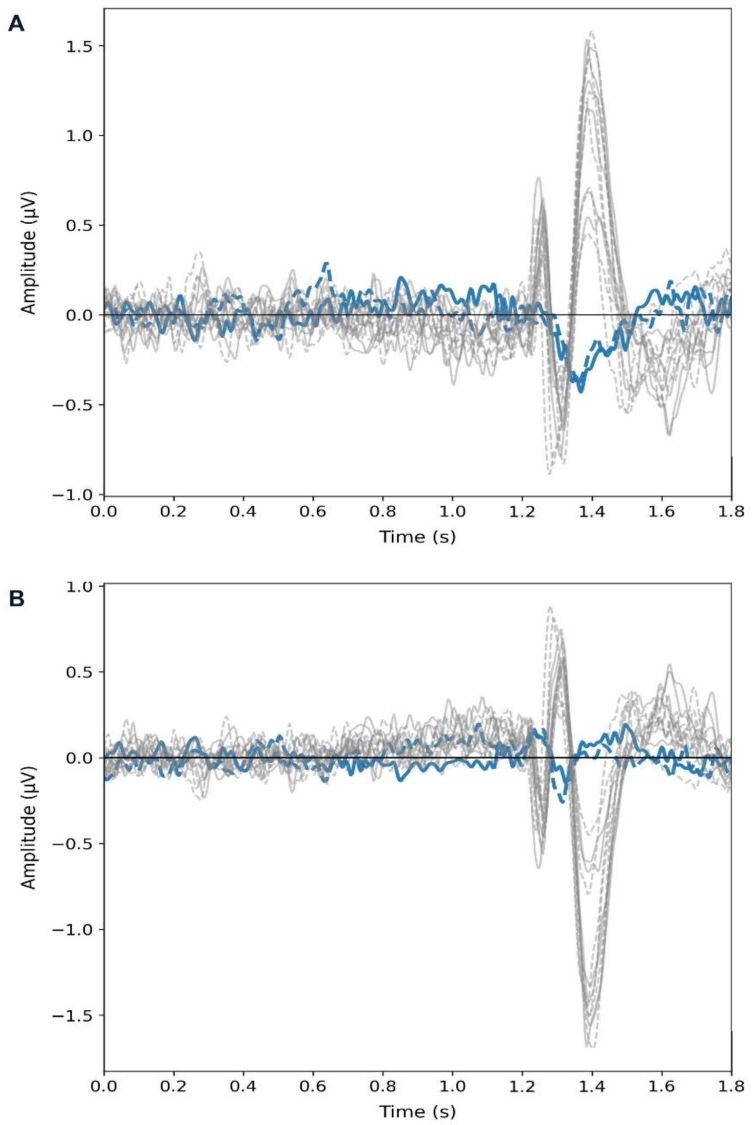


**Supplementary Figure S2. Multi-site grand average event-related potential overlays across clinical centres.**

Grand average event-related potential (ERP) waveforms across all seven data collection centres plotted by diagnostic group for the *omission xxxxY-standard xxxxY* contrast. Traces depict mean voltage trajectories (μV) across the complete epoch (0.0 to 1.8 s) extracted from the physical intersection of shared scalp electrodes. Solid lines denote the chronic subjective tinnitus group; dashed lines denote healthy controls. Traces are color-coded to highlight the excluded site: Austin is rendered in blue, while all other six retained centres are displayed in neutral grey (Dublin, Ghent, Illinois, Regensburg, Tübingen, and Zürich). (A) Waveforms averaged across the sensor region of interest (ROI) defining the early positive omission cluster. (B) Waveforms averaged across the sensor ROI defining the late negative omission cluster. Across both ROIs, the six retained centres exhibit consistent, time-locked evoked morphologies following stimulus expectation violation (1.25–1.70 s). In contrast, recordings from Austin show attenuated evoked excursions and elevated baseline noise, supporting site-level quality control exclusion.

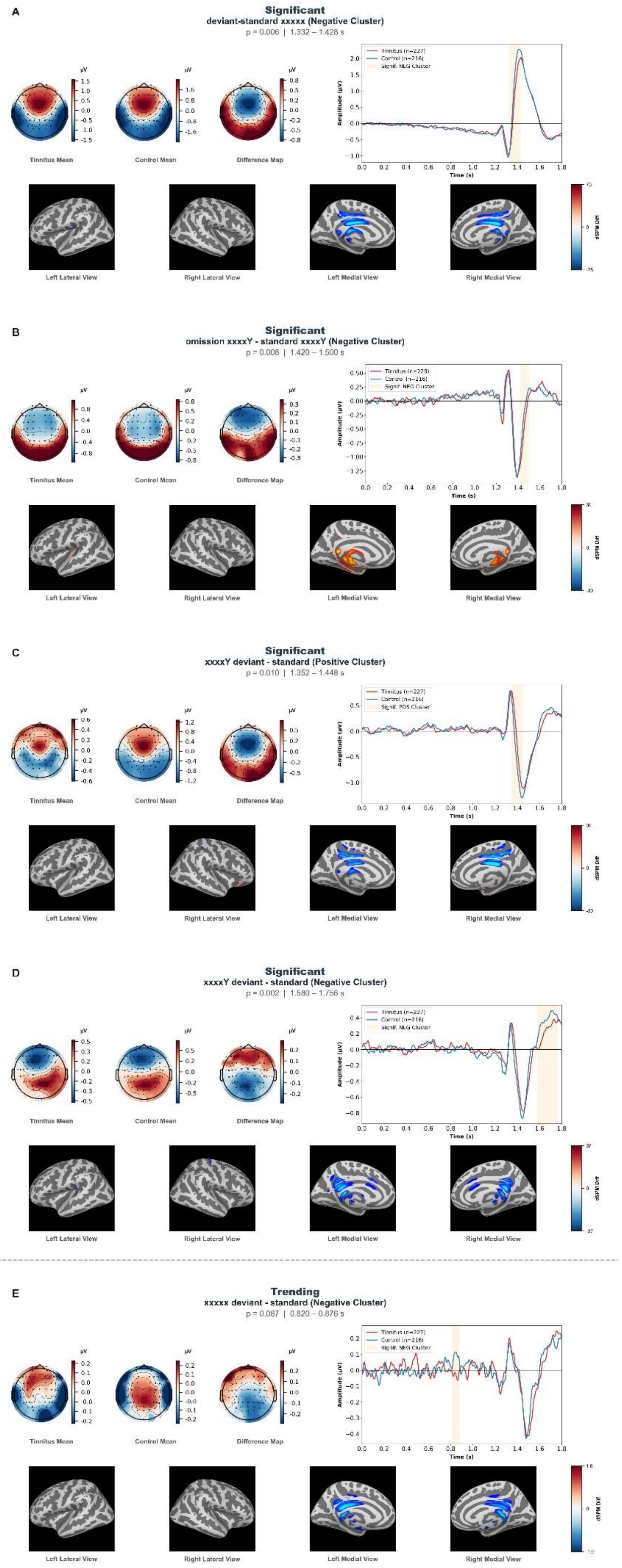
A
Significant
deviant-standard xxxxx (Negative Cluster)
p = 0.006 | 1.332 – 1.428 s
Tinnitus Mean
Control Mean
Difference Map
Tinnitus (n=227)
Control (n=216)
Signif. NEG Cluster
Amplitude (µV)
Time (s)
Left Lateral View
Right Lateral View
Left Medial View
Right Medial View
dSPM Diff
B
Significant
omission xxxxY - standard xxxxY (Negative Cluster)
p = 0.008 | 1.420 – 1.500 s
Tinnitus Mean
Control Mean
Difference Map
Tinnitus (n=228)
Control (n=216)
Signif. NEG Cluster
Amplitude (µV)
Time (s)
Left Lateral View
Right Lateral View
Left Medial View
Right Medial View
dSPM Diff
C
Significant
xxxxY deviant - standard (Positive Cluster)
p = 0.010 | 1.352 – 1.448 s
Tinnitus Mean
Control Mean
Difference Map
Tinnitus (n=227)
Control (n=216)
Signif. POS Cluster
Amplitude (µV)
Time (s)
Left Lateral View
Right Lateral View
Left Medial View
Right Medial View
dSPM Diff
D
Significant
xxxxY deviant - standard (Negative Cluster)
p = 0.002 | 1.580 – 1.756 s
Tinnitus Mean
Control Mean
Difference Map
Tinnitus (n=227)
Control (n=216)
Signif. NEG Cluster
Amplitude (µV)
Time (s)
Left Lateral View
Right Lateral View
Left Medial View
Right Medial View
dSPM Diff
E
Trending
xxxxx deviant - standard (Negative Cluster)
p = 0.087 | 0.820 – 0.876 s
Tinnitus Mean
Control Mean
Difference Map
Tinnitus (n=227)
Control (n=216)
Signif. NEG Cluster
Amplitude (µV)
Time (s)
Left Lateral View
Right Lateral View
Left Medial View
Right Medial View
dSPM Diff

**Supplementary Figure S3. Uncontrolled mass-univariate spatiotemporal clusters and cortical source reconstructions.**

Event-related potential group differences between chronic subjective tinnitus cases ($n = 226$) and controls ($n = 214$) in the unadjusted (uncontrolled) mass-univariate analysis. For each panel, top rows display unadjusted sensor-space scalp potential topographies ($\mu$V) for the Tinnitus group mean, Control group mean, and the corresponding difference map, alongside group ERP waveforms across the 0.0–1.8-s epoch (tinnitus: red; control: blue; orange shading: significant cluster window). Difference maps and waveforms reflect the signed contrast (Tinnitus $-$ Control), where positive voltage clusters indicate higher amplitude trajectories in the tinnitus cohort and negative voltage clusters indicate lower or inverted trajectory deflections relative to controls. Bottom rows depict exploratory dynamic statistical parametric mapping (dSPM) cortical source reconstructions mapped onto the template inflated *fsaverage* surface across lateral and medial cortical views. **(A)** Significant negative cluster in the *deviant-standard xxxxx* contrast (local deviance; two-tailed cluster permutation $P = .006$, peak $t_{442} = -4.794$, 1.332–1.428 s). **(B)** Significant negative cluster in the *omission xxxxY-standard xxxxY* contrast (within-block omission; $P = .008$, peak $t_{443} = -3.83$, 1.420–1.500 s). **(C)** Significant positive cluster in the *xxxxY deviant-standard* contrast (global deviance; $P = .009$, peak $t_{442} = 4.209$, 1.352–1.448 s). **(D)** Significant negative cluster in the *xxxxY deviant-standard* contrast (global deviance; $P = .002$, peak $t_{442} = -4.477$, 1.580–1.756 s). **(E)** Trending negative cluster in the *xxxxx deviant-standard* contrast (global deviance; $P = .087$, peak $t_{442} = -3.490$, 0.820–0.876 s).

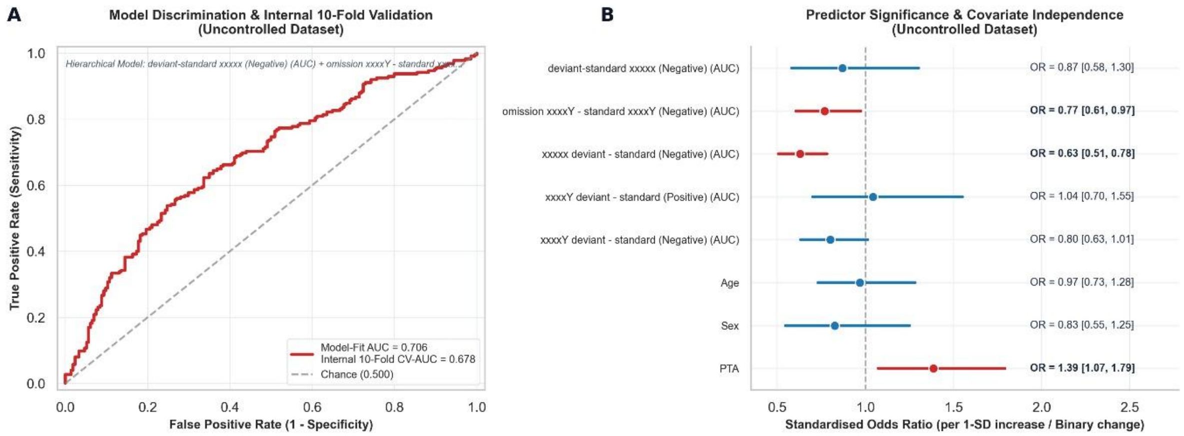


**Supplementary Figure S4. Diagnostic prediction performance and covariate independence in the uncontrolled dataset.**

Out-of-sample diagnostic classification and predictor contributions evaluated on unadjusted electrophysiological features. **(A)** Receiver operating characteristic (ROC) curve showing out-of-sample classification discrimination for the integrated multivariate logistic regression model (incorporating unadjusted spatiotemporal ERP AUC features, Age, Sex, and pure-tone hearing loss [PTA4]). The solid red curve depicts 10-fold cross-validated performance (CV-AUC = 0.678, 95% CI [0.627, 0.726]; Model-Fit AUC = 0.706) relative to chance discrimination (dashed diagonal line; 0.500). **(B)** Forest plot of standardised odds ratios (OR) per 1-SD increase in continuous variables or binary unit change in categorical variables. Points represent estimated odds ratios, and horizontal bars denote 95% confidence intervals derived from the final model fit. Statistically significant predictors ($P < .05$) are highlighted in red; non-significant predictors are displayed in blue. Vertical dashed line indicates the null effect threshold (OR = 1.0). Exact numerical odds ratios and 95% confidence bounds for each candidate predictor are provided along the right axis.

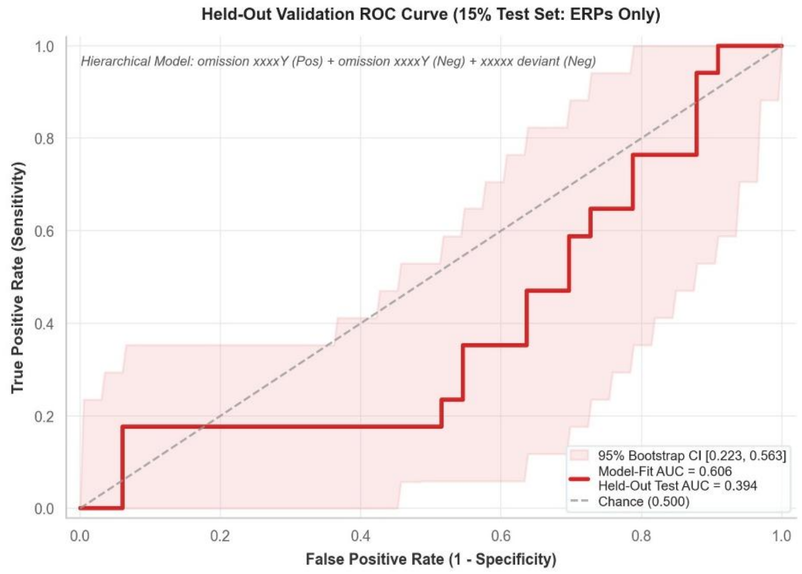


**Supplementary Figure S5. Held-out validation ROC curve for the ERP-only classifier in the 15% test set.** Receiver operating characteristic (ROC) curve evaluated in the untouched 15% held-out test subset for the multivariate ERP-only model (combining the *omission xxxxY-standard xxxxY* early positive and late negative cluster AUC features and the *xxxxx deviant-standard* negative cluster AUC feature; model-fit AUC = 0.606, held-out test AUC = 0.394, McFadden's $\Delta R^2 = 0.067$, $p(\Delta R^2) = .231$) against chance level (dashed grey line), with a shaded 95% bootstrap confidence interval (0.223–0.563).
