## Supplementary Tables for "Multicentre electrophysiology reveals the limits of auditory prediction responses as biomarkers of chronic tinnitus"

### Table 1. Equipment and devices across the 7 TIDE sites

| | | UTA | UIUC | TCD | BCG | UZH | UKT | UR |
|---|---|---|---|---|---|---|---|---|
| **Audiometry** | Headset | RadioEar DD450 | HDA200 | HDA300 | HDA200 | HDA200 | HDA300 | HDA300 |
| | Audiometer | GSI (Audiostar) | Equinox (Interacoustics) | Hekto (Horentek) | (MedRx) | RME UCX II | AT1000 (Auritec) | Astera 2 (Otometrics) |
| | dB steps (dB) | 5 dB | 5 dB | 5 dB | 5 dB | 1 dB | 1 dB | 5 dB |
| | Max freq (kHz) and output (dB HL) | 16 (60 dB) | 16 (60 dB) | 12.5 (95 dB) | 16 (60 dB) | 12.5 (90 dB) | 14 (75 dB) | 16 (40 dB) |
| | Tinnitus Loudness (dB HL / SL /SPL) | 2 (dB SL and HL) | dB HL and dB SL | dB HL | dB HL | dB SPL | dB HL | dB HL |
| | Frequency steps | 50 Hz | 0.25–8 kHz: ½ octave; >8 kHz: ⅙ octave | 25 Hz | 1 Hz | 1 Hz | At PTA frequency | 10 Hz |
| | Loudness steps | 1 dB | 2 dB | 5 dB | 1 dB | 1 dB | 1dB | 5 dB |
| | Inserts | EARtone GOLD 3A (3M) | EARtone GOLD 3A (3M) | ER2 (Etymotic) | ER2 (Etymotic) | ER2 (Etymotic) | ER2 (Etymotic) | ER2 (Etymotic) |
| | Acoustic chamber | Yes | No | No | No | Yes | Yes | No |
| **Electroencephalography** | EEG system | EGI Philips (Magstim) | SynAmps 2 (Neuroscan) | ActiveTwo (BioSemi) | EEGoMylab (AntNeuro) | ActiChamp (Brainproducts) | ActiChamp (Brainproducts) | ActiChamp (Brainproducts) |
| | Electrodes | Passive | Passive | Active | Passive | Active | Active | Passive |
| | Electrode cap | Hydrocel GSN 64 (EGI) | Quik-Cap Neo Net 64 (Compumedics NeuroScan) | 64 (ECI) | Waveguard original cap 64ch 10/10 shildes tyco 68 (ANT-neuro) | actiCAP slim 64 (Brainproducts) | actiCAP slim 64 (Brainproducts) | Braincap TMS 64 (Brainproducts) |
| | Ref. Electrode | Cz | Between Cz and CPz | Cz | CPz | Fz | FCz | FCz |
| | Blink recording | Yes | Yes | No | No | Yes | No | No |
| | Line noise (Hz) | 60 | 60 | 50 | 50 | 50 | 50 | 50 |
| **Presentation** | OS | Win 10 | Win 10 | Win 10 | | Win 11 | Win 10 | Win 10 |
| | Psychopy version | 2021.2.3 | 2023.2.3 | 2021.2.3 | 2021.2.3 | 2022.2.5 | 2021.2.3 | 2021.2.3 |
| | Sound Calibration | CAL250 calibrator | B&K 4231 / CAL250 calibrator | GRAS 43AG Ear & Cheek Simulator | GRAS 43AG Ear & Cheek Simulator | B&K 4152 / sound level meter (NTi Audio XLR II) | B&K 4152 KEMAR | sound level meter (NTi Audio XLR) |
| | CPU | Intel Core i3-41060 3.60 GHz | Intel (R) Xeon (R) E-2146G 3.60 GHz | i5 3.0 GHz | Intel(R) Pentium(R) Gold G5400 3.70 GHz | i7 10th gen | AMD Ryzen 3 5300U 2.60 GHz | i7 2.9 GHz |
| | RAM | 4 GB | 32 GB | 16 GB | 4 GB | 16 GB | 8 GB | 16 GB |

**Table 2**

**Participant inclusion and exclusions by centre**

| Centre | Eligible, n | Complete task datasets, n | Excluded after acquisition, n | Final tinnitus, n | Final controls, n | Final total, n |
|---|---|---|---|---|---|---|
| **Austin** | 83 | 82 | 82 | 0 | 0 | 0 |
| **Dublin** | 87 | 83 | 4 | 38 | 41 | 79 |
| **Ghent** | 83 | 77 | 13 | 35 | 29 | 64 |
| **Illinois** | 83 | 83 | 13 | 35 | 35 | 70 |
| **Regensburg** | 80 | 80 | 5 | 39 | 36 | 75 |
| **Tübingen** | 81 | 79 | 0 | 40 | 39 | 79 |
| **Zürich** | 87 | 78 | 5 | 39 | 34 | 73 |
| **Total** | 584 | 562 | 122 | 226 | 214 | 440 |

**Table 3**

**Spatiotemporal Cluster Statistics by Uncontrolled and Controlled Covariates**

| Contrast | Uncontrolled | Site | Sex | Age | PTA4 | Age, Sex | Age, Sex, PTA4 | Age, Sex, PTA4, Site | Age, Sex, PTA4, Site, HADS |
|---|---|---|---|---|---|---|---|---|---|
| ***deviant - standard xxxxx*** | | | | | | | | | |
| *Positive Cluster* | **[1.340, 1.424] s t = 4.22, p = .018*** | **[1.340, 1.420] s t = 4.12, p = .023*** | **[1.348, 1.420] s t = 3.79, p = .043*** | [1.352, 1.408] s t = 3.03, p = .385 | [1.348, 1.408] s t = 3.19, p = .203 | [0.032, 0.116] s t = 3.73, p = .685 | [1.680, 1.756] s t = 3.10, p = .902 | [1.684, 1.756] s t = 3.09, p = .909 | [1.684, 1.744] s t = 3.00, p = .953 |
| *Negative Cluster* | **[1.332, 1.428] s t = -4.79, p = .006*** | **[1.332, 1.424] s t = -4.65, p = .007*** | **[1.332, 1.424] s t = -4.25, p = .014*** | [1.328, 1.416] s t = -3.55, p = .105 | [1.328, 1.416] s t = -3.66, p = .070 | [1.360, 1.412] s t = -3.07, p = .518 | [1.360, 1.404] s t = -2.92, p = .877 | [1.360, 1.404] s t = -2.91, p = .877 | [1.276, 1.316] s t = -3.04, p = .985 |
| ***deviant - standard xxxxY*** | | | | | | | | | |
| *Positive Cluster* | [1.264, 1.300] s t = 3.66, p = .241 | [1.424, 1.488] s t = 3.32, p = .277 | [1.264, 1.300] s t = 3.69, p = .385 | [1.264, 1.308] s t = 3.98, p = .122 | [1.264, 1.304] s t = 3.72, p = .287 | [1.264, 1.308] s t = 3.94, p = .150 | [1.264, 1.304] s t = 3.85, p = .194 | [1.264, 1.304] s t = 3.85, p = .194 | [1.264, 1.316] s t = 4.16, p = .098 |
| *Negative Cluster* | [1.600, 1.672] s t = -3.42, p = .121 | [1.600, 1.672] s t = -3.33, p = .149 | [1.600, 1.672] s t = -3.25, p = .353 | [1.600, 1.672] s t = -3.18, p = .374 | [1.600, 1.672] s t = -3.28, p = .216 | [0.824, 0.872] s t = -3.35, p = .635 | [0.820, 0.872] s t = -3.48, p = .406 | [0.820, 0.872] s t = -3.48, p = .406 | [0.824, 0.872] s t = -3.31, p = .592 |
| ***omission xxxxx - standard omission*** | | | | | | | | | |
| *Positive Cluster* | [0.988, 1.044] s t = 4.62, p = .326 | [0.988, 1.044] s t = 4.58, p = .338 | [0.984, 1.040] s t = 4.50, p = .574 | [0.988, 1.044] s t = 4.61, p = .434 | [0.780, 0.832] s t = 3.38, p = .440 | [0.996, 1.040] s t = 4.46, p = .728 | [0.996, 1.040] s t = 4.49, p = .756 | [0.996, 1.040] s t = 4.48, p = .756 | [0.780, 0.828] s t = 3.23, p = .615 |
| *Negative Cluster* | [0.984, 1.044] s t = -3.74, p = .202 | [0.984, 1.044] s t = -3.77, p = .203 | [0.984, 1.044] s t = -3.79, p = .334 | [0.984, 1.044] s t = -3.63, p = .321 | [0.984, 1.044] s t = -3.69, p = .495 | [0.984, 1.044] s t = -3.70, p = .340 | [0.984, 1.048] s t = -3.72, p = .419 | [0.984, 1.048] s t = -3.72, p = .409 | [0.984, 1.044] s t = -3.79, p = .375 |
| ***omission xxxxY - standard omission*** | | | | | | | | | |
| *Positive Cluster* | [0.520, 0.560] s t = 3.73, p = .254 | [0.520, 0.552] s t = 3.68, p = .312 | [0.520, 0.560] s t = 3.75, p = .404 | [0.428, 0.460] s t = 3.17, p = .496 | [0.520, 0.552] s t = 3.62, p = .441 | [0.520, 0.552] s t = 3.62, p = .503 | [0.520, 0.552] s t = 3.58, p = .494 | [0.520, 0.552] s t = 3.58, p = .494 | [0.520, 0.556] s t = 3.86, p = .323 |
| *Negative Cluster* | [1.756, 1.800] s t = -3.59, p = .590 | [1.760, 1.800] s t = -3.60, p = .630 | [0.660, 0.700] s t = -3.26, p = .655 | [1.760, 1.800] s t = -3.82, p = .658 | [1.760, 1.800] s t = -3.69, p = .840 | [1.760, 1.800] s t = -3.86, p = .604 | [1.764, 1.800] s t = -3.83, p = .634 | [1.764, 1.800] s t = -3.83, p = .634 | [1.764, 1.800] s t = -4.04, p = .433 |

| | | | | | | | | | |
|---|---|---|---|---|---|---|---|---|---|
| ***omission xxxxx - standard xxxxx*** | | | | | | | | | |
| *Positive Cluster* | [0.760, 0.808] s<br>t = 3.55, p = .393 | [0.760, 0.808] s<br>t = 3.47, p = .442 | [0.760, 0.804] s<br>t = 3.31, p = .801 | [0.760, 0.808] s<br>t = 3.22, p = .620 | [0.756, 0.812] s<br>t = 3.37, p = .481 | [0.760, 0.804] s<br>t = 3.01, p = .877 | [0.756, 0.804] s<br>t = 3.05, p = .808 | [0.756, 0.804] s<br>t = 3.05, p = .808 | [0.760, 0.804] s<br>t = 3.24, p = .685 |
| *Negative Cluster* | [0.612, 0.652] s<br>t = -3.42, p = .693 | [0.612, 0.652] s<br>t = -3.35, p = .666 | [0.612, 0.652] s<br>t = -3.43, p = .846 | [0.612, 0.652] s<br>t = -3.39, p = .803 | [1.604, 1.636] s<br>t = -3.98, p = .757 | [0.612, 0.652] s<br>t = -3.38, p = .771 | [0.608, 0.652] s<br>t = -3.22, p = .836 | [0.608, 0.652] s<br>t = -3.22, p = .846 | [0.608, 0.648] s<br>t = -3.27, p = .853 |
| ***omission xxxxY - standard xxxxY*** | | | | | | | | | |
| *Positive Cluster* | **[1.420, 1.520] s<br>t = 3.80, p = .003*** | **[1.420, 1.520] s<br>t = 3.81, p = .004*** | **[1.420, 1.512] s<br>t = 3.63, p = .014*** | **[1.212, 1.300] s<br>t = 4.24, p = .011*** | **[1.244, 1.296] s<br>t = 4.05, p = .029*** | **[1.248, 1.300] s<br>t = 4.29, p = .019*** | **[1.244, 1.300] s<br>t = 4.19, p = .020*** | **[1.244, 1.300] s<br>t = 4.18, p = .021*▲** | **[1.248, 1.300] s<br>t = 4.19, p = .033*** |
| *Negative Cluster* | **[1.420, 1.500] s<br>t = -3.80, p = .008*** | **[1.416, 1.496] s<br>t = -3.83, p = .009*** | **[1.416, 1.496] s<br>t = -3.54, p = .024*** | [1.240, 1.300] s<br>t = -3.80, p = .130 | **[1.556, 1.676] s<br>t = -3.29, p = .015*** | [1.236, 1.300] s<br>t = -3.77, p = .108 | **[1.556, 1.676] s<br>t = -3.27, p = .041*** | **[1.556, 1.676] s<br>t = -3.26, p = .044*▲** | **[1.556, 1.676] s<br>t = -3.20, p = .033*** |
| ***xxxxx (deviant - standard)*** | | | | | | | | | |
| *Positive Cluster* | [0.460, 0.500] s<br>t = 3.37, p = .377 | [0.460, 0.500] s<br>t = 3.22, p = .471 | [0.460, 0.500] s<br>t = 3.37, p = .427 | [0.460, 0.500] s<br>t = 3.52, p = .383 | [0.792, 0.876] s<br>t = 3.38, p = .375 | [0.456, 0.500] s<br>t = 3.50, p = .273 | [0.792, 0.880] s<br>t = 3.37, p = .329 | [0.792, 0.880] s<br>t = 3.37, p = .328 | [0.456, 0.496] s<br>t = 3.86, p = .291 |
| *Negative Cluster* | [0.820, 0.876] s<br>t = -3.49, p = .087 | [0.820, 0.876] s<br>t = -3.45, p = .096 | [0.816, 0.876] s<br>t = -3.36, p = .160 | [0.820, 0.876] s<br>t = -3.58, p = .098 | [0.816, 0.876] s<br>t = -3.64, p = .077 | [0.816, 0.876] s<br>t = -3.48, p = .125 | [0.816, 0.876] s<br>t = -3.54, p = .086 | [0.816, 0.876] s<br>t = -3.54, p = .086 | [0.820, 0.876] s<br>t = -3.54, p = .175 |
| ***xxxxY (deviant - standard)*** | | | | | | | | | |
| *Positive Cluster* | **[1.352, 1.448] s<br>t = 4.21, p = .010*** | **[1.352, 1.444] s<br>t = 4.13, p = .014*** | **[1.356, 1.444] s<br>t = 3.83, p = .031*** | [1.356, 1.436] s<br>t = 3.12, p = .259 | [1.356, 1.428] s<br>t = 3.35, p = .129 | [0.032, 0.116] s<br>t = 4.26, p = .325 | [1.356, 1.424] s<br>t = 2.81, p = .530 | [1.356, 1.424] s<br>t = 2.80, p = .554 | [1.188, 1.252] s<br>t = 3.48, p = .714 |
| *Negative Cluster* | **[1.580, 1.756] s<br>t = -4.48, p = .002*** | **[1.348, 1.448] s<br>t = -5.06, p = .005*** | **[1.356, 1.448] s<br>t = -4.76, p = .009*** | [1.356, 1.456] s<br>t = -3.95, p = .059 | **[1.348, 1.448] s<br>t = -4.11, p = .041*** | [1.360, 1.468] s<br>t = -3.65, p = .102 | [1.356, 1.452] s<br>t = -3.54, p = .134 | [1.356, 1.452] s<br>t = -3.53, p = .135 | [1.356, 1.424] s<br>t = -3.34, p = .253 |

*Note. Values inside brackets show the cluster's temporal duration. t represents peak t-statistic value.*

** $p < .05$. Bold fields indicate clusters that reached statistical significance.* ▲ After a Bonferroni correction for multiple comparisons, both cluster do not survive.

**Table 4**

**Hierarchical Logistic Regression Model Results - Controlled Dataset**

| Predictor | Internal CV AUC [95% CI] | Model Fit AUC | Model P-value | Standardised β | Odds Ratio | Predictor P-value | p_fdr | Δ $R^2$ | p (Δ $R^2$) |
|---|---|---|---|---|---|---|---|---|---|
| **Behavioural & Demographic Variables** | | | | | | | | | |
| Age | .587 [.537, .639] | .594 | < .001 | **.344** | **1.410** | **< .001*** | **-** | .021 | **< .001*** |
| Sex | .512 [.457, .567] | .554 | .023 | **-.218** | **.804** | **.023*** | **-** | .009 | **.023*** |
| PTA4 | .596 [.545, .649] | .604 | < .001 | **.411** | **1.508** | **< .001*** | **-** | .028 | **< .001*** |
| HADS Anxiety | .608 [.556, .662] | .615 | < .001 | **.415** | **1.514** | **< .001*** | **-** | .029 | **< .001*** |
| HADS Depression | .591 [.537, .646] | .599 | .002 | **.304** | **1.355** | **.003*** | **-** | .016 | **.002*** |
| **Clinical Baseline Model (Age + Sex + PTA4)** | | | | | | | | | |
| Age | .608 [.551, .662] | | | .164 | 1.178 | .171 | .171 | | - |
| Sex | .608 [.551, .662] | | | -.180 | .835 | .067 | .101 | | - |
| PTA4 | .608 [.551, .662] | | | **.298** | **1.347** | **.017*** | **.052** | | - |
| *Overall Model Diagnostics (Clinical Baseline)* | *.608 [.551, .662]* | *.627* | *< .001* | | | | | *.037* | **< .001*** |
| **ERP Only** | | | | | | | | | |
| omission *xxxxY* - standard *xxxxY* (Positive Cluster) PTP | .485 [.433, .541] | .516 | .586 | -.052 | .949 | .587 | .587 | .000 | .586 |
| omission *xxxxY* - standard *xxxxY* (Negative Cluster) PTP | .522 [.466, .576] | .532 | .143 | .141 | 1.152 | .147 | .221 | .004 | .143 |
| *xxxxx* deviant - standard (Negative Cluster) PTP | .492 [.432, .539] | .511 | .525 | .061 | 1.063 | .525 | .587 | .001 | .525 |
| omission *xxxxY* - standard *xxxxY* (Positive Cluster) AUC | .581 [.524, .634] | .587 | < .001 | **.332** | **1.393** | **< .001*** | **.003** | .019 | **< .001*** |
| omission *xxxxY* - standard *xxxxY* (Negative Cluster) AUC | .569 [.515, .617] | .570 | .013 | **-.246** | **.782** | **.016*** | **.032** | .010 | **.013*** |
| *xxxxx* deviant - standard (Negative Cluster) AUC | .591 [.535, .643] | .593 | < .001 | **-.392** | **.676** | **< .001*** | **< .001** | .026 | **< .001*** |
| **ERP + Age + Sex + PTA4** | | | | | | | | | |
| omission *xxxxY* - standard *xxxxY* (Positive Cluster) PTP | .604 [.549, .656] | .629 | < .001 | -.062 | .940 | .538 | .538 | .037 | **< .001*** |
| omission *xxxxY* - standard *xxxxY* (Negative Cluster) PTP | .617 [.564, .668] | .636 | < .001 | .180 | 1.197 | .074 | .103 | .039 | **< .001*** |
| *xxxxx* deviant - standard (Negative Cluster) PTP | .609 [.552, .662] | .629 | < .001 | .132 | 1.141 | .191 | .229 | .039 | **< .001*** |
| omission *xxxxY* - standard *xxxxY* (Positive Cluster) AUC | .646 [.596, .696] | .659 | < .001 | **.365** | **1.441** | **< .001*** | **.003** | .039 | **< .001*** |

| Predictor | Internal CV AUC [95% CI] | Model Fit AUC | Model P-value | Standardised β | Odds Ratio | Predictor P-value | p_fdr | Δ $R^2$ | p (Δ $R^2$) |
|---|---|---|---|---|---|---|---|---|---|
| omission *xxxxY* - standard *xxxxY* (Negative Cluster) AUC | .624 [.574, .678] | .641 | < .001 | **-.267** | **.765** | **.011*** | **.033** | .038 | **< .001*** |
| *xxxxx* deviant - standard (Negative Cluster) AUC | .661 [.611, .709] | .678 | < .001 | **-.436** | **.646** | **< .001*** | **< .001** | .041 | **< .001*** |
| **ERP + Age + Sex + PTA4 + HADS** | | | | | | | | | |
| omission *xxxxY* - standard *xxxxY* (Positive Cluster) PTP | .653 [.601, .703] | .674 | < .001 | -.068 | .935 | .513 | .537 | .039 | **< .001*** |
| omission *xxxxY* - standard *xxxxY* (Negative Cluster) PTP | .663 [.613, .712] | .681 | < .001 | .180 | 1.198 | .080 | .109 | .039 | **< .001*** |
| *xxxxx* deviant - standard (Negative Cluster) PTP | .655 [.601, .703] | .676 | < .001 | .119 | 1.127 | .245 | .300 | .039 | **< .001*** |
| omission *xxxxY* - standard *xxxxY* (Positive Cluster) AUC | .681 [.632, .728] | .695 | < .001 | **.355** | **1.427** | **< .001*** | **.005** | .037 | **< .001*** |
| omission *xxxxY* - standard *xxxxY* (Negative Cluster) AUC | .675 [.625, .726] | .690 | < .001 | **-.284** | **.753** | **.010*** | **.027** | .039 | **< .001*** |
| *xxxxx* deviant - standard (Negative Cluster) AUC | .692 [.643, .739] | .708 | < .001 | **-.424** | **.654** | **< .001*** | **.002** | .036 | **< .001*** |
| **Combined Multivariate ERPs (PTP) - ERP Only** | | | | | | | | | |
| omission *xxxxY* - standard *xxxxY* (Positive Cluster) PTP | | | | -.126 | .882 | .226 | .271 | | - |
| omission *xxxxY* - standard *xxxxY* (Negative Cluster) PTP | | | | .180 | 1.197 | .112 | .169 | | - |
| *xxxxx* deviant - standard (Negative Cluster) PTP | | | | .019 | 1.020 | .856 | .856 | | - |
| *Overall Model Diagnostics* | *.523 [.468, .580]* | *.553* | *.305* | | | | | *.006* | *.305* |
| **Combined Multivariate ERPs (PTP) - ERP + Age + Sex + PTA4** | | | | | | | | | |
| omission *xxxxY* - standard *xxxxY* (Positive Cluster) PTP | | | | -.180 | .836 | .109 | .140 | | - |
| omission *xxxxY* - standard *xxxxY* (Negative Cluster) PTP | | | | .211 | 1.235 | .071 | .103 | | - |
| *xxxxx* deviant - standard (Negative Cluster) PTP | | | | .103 | 1.108 | .359 | .380 | | - |
| *Overall Model Diagnostics* | *.619 [.567, .667]* | *.646* | *< .001* | | | | | *.041* | **< .001*** |
| **Combined Multivariate ERPs (PTP) - ERP + Age + Sex + PTA4 + HADS** | | | | | | | | | |
| omission *xxxxY* - standard *xxxxY* (Positive Cluster) PTP | | | | -.189 | .828 | .101 | .130 | | - |
| omission *xxxxY* - standard *xxxxY* (Negative Cluster) PTP | | | | .221 | 1.247 | .066 | .104 | | - |
| *xxxxx* deviant - standard (Negative Cluster) PTP | | | | .090 | 1.094 | .430 | .474 | | - |

| Predictor | Internal CV AUC [95% CI] | Model Fit AUC | Model P-value | Standardised β | Odds Ratio | Predictor P-value | p_fdr | Δ R² | p (Δ R²) |
|---|---|---|---|---|---|---|---|---|---|
| *Overall Model Diagnostics* | *.663 [.612, .712]* | *.685* | *< .001* | | | | | *.039* | **< .001*** |
| **Combined Multivariate ERPs (AUC) - ERP Only** | | | | | | | | | |
| omission *xxxxY* - standard *xxxxY* (Positive Cluster) AUC | | | | **.278** | **1.320** | **.008*** | **.024** | | - |
| omission *xxxxY* - standard *xxxxY* (Negative Cluster) AUC | | | | -.202 | .817 | .061 | .123 | | - |
| *xxxxx* deviant - standard (Negative Cluster) AUC | | | | **-.383** | **.682** | **< .001*** | **.002** | | - |
| *Overall Model Diagnostics* | *.624 [.570, .673]* | *.636* | *< .001* | | | | | *.048* | **< .001*** |
| **Combined Multivariate ERPs (AUC) - ERP + Age + Sex + PTA4** | | | | | | | | | |
| omission *xxxxY* - standard *xxxxY* (Positive Cluster) AUC | | | | **.317** | **1.372** | **.004*** | **.016** | | - |
| omission *xxxxY* - standard *xxxxY* (Negative Cluster) AUC | | | | **-.229** | **.795** | **.041*** | **.087** | | - |
| *xxxxx* deviant - standard (Negative Cluster) AUC | | | | **-.429** | **.651** | **< .001*** | **< .001** | | - |
| *Overall Model Diagnostics* | *.689 [.640, .738]* | *.707* | *< .001* | | | | | *.045* | **< .001*** |
| **Combined Multivariate ERPs (AUC) - ERP + Age + Sex + PTA4 + HADS** | | | | | | | | | |
| omission *xxxxY* - standard *xxxxY* (Positive Cluster) AUC | | | | **.309** | **1.362** | **.006*** | **.019** | | - |
| omission *xxxxY* - standard *xxxxY* (Negative Cluster) AUC | | | | **-.244** | **.784** | **.038*** | **.076** | | - |
| *xxxxx* deviant - standard (Negative Cluster) AUC | | | | **-.417** | **.659** | **< .001*** | **.002** | | - |
| *Overall Model Diagnostics* | *.716 [.670, .763]* | *.732* | *< .001* | | | | | *.034* | **< .001*** |

*Note. CV AUC [95% CI] = 10-fold cross-validated Area Under Curve with 95% confidence intervals. Odds Ratio = Standardised Odds Ratio. p_fdr = false discovery rate corrected p-value. Δ R² = McFadden's Pseudo-R² change value. p (Δ R²) = statistical significance of the change in Pseudo-R² calculated via Likelihood Ratio testing.*

** p < .05. Bold fields indicate predictors/steps that reached statistical significance.*

**Table 5**

**Sensitivity Analysis: Model Robustness Across Leave-One-Site-Out Folds - Controlled Dataset**

| Predictor | Standardised β | SE | Odds Ratio [95% CI] | z-value | p-value |
|---|---|---|---|---|---|
| **Excluded Site: Dublin (Remaining N = 360)** | | | | | |
| omission *xxxxY* - standard *xxxxY* (Positive Cluster) AUC | .141 | 4.168 | 1.15 [0.91, 1.46] | 1.17 | .242 |
| **omission *xxxxY* - standard *xxxxY* (Negative Cluster) AUC** | **-.342** | **2.162** | **0.71 [0.54, 0.94]** | **-2.43** | **.015*** |
| ***xxxxx* deviant - standard (Negative Cluster) AUC** | **-.480** | **6.181** | **0.62 [0.48, 0.79]** | **-3.87** | **< .001*** |
| Age | .175 | .009 | 1.19 [0.91, 1.56] | 1.28 | .199 |
| Sex | -.115 | .228 | 0.89 [0.71, 1.11] | -1.02 | .310 |
| **PTA4** | **.385** | **.015** | **1.47 [1.10, 1.95]** | **2.65** | **.008*** |
| *Overall Model Diagnostics: Model-Fit AUC = .696, Pseudo-R² = .088, Internal 10-Fold CV-AUC = .659 [0.601, 0.713]* | | | | | |
| **Excluded Site: Ghent (Remaining N = 375)** | | | | | |
| **omission *xxxxY* - standard *xxxxY* (Positive Cluster) AUC** | **.402** | **3.966** | **1.49 [1.17, 1.90]** | **3.26** | **.001*** |
| **omission *xxxxY* - standard *xxxxY* (Negative Cluster) AUC** | **-.425** | **2.371** | **0.65 [0.49, 0.87]** | **-2.94** | **.003*** |
| ***xxxxx* deviant - standard (Negative Cluster) AUC** | **-.498** | **6.274** | **0.61 [0.48, 0.77]** | **-4.11** | **< .001*** |
| Age | -.020 | .009 | 0.98 [0.75, 1.29] | -.14 | .886 |
| Sex | -.192 | .228 | 0.83 [0.66, 1.03] | -1.69 | .092 |
| **PTA4** | **.582** | **.015** | **1.79 [1.33, 2.40]** | **3.87** | **< .001*** |
| *Overall Model Diagnostics: Model-Fit AUC = .735, Pseudo-R² = .122, Internal 10-Fold CV-AUC = .697 [0.645, 0.748]* | | | | | |
| **Excluded Site: Illinois (Remaining N = 370)** | | | | | |
| **omission *xxxxY* - standard *xxxxY* (Positive Cluster) AUC** | **.383** | **3.724** | **1.47 [1.15, 1.86]** | **3.14** | **.002*** |
| omission *xxxxY* - standard *xxxxY* (Negative Cluster) AUC | -.197 | 2.103 | 0.82 [0.65, 1.03] | -1.68 | .093 |

| Predictor | Standardised β | SE | Odds Ratio [95% CI] | z-value | p-value |
|---|---|---|---|---|---|
| ***xxxxx* deviant - standard (Negative Cluster) AUC** | **-.451** | **5.779** | **0.64 [0.50, 0.81]** | **-3.72** | **< .001*** |
| Age | .218 | .009 | 1.24 [0.95, 1.63] | 1.58 | .115 |
| Sex | -.211 | .228 | 0.81 [0.65, 1.01] | -1.86 | .063 |
| **PTA4** | **.381** | **.015** | **1.46 [1.10, 1.95]** | **2.61** | **.009*** |
| *Overall Model Diagnostics: Model-Fit AUC = .717, Pseudo-$R^2$ = .104, Internal 10-Fold CV-AUC = .690 [0.639, 0.744]* | | | | | |
| **Excluded Site: Regensburg (Remaining N = 364)** | | | | | |
| **omission *xxxxY* - standard *xxxxY* (Positive Cluster) AUC** | **.337** | **3.862** | **1.40 [1.10, 1.78]** | **2.73** | **.006*** |
| **omission *xxxxY* - standard *xxxxY* (Negative Cluster) AUC** | **-.306** | **2.151** | **0.74 [0.56, 0.97]** | **-2.18** | **.029*** |
| ***xxxxx* deviant - standard (Negative Cluster) AUC** | **-.474** | **6.176** | **0.62 [0.49, 0.79]** | **-3.81** | **< .001*** |
| **Age** | **.313** | **.009** | **1.37 [1.04, 1.79]** | **2.27** | **.023*** |
| **Sex** | **-.290** | **.229** | **0.75 [0.60, 0.94]** | **-2.54** | **.011*** |
| **PTA4** | **.289** | **.015** | **1.34 [1.01, 1.76]** | **2.06** | **.040*** |
| *Overall Model Diagnostics: Model-Fit AUC = .723, Pseudo-$R^2$ = .111, Internal 10-Fold CV-AUC = .693 [0.635, 0.742]* | | | | | |
| **Excluded Site: Tuebingen (Remaining N = 360)** | | | | | |
| **omission *xxxxY* - standard *xxxxY* (Positive Cluster) AUC** | **.468** | **4.002** | **1.60 [1.25, 2.03]** | **3.78** | **< .001*** |
| omission *xxxxY* - standard *xxxxY* (Negative Cluster) AUC | -.179 | 2.115 | 0.84 [0.64, 1.09] | -1.34 | .180 |
| ***xxxxx* deviant - standard (Negative Cluster) AUC** | **-.360** | **6.169** | **0.70 [0.55, 0.89]** | **-2.95** | **.003*** |
| Age | .184 | .010 | 1.20 [0.91, 1.58] | 1.31 | .189 |
| Sex | -.203 | .226 | 0.82 [0.65, 1.02] | -1.80 | .072 |
| **PTA4** | **.364** | **.014** | **1.44 [1.08, 1.92]** | **2.46** | **.014*** |

| Predictor | Standardised β | SE | Odds Ratio [95% CI] | z-value | p-value |
|---|---|---|---|---|---|
| *Overall Model Diagnostics: Model-Fit AUC = .696, Pseudo-R² = .088, Internal 10-Fold CV-AUC = .664 [0.609, 0.719]* | | | | | |
| **Excluded Site: Zuerich (Remaining N = 366)** | | | | | |
| **omission *xxxxY* - standard *xxxxY* (Positive Cluster) AUC** | **.466** | **3.980** | **1.59 [1.24, 2.04]** | **3.69** | **< .001*** |
| omission *xxxxY* - standard *xxxxY* (Negative Cluster) AUC | -.199 | 2.266 | 0.82 [0.62, 1.08] | -1.43 | .152 |
| ***xxxxx* deviant - standard (Negative Cluster) AUC** | **-.377** | **6.416** | **0.69 [0.54, 0.87]** | **-3.08** | **.002*** |
| Age | .173 | .009 | 1.19 [0.90, 1.57] | 1.22 | .224 |
| **Sex** | **-.309** | **.230** | **0.73 [0.59, 0.92]** | **-2.68** | **.007*** |
| **PTA4** | **.490** | **.016** | **1.63 [1.21, 2.20]** | **3.20** | **.001*** |
| *Overall Model Diagnostics: Model-Fit AUC = .726, Pseudo-R² = .119, Internal 10-Fold CV-AUC = .707 [0.655, 0.759]* | | | | | |

*Note. Excluded Site indicates the clinical centre omitted from the training dataset fold. CV-AUC [95% CI] = 10-fold cross-validated Area Under the Curve with 95% empirical bootstrap confidence intervals. Odds Ratio [95% CI] = Standardised Odds Ratio (per 1-SD increase / Binary change) with 95% confidence intervals.*

** p < .05. Bold fields indicate predictors that reached statistical significance.*

**Table 6**

**Calibration and subgroup discrimination of the integrated tinnitus classifier**

*The integrated model combined three ERP features with age, sex and PTA4 hearing thresholds (n = 439).*

#### A Overall calibration

| Measure | Estimate |
|---|---|
| Brier score | 0.2244 |
| Out-of-fold calibration slope | 0.8955 |
| Out-of-fold calibration intercept | −0.0814 |

#### B Subgroup discrimination

| Characteristic | Subgroup | n | CV ROC-AUC | 95% CI |
|---|---|---|---|---|
| Sex | Male | 209 | 0.690 | 0.620–0.760 |
| Sex | Female | 230 | 0.676 | 0.608–0.746 |
| Age | <50 years | 215 | 0.665 | 0.594–0.742 |
| Age | ≥50 years | 224 | 0.657 | 0.583–0.729 |
| PTA4 | <15 dB HL | 284 | 0.647 | 0.585–0.712 |
| PTA4 | ≥15 dB HL | 155 | 0.701 | 0.621–0.778 |

*Notes. Discrimination was evaluated using 10-fold cross-validation. Calibration measures were calculated from out-of-fold predicted probabilities. Subgroup estimates are descriptive; overlapping confidence intervals do not establish equivalent performance. ERP feature selection preceded cross-validation, so estimates are conditional on the selected features and do not validate the complete feature-discovery pipeline. Confidence intervals for calibration measures were not supplied.*

*Abbreviations: CI, confidence interval; CV, cross-validation; ERP, event-related potential; PTA4, four-frequency pure-tone average; dB HL, decibels hearing level; ROC-AUC, area under the receiver operating characteristic curve.*

**Table 7**

**Linear Regression Results Characterising Clinical Prediction - THI Score Models**

| Predictor | N | Standardised β | t-value | p-value | p_fdr | r coefficient | Δ $R^2$ | p (Δ $R^2$) | MAE | RMSE |
|---|---|---|---|---|---|---|---|---|---|---|
| **Behavioural & Demographic Variables** | | | | | | | | | | |
| Age (Years) | 226 | -.013 | -.191 | .849 | - | -.214 | .000 | .849 | 13.909 | 17.192 |
| Sex (Female vs. Male) | 225 | -.023 | -.339 | .735 | - | -.193 | .001 | .735 | 13.985 | 17.215 |
| Hearing Loss (PTA4) | 226 | -.013 | -.192 | .848 | - | -.195 | .000 | .848 | 13.901 | 17.155 |
| Tinnitus Loudness (VAS) | 218 | **.456** | **7.531** | **< .001*** | **-** | .438 | .208 | **< .001*** | 11.889 | 15.256 |
| **ERP Only** | | | | | | | | | | |
| omission *xxxxY* - standard *xxxxY* (Positive Cluster) (PTP) | 217 | .046 | .675 | .500 | .702 | -.161 | .002 | .500 | 13.797 | 17.089 |
| omission *xxxxY* - standard *xxxxY* (Positive Cluster) (AUC) | 217 | -.071 | -1.036 | .301 | .690 | -.053 | .005 | .301 | 13.706 | 17.060 |
| omission *xxxxY* - standard *xxxxY* (Negative Cluster) (PTP) | 217 | -.050 | -.735 | .463 | .702 | -.037 | .003 | .463 | 13.690 | 17.045 |
| omission *xxxxY* - standard *xxxxY* (Negative Cluster) (AUC) | 217 | .122 | 1.798 | .074 | .509 | .037 | .015 | .074 | 13.676 | 16.959 |
| *xxxxx* deviant - standard (Negative Cluster) (PTP) | 217 | -.025 | -.360 | .720 | .797 | -.182 | .001 | .720 | 13.779 | 17.083 |
| *xxxxx* deviant - standard (Negative Cluster) (AUC) | 217 | -.024 | -.345 | .731 | .797 | -.107 | .001 | .731 | 13.934 | 17.169 |
| **ERP + Age + Sex + PTA4** | | | | | | | | | | |
| omission *xxxxY* - standard *xxxxY* (Positive Cluster) (PTP) | 217 | .049 | .703 | .483 | .994 | -.250 | .001 | .980 | 13.927 | 17.233 |
| omission *xxxxY* - standard *xxxxY* (Positive Cluster) (AUC) | 217 | -.070 | -1.013 | .312 | .994 | -.166 | .001 | .991 | 13.836 | 17.212 |
| omission *xxxxY* - standard *xxxxY* (Negative Cluster) (PTP) | 217 | -.053 | -.755 | .451 | .994 | -.130 | .001 | .981 | 13.837 | 17.184 |

| Predictor | N | Standardised β | t-value | p-value | p_fdr | r coefficient | Δ R² | p (Δ R²) | MAE | RMSE |
|---|---|---|---|---|---|---|---|---|---|---|
| omission *xxxxY* - standard *xxxxY* (Negative Cluster) (AUC) | 217 | .125 | 1.813 | .071 | .745 | -.087 | .001 | .971 | 13.824 | 17.108 |
| *xxxxx* deviant - standard (Negative Cluster) (PTP) | 217 | -.026 | -.367 | .714 | .994 | -.240 | .001 | .985 | 13.916 | 17.224 |
| *xxxxx* deviant - standard (Negative Cluster) (AUC) | 217 | -.021 | -.301 | .764 | .994 | -.193 | .001 | .990 | 14.072 | 17.317 |
| **ERP + Age + Sex + PTA4 + VAS** | | | | | | | | | | |
| omission *xxxxY* - standard *xxxxY* (Positive Cluster) (PTP) | 217 | .056 | .895 | .372 | .701 | .421 | .217 | **< .001*** | 12.015 | 15.427 |
| omission *xxxxY* - standard *xxxxY* (Positive Cluster) (AUC) | 217 | -.028 | -.451 | .652 | .767 | .425 | .212 | **< .001*** | 12.025 | 15.383 |
| omission *xxxxY* - standard *xxxxY* (Negative Cluster) (PTP) | 217 | -.024 | -.379 | .705 | .783 | .422 | .214 | **< .001*** | 12.007 | 15.411 |
| omission *xxxxY* - standard *xxxxY* (Negative Cluster) (AUC) | 217 | **.123** | **2.022** | **.044*** | **.222** | .438 | .216 | **< .001*** | 11.927 | 15.272 |
| *xxxxx* deviant - standard (Negative Cluster) (PTP) | 217 | -.046 | -.737 | .462 | .701 | .424 | .218 | **< .001*** | 12.027 | 15.394 |
| *xxxxx* deviant - standard (Negative Cluster) (AUC) | 217 | -.086 | -1.386 | .167 | .558 | .418 | .223 | **< .001*** | 12.067 | 15.461 |
| **Combined Multivariate ERPs (PTP) - ERP Only** | | | | | | | | | | |
| omission *xxxxY* - standard *xxxxY* (Positive Cluster) (PTP) | 217 | .080 | 1.067 | .287 | .690 | - | - | - | - | - |
| omission *xxxxY* - standard *xxxxY* (Negative Cluster) (PTP) | 217 | -.077 | -.948 | .344 | .690 | - | - | - | - | - |
| *xxxxx* deviant - standard (Negative Cluster) (PTP) | 217 | -.011 | -.147 | .883 | .883 | - | - | - | - | - |
| *Overall Model Diagnostics* | *217* | - | - | *.643* | - | *-.073* | *.008* | *.643* | *13.728* | *17.151* |
| **Combined Multivariate ERPs (PTP) - ERP + Age + Sex + PTA4** | | | | | | | | | | |
| omission *xxxxY* - standard *xxxxY* (Positive Cluster) (PTP) | 217 | .091 | 1.154 | .250 | .994 | - | - | - | - | - |
| omission *xxxxY* - standard *xxxxY* (Negative Cluster) (PTP) | 217 | -.084 | -1.013 | .312 | .994 | - | - | - | - | - |

| Predictor | N | Standardised β | t-value | p-value | p_fdr | r coefficient | Δ R² | p (Δ R²) | MAE | RMSE |
|---|---|---|---|---|---|---|---|---|---|---|
| *xxxxx* deviant - standard (Negative Cluster) (PTP) | 217 | -.018 | -.220 | .826 | .994 | - | - | - | - | - |
| *Overall Model Diagnostics* | *217* | - | - | *.942* | - | *-.135* | *.002* | *.942* | *13.813* | *17.277* |
| **Combined Multivariate ERPs (PTP) - ERP + Age + Sex + PTA4 + VAS** | | | | | | | | | | |
| omission *xxxxY* - standard *xxxxY* (Positive Cluster) (PTP) | 217 | .089 | 1.283 | .201 | .574 | - | - | - | - | - |
| omission *xxxxY* - standard *xxxxY* (Negative Cluster) (PTP) | 217 | -.036 | -.492 | .623 | .767 | - | - | - | - | - |
| *xxxxx* deviant - standard (Negative Cluster) (PTP) | 217 | -.058 | -.825 | .410 | .701 | - | - | - | - | - |
| *Overall Model Diagnostics* | *217* | **-** | **-** | ***< .001**** | **-** | *.414* | *.216* | ***< .001**** | *12.040* | *15.495* |
| **Combined Multivariate ERPs (AUC) - ERP Only** | | | | | | | | | | |
| omission *xxxxY* - standard *xxxxY* (Positive Cluster) (AUC) | 217 | -.065 | -.947 | .345 | .690 | - | - | - | - | - |
| omission *xxxxY* - standard *xxxxY* (Negative Cluster) (AUC) | 217 | .118 | 1.731 | .085 | .509 | - | - | - | - | - |
| *xxxxx* deviant - standard (Negative Cluster) (AUC) | 217 | -.044 | -.634 | .527 | .702 | - | - | - | - | - |
| *Overall Model Diagnostics* | *217* | - | - | *.229* | - | *-.018* | *.020* | *.229* | *13.754* | *17.109* |
| **Combined ERPs (AUC) - ERP + Age + Sex + PTA4** | | | | | | | | | | |
| omission *xxxxY* - standard *xxxxY* (Positive Cluster) (AUC) | 217 | -.065 | -.924 | .356 | .994 | - | - | - | - | - |
| omission *xxxxY* - standard *xxxxY* (Negative Cluster) (AUC) | 217 | .121 | 1.743 | .083 | .745 | - | - | - | - | - |
| *xxxxx* deviant - standard (Negative Cluster) (AUC) | 217 | -.039 | -.559 | .577 | .994 | - | - | - | - | - |
| *Overall Model Diagnostics* | *217* | - | - | *.986* | - | *-.096* | *.001* | *.986* | *13.870* | *17.260* |
| **Combined ERPs (AUC) - ERP + Age + Sex + PTA4 + VAS** | | | | | | | | | | |

| **Predictor** | **N** | **Standardised β** | **t-value** | **p-value** | **p_fdr** | **r coefficient** | **Δ R²** | **p (Δ R²)** | **MAE** | **RMSE** |
|---|---|---|---|---|---|---|---|---|---|---|
| omission *xxxxY* - standard *xxxxY* (Positive Cluster) (AUC) | 217 | -.031 | -.499 | .618 | .767 | - | - | - | - | - |
| omission *xxxxY* - standard *xxxxY* (Negative Cluster) (AUC) | 217 | **.126** | **2.063** | **.040*** | **.222** | - | - | - | - | - |
| *xxxxx* deviant - standard (Negative Cluster) (AUC) | 217 | -.099 | -1.581 | .115 | .462 | - | - | - | - | - |
| *Overall Model Diagnostics* | *217* | **-** | **-** | ***< .001**** | **-** | *.430* | *.221* | ***< .001**** | *11.914* | *15.353* |

*Note. N = participant count inside the given model step. Standardised β = Standardised regression coefficient. r coefficient = 10-fold cross-validated correlation coefficient r evaluating internal model shrinkage; MAE and RMSE are computed from held-out folds. Δ $R^2$ = change in variance explained at the given hierarchical step. p (Δ $R^2$) = statistical significance of the change in $R^2$ calculated via hierarchical F-change testing. For Behavioural & Demographic Variables, unadjusted univariable statistics are computed independently. For ERP Only, Δ $R^2$ represents the unique variance explained by the ERP alone. For subsequent hierarchical blocks, Δ $R^2$ represents the incremental variance explained by the stepwise addition of demographic, hearing-level, and clinical scale covariates.*

** $p < .05$. Bold fields indicate predictors that reached statistical significance.*

**Table 8**

**Sensitivity Analysis Table - Tinnitus Severity (THI)**

**Multi-Site Sensitivity Analysis: Parameter Stability Across Leave-One-Site-Out Folds**

| Predictor | Standardised β | SE | Unstandardised B [95% CI] | t-value | p-value |
|---|---|---|---|---|---|
| **Excluded Site: Dublin (Remaining N = 179)** | | | | | |
| omission *xxxxY* - standard *xxxxY* (Negative Cluster) AUC | .097 | 23.153 | 33.339 [-12.359, 79.037] | 1.44 | .152 |
| Age (Years) | -.060 | .099 | -0.072 [-0.267, 0.123] | -.73 | .468 |
| Sex (Female vs. Male) | -.013 | 2.362 | -0.447 [-5.109, 4.215] | -.19 | .850 |
| Hearing Thresholds (PTA4) | -.110 | .136 | -0.183 [-0.451, 0.085] | -1.34 | .180 |
| **Subjective Loudness (VAS)** | **.476** | **.051** | **0.358 [0.258, 0.459]** | **7.02** | **< .001*** |
| *Overall Model Diagnostics: Model-Fit r = .483, MAE = 11.73, RMSE = 15.22, Internal 10-Fold CV r = .484 [0.006, 0.896]* | | | | | |
| **Excluded Site: Ghent (Remaining N = 183)** | | | | | |
| omission *xxxxY* - standard *xxxxY* (Negative Cluster) AUC | .061 | 22.338 | 20.244 [-23.839, 64.328] | .91 | .366 |
| Age (Years) | -.157 | .089 | -0.168 [-0.344, 0.008] | -1.88 | .062 |
| Sex (Female vs. Male) | .024 | 2.126 | 0.757 [-3.438, 4.952] | .36 | .722 |
| Hearing Thresholds (PTA4) | .062 | .119 | 0.087 [-0.148, 0.322] | .73 | .468 |
| **Subjective Loudness (VAS)** | **.446** | **.047** | **0.309 [0.216, 0.402]** | **6.55** | **< .001*** |
| *Overall Model Diagnostics: Model-Fit r = .464, MAE = 10.84, RMSE = 13.97, Internal 10-Fold CV r = .243 [-0.334, 0.737]* | | | | | |

| Predictor | Standardised β | SE | Unstandardised B [95% CI] | t-value | p-value |
|---|---|---|---|---|---|
| **Excluded Site: Illinois (Remaining N = 190)** | | | | | |
| **omission *xxxxY* - standard *xxxxY* (Negative Cluster) AUC** | **.159** | **21.377** | **51.669 [9.494, 93.844]** | **2.42** | **.017*** |
| Age (Years) | -.053 | .097 | -0.063 [-0.253, 0.128] | -.65 | .517 |
| Sex (Female vs. Male) | -.012 | 2.268 | -0.394 [-4.869, 4.082] | -.17 | .862 |
| Hearing Thresholds (PTA4) | -.064 | .124 | -0.100 [-0.346, 0.145] | -.81 | .422 |
| **Subjective Loudness (VAS)** | **.447** | **.050** | **0.334 [0.235, 0.433]** | **6.65** | **< .001*** |
| *Overall Model Diagnostics: Model-Fit r = .464, MAE = 11.81, RMSE = 15.01, Internal 10-Fold CV r = .299 [-0.138, 0.680]* | | | | | |
| **Excluded Site: Regensburg (Remaining N = 178)** | | | | | |
| omission *xxxxY* - standard *xxxxY* (Negative Cluster) AUC | .125 | 22.230 | 41.802 [-2.077, 85.681] | 1.88 | .062 |
| Age (Years) | -.087 | .097 | -0.106 [-0.297, 0.086] | -1.09 | .278 |
| Sex (Female vs. Male) | -.060 | 2.291 | -2.069 [-6.591, 2.454] | -.90 | .368 |
| Hearing Thresholds (PTA4) | -.068 | .140 | -0.121 [-0.398, 0.155] | -.87 | .387 |
| **Subjective Loudness (VAS)** | **.491** | **.050** | **0.373 [0.274, 0.472]** | **7.44** | **< .001*** |
| *Overall Model Diagnostics: Model-Fit r = .511, MAE = 11.73, RMSE = 14.81, Internal 10-Fold CV r = .478 [-0.013, 0.784]* | | | | | |
| **Excluded Site: Tuebingen (Remaining N = 177)** | | | | | |
| omission *xxxxY* - standard *xxxxY* (Negative Cluster) AUC | .091 | 23.141 | 31.058 [-14.621, 76.738] | 1.34 | .181 |

| **Predictor** | **Standardised β** | **SE** | **Unstandardised B [95% CI]** | **t-value** | **p-value** |
|---|---|---|---|---|---|
| Age (Years) | -.101 | .105 | -0.125 [-0.333, 0.083] | -1.19 | .237 |
| Sex (Female vs. Male) | -.029 | 2.386 | -0.999 [-5.708, 3.711] | -.42 | .676 |
| Hearing Thresholds (PTA4) | -.072 | .130 | -0.111 [-0.368, 0.147] | -.85 | .398 |
| **Subjective Loudness (VAS)** | **.468** | **.051** | **0.347 [0.246, 0.448]** | **6.79** | **< .001*** |
| *Overall Model Diagnostics: Model-Fit r = .469, MAE = 12.01, RMSE = 15.28, Internal 10-Fold CV r = .139 [-0.423, 0.663]* | | | | | |
| **Excluded Site: Zuerich (Remaining N = 178)** | | | | | |
| **omission *xxxxY* - standard *xxxxY* (Negative Cluster) AUC** | **.186** | **22.452** | **64.188 [19.871, 108.506]** | **2.86** | **.005*** |
| Age (Years) | .066 | .094 | 0.077 [-0.108, 0.262] | .83 | .410 |
| Sex (Female vs. Male) | -.011 | 2.203 | -0.363 [-4.710, 3.985] | -.16 | .869 |
| **Hearing Thresholds (PTA4)** | **-.159** | **.126** | **-0.249 [-0.497, -0.001]** | **-1.98** | **.049*** |
| **Subjective Loudness (VAS)** | **.505** | **.048** | **0.366 [0.271, 0.461]** | **7.62** | **< .001*** |
| *Overall Model Diagnostics: Model-Fit r = .527, MAE = 11.19, RMSE = 14.25, Internal 10-Fold CV r = .228 [-0.387, 0.703]* | | | | | |

*Note. Excluded Site indicates clinical centre omitted from the subcohort to test multi-centre parameter robustness. Standardised β and Unstandardised B [95% CI] are derived from linear regression models fitted to the remaining training cohort (N). Model-Fit r, MAE, and RMSE quantify in-sample explanatory fit; Internal 10-Fold CV r [95% CI] represents cross-validated shrinkage within the remaining cohort.*

**Table 9**

**Linear Regression Results Characterising Clinical Prediction - VAS Models**

| Predictor | N | Standardised β | t-value | p-value | p_fdr | r coefficient | Δ R² | p (Δ R²) | MAE | RMSE |
|---|---|---|---|---|---|---|---|---|---|---|
| **Behavioural & Demographic Variables** | | | | | | | | | | |
| Age (Years) | 218 | **.162** | **2.420** | **.016*** | **-** | .106 | .026 | **.016*** | 19.780 | 22.905 |
| Sex (Female vs. Male) | 217 | .002 | .030 | .976 | - | -.293 | .000 | .976 | 20.182 | 23.219 |
| Hearing Loss (PTA4) | 218 | **.170** | **2.535** | **.012*** | **-** | .105 | .029 | **.012*** | 19.742 | 22.925 |
| Tinnitus Distress (THI) | 218 | **.456** | **7.531** | **< .001*** | **-** | .439 | .208 | **< .001*** | 17.201 | 20.666 |
| **ERP Only** | | | | | | | | | | |
| omission *xxxxY* - standard *xxxxY* (Positive Cluster) (PTP) | 217 | -.003 | -.046 | .964 | .981 | -.206 | .000 | .964 | 20.227 | 23.225 |
| omission *xxxxY* - standard *xxxxY* (Positive Cluster) (AUC) | 217 | -.087 | -1.273 | .204 | .565 | -.096 | .007 | .204 | 20.098 | 23.246 |
| omission *xxxxY* - standard *xxxxY* (Negative Cluster) (PTP) | 217 | -.081 | -1.190 | .235 | .565 | -.042 | .007 | .235 | 20.207 | 23.202 |
| omission *xxxxY* - standard *xxxxY* (Negative Cluster) (AUC) | 217 | .020 | .291 | .771 | .928 | -.277 | .000 | .771 | 20.170 | 23.186 |
| *xxxxx* deviant - standard (Negative Cluster) (PTP) | 217 | .020 | .288 | .774 | .928 | -.179 | .000 | .774 | 20.263 | 23.413 |
| *xxxxx* deviant - standard (Negative Cluster) (AUC) | 217 | **.161** | **2.385** | **.018*** | **.178** | .065 | .026 | **.018*** | 20.010 | 22.982 |
| **ERP + Age + Sex + PTA4** | | | | | | | | | | |
| omission *xxxxY* - standard *xxxxY* (Positive Cluster) (PTP) | 217 | -.013 | -.194 | .846 | .971 | .051 | .034 | .060 | 19.940 | 23.192 |
| omission *xxxxY* - standard *xxxxY* (Positive Cluster) (AUC) | 217 | -.089 | -1.319 | .188 | .565 | .054 | .034 | .057 | 19.804 | 23.240 |
| omission *xxxxY* - standard *xxxxY* (Negative Cluster) (PTP) | 217 | -.062 | -.900 | .369 | .604 | .067 | .031 | .079 | 19.917 | 23.176 |

| Predictor | N | Standardised β | t-value | p-value | p_fdr | r coefficient | Δ R² | p (Δ R²) | MAE | RMSE |
|---|---|---|---|---|---|---|---|---|---|---|
| omission *xxxxY* - standard *xxxxY* (Negative Cluster) (AUC) | 217 | .003 | .048 | .962 | .971 | .057 | .034 | .063 | 19.868 | 23.126 |
| *xxxxx* deviant - standard (Negative Cluster) (PTP) | 217 | .043 | .614 | .540 | .809 | .035 | .035 | .053 | 19.906 | 23.267 |
| *xxxxx* deviant - standard (Negative Cluster) (AUC) | 217 | **.134** | **1.977** | **.049*** | **.565** | .098 | .026 | .127 | 19.855 | 23.043 |
| **ERP + Age + Sex + PTA4 + THI** | | | | | | | | | | |
| omission *xxxxY* - standard *xxxxY* (Positive Cluster) (PTP) | 217 | -.036 | -.589 | .557 | .603 | .435 | .210 | **< .001*** | 17.129 | 20.731 |
| omission *xxxxY* - standard *xxxxY* (Positive Cluster) (AUC) | 217 | -.058 | -.956 | .340 | .562 | .435 | .205 | **< .001*** | 17.095 | 20.737 |
| omission *xxxxY* - standard *xxxxY* (Negative Cluster) (PTP) | 217 | -.038 | -.618 | .537 | .603 | .434 | .207 | **< .001*** | 17.112 | 20.754 |
| omission *xxxxY* - standard *xxxxY* (Negative Cluster) (AUC) | 217 | -.055 | -.898 | .370 | .562 | .443 | .212 | **< .001*** | 17.029 | 20.621 |
| *xxxxx* deviant - standard (Negative Cluster) (PTP) | 217 | .055 | .886 | .376 | .562 | .432 | .210 | **< .001*** | 17.094 | 20.764 |
| *xxxxx* deviant - standard (Negative Cluster) (AUC) | 217 | **.144** | **2.398** | **.017*** | **.107** | .452 | .212 | **< .001*** | 16.939 | 20.547 |
| **Combined Multivariate ERPs (PTP) - ERP Only** | | | | | | | | | | |
| omission *xxxxY* - standard *xxxxY* (Positive Cluster) (PTP) | 217 | .027 | .365 | .715 | .928 | - | - | - | - | - |
| omission *xxxxY* - standard *xxxxY* (Negative Cluster) (PTP) | 217 | -.123 | -1.524 | .129 | .515 | - | - | - | - | - |
| *xxxxx* deviant - standard (Negative Cluster) (PTP) | 217 | .069 | .890 | .374 | .642 | - | - | - | - | - |
| *Overall Model Diagnostics* | *217* | - | - | *.491* | - | *-.056* | *.011* | *.491* | *20.384* | *23.569* |
| **Combined ERPs (PTP) - ERP + Age + Sex + PTA4** | | | | | | | | | | |
| omission *xxxxY* - standard *xxxxY* (Positive Cluster) (PTP) | 217 | .003 | .037 | .971 | .971 | - | - | - | - | - |
| omission *xxxxY* - standard *xxxxY* (Negative Cluster) (PTP) | 217 | -.100 | -1.233 | .219 | .565 | - | - | - | - | - |

| Predictor | N | Standardised β | t-value | p-value | p_fdr | r coefficient | Δ R² | p (Δ R²) | MAE | RMSE |
|---|---|---|---|---|---|---|---|---|---|---|
| *xxxxx* deviant - standard (Negative Cluster) (PTP) | 217 | .086 | 1.101 | .272 | .565 | - | - | - | - | - |
| *Overall Model Diagnostics* | *217* | - | - | *.073* | - | *.043* | *.032* | *.073* | *20.055* | *23.457* |
| **Combined ERPs (PTP) - ERP + Age + Sex + PTA4 + THI** | | | | | | | | | | |
| omission *xxxxY* - standard *xxxxY* (Positive Cluster) (PTP) | 217 | -.039 | -.565 | .573 | .603 | - | - | - | - | - |
| omission *xxxxY* - standard *xxxxY* (Negative Cluster) (PTP) | 217 | -.062 | -.855 | .393 | .562 | - | - | - | - | - |
| *xxxxx* deviant - standard (Negative Cluster) (PTP) | 217 | .094 | 1.358 | .176 | .440 | - | - | - | - | - |
| *Overall Model Diagnostics* | *217* | ***-*** | ***-*** | ***< .001**** | ***-*** | *.417* | *.208* | ***< .001**** | *17.269* | *20.990* |
| **Combined Multivariate ERPs (AUC) - ERP Only** | | | | | | | | | | |
| omission *xxxxY* - standard *xxxxY* (Positive Cluster) (AUC) | 217 | -.061 | -.892 | .373 | .642 | - | - | - | - | - |
| omission *xxxxY* - standard *xxxxY* (Negative Cluster) (AUC) | 217 | .002 | .023 | .981 | .981 | - | - | - | - | - |
| *xxxxx* deviant - standard (Negative Cluster) (AUC) | 217 | **.150** | **2.190** | **.030*** | **.178** | - | - | - | - | - |
| *Overall Model Diagnostics* | *217* | - | - | *.094* | - | *.033* | *.029* | *.094* | *20.108* | *23.172* |
| **Combined ERPs (AUC) - ERP + Age + Sex + PTA4** | | | | | | | | | | |
| omission *xxxxY* - standard *xxxxY* (Positive Cluster) (AUC) | 217 | -.070 | -1.018 | .310 | .565 | - | - | - | - | - |
| omission *xxxxY* - standard *xxxxY* (Negative Cluster) (AUC) | 217 | -.011 | -.162 | .871 | .971 | - | - | - | - | - |
| *xxxxx* deviant - standard (Negative Cluster) (AUC) | 217 | .123 | 1.782 | .076 | .565 | - | - | - | - | - |
| *Overall Model Diagnostics* | *217* | - | - | *.117* | - | *.074* | *.027* | *.117* | *19.916* | *23.239* |
| **Combined ERPs (AUC) - ERP + Age + Sex + PTA4 + THI** | | | | | | | | | | |

| Predictor | N | Standardised β | t-value | p-value | p_fdr | r coefficient | Δ $R^2$ | p (Δ $R^2$) | MAE | RMSE |
|---|---|---|---|---|---|---|---|---|---|---|
| omission *xxxxY* - standard *xxxxY* (Positive Cluster) (AUC) | 217 | -.040 | -.655 | .513 | .603 | - | - | - | - | - |
| omission *xxxxY* - standard *xxxxY* (Negative Cluster) (AUC) | 217 | -.067 | -1.113 | .267 | .534 | - | - | - | - | - |
| *xxxxx* deviant - standard (Negative Cluster) (AUC) | 217 | **.141** | **2.319** | **.021*** | **.107** | - | - | - | - | - |
| *Overall Model Diagnostics* | *217* | ***-*** | ***-*** | ***<.001**** | ***-*** | *.445* | *.213* | ***<.001**** | *17.038* | *20.636* |

*Note. N = participant count inside the given model step. Standardised β = Standardised regression coefficient. r coefficient = 10-fold cross-validated correlation coefficient r evaluating internal model shrinkage; MAE and RMSE are computed from held-out folds. Δ $R^2$ = change in variance explained at the given hierarchical step. p (Δ $R^2$) = statistical significance of the change in $R^2$ calculated via hierarchical F-change testing. For Behavioural & Demographic Variables, unadjusted univariable statistics are computed independently. For ERP Only, Δ $R^2$ represents the unique variance explained by the ERP alone. For subsequent hierarchical blocks, Δ $R^2$ represents the incremental variance explained by the stepwise addition of demographic, hearing-level, and clinical scale covariates.*

** p < .05. Bold fields indicate predictors that reached statistical significance.*

**Table 10**

**Sensitivity Analysis Table - Subjective Loudness (VAS)**

**Multi-Site Sensitivity Analysis: Parameter Stability Across Leave-One-Site-Out Folds**

| Predictor | Standardised β | SE | Unstandardised B [95% CI] | t-value | p-value |
|---|---|---|---|---|---|
| **Excluded Site: Dublin (Remaining N = 179)** | | | | | |
| ***xxxxx* deviant - standard (Negative Cluster) AUC** | **.151** | **78.401** | **180.752 [26.006, 335.498]** | **2.31** | **.022*** |
| Age (Years) | .090 | .128 | 0.144 [-0.109, 0.396] | 1.12 | .264 |
| Sex (Female vs. Male) | .014 | 3.051 | 0.667 [-5.354, 6.689] | .22 | .827 |
| Hearing Thresholds (PTA4) | .128 | .176 | 0.283 [-0.064, 0.630] | 1.61 | .109 |
| **Tinnitus Severity (THI)** | **.463** | **.086** | **0.616 [0.445, 0.786]** | **7.12** | **< .001*** |
| *Overall Model Diagnostics: Model-Fit r = .520, MAE = 16.19, RMSE = 19.73, Internal 10-Fold CV r = .492 [0.036, 0.818]* | | | | | |
| **Excluded Site: Ghent (Remaining N = 183)** | | | | | |
| ***xxxxx* deviant - standard (Negative Cluster) AUC** | **.132** | **81.742** | **163.088 [1.774, 324.403]** | **2.00** | **.048*** |
| Age (Years) | .084 | .127 | 0.129 [-0.121, 0.380] | 1.02 | .310 |
| Sex (Female vs. Male) | .025 | 2.998 | 1.128 [-4.788, 7.044] | .38 | .707 |
| Hearing Thresholds (PTA4) | .104 | .168 | 0.210 [-0.121, 0.542] | 1.25 | .212 |
| **Tinnitus Severity (THI)** | **.446** | **.095** | **0.643 [0.455, 0.831]** | **6.75** | **< .001*** |
| *Overall Model Diagnostics: Model-Fit r = .496, MAE = 16.61, RMSE = 19.76, Internal 10-Fold CV r = .266 [-0.335, 0.764]* | | | | | |

| Predictor | Standardised β | SE | Unstandardised B [95% CI] | t-value | p-value |
|---|---|---|---|---|---|
| **Excluded Site: Illinois (Remaining N = 190)** | | | | | |
| ***xxxxx* deviant - standard (Negative Cluster) AUC** | **.181** | **74.947** | **211.816 [63.950, 359.682]** | **2.83** | **.005*** |
| Age (Years) | .122 | .124 | 0.194 [-0.050, 0.439] | 1.57 | .119 |
| Sex (Female vs. Male) | .062 | 2.920 | 2.805 [-2.957, 8.567] | .96 | .338 |
| Hearing Thresholds (PTA4) | .106 | .161 | 0.222 [-0.095, 0.539] | 1.38 | .169 |
| **Tinnitus Severity (THI)** | **.424** | **.084** | **0.567 [0.401, 0.734]** | **6.71** | **< .001*** |
| *Overall Model Diagnostics: Model-Fit r = .517, MAE = 16.12, RMSE = 19.42, Internal 10-Fold CV r = .351 [-0.004, 0.687]* | | | | | |
| **Excluded Site: Regensburg (Remaining N = 178)** | | | | | |
| ***xxxxx* deviant - standard (Negative Cluster) AUC** | **.135** | **80.699** | **163.494 [4.207, 322.781]** | **2.03** | **.044*** |
| Age (Years) | .094 | .127 | 0.149 [-0.101, 0.399] | 1.18 | .241 |
| Sex (Female vs. Male) | .079 | 2.985 | 3.580 [-2.311, 9.472] | 1.20 | .232 |
| Hearing Thresholds (PTA4) | .033 | .184 | 0.077 [-0.286, 0.440] | .42 | .677 |
| **Tinnitus Severity (THI)** | **.488** | **.086** | **0.642 [0.472, 0.812]** | **7.46** | **< .001*** |
| *Overall Model Diagnostics: Model-Fit r = .519, MAE = 16.00, RMSE = 19.38, Internal 10-Fold CV r = .333 [-0.282, 0.723]* | | | | | |
| **Excluded Site: Tuebingen (Remaining N = 177)** | | | | | |
| ***xxxxx* deviant - standard (Negative Cluster) AUC** | **.190** | **82.412** | **233.026 [70.350, 395.703]** | **2.83** | **.005*** |

| Predictor | Standardised β | SE | Unstandardised B [95% CI] | t-value | p-value |
|---|---|---|---|---|---|
| Age (Years) | .108 | .137 | 0.181 [-0.090, 0.451] | 1.32 | .189 |
| Sex (Female vs. Male) | .013 | 3.103 | 0.599 [-5.527, 6.725] | .19 | .847 |
| Hearing Thresholds (PTA4) | .084 | .170 | 0.175 [-0.160, 0.510] | 1.03 | .305 |
| **Tinnitus Severity (THI)** | **.463** | **.088** | **0.623 [0.449, 0.797]** | **7.05** | **< .001*** |
| *Overall Model Diagnostics: Model-Fit r = .520, MAE = 16.52, RMSE = 19.88, Internal 10-Fold CV r = .101 [-0.423, 0.515]* | | | | | |
| **Excluded Site: Zuerich (Remaining N = 178)** | | | | | |
| *xxxxx* deviant - standard (Negative Cluster) AUC | .070 | 83.786 | 89.177 [-76.204, 254.558] | 1.06 | .289 |
| Age (Years) | .002 | .130 | 0.004 [-0.253, 0.260] | .03 | .978 |
| Sex (Female vs. Male) | .034 | 3.028 | 1.596 [-4.381, 7.573] | .53 | .599 |
| **Hearing Thresholds (PTA4)** | **.200** | **.173** | **0.432 [0.092, 0.773]** | **2.50** | **.013*** |
| **Tinnitus Severity (THI)** | **.481** | **.090** | **0.664 [0.487, 0.841]** | **7.40** | **< .001*** |
| *Overall Model Diagnostics: Model-Fit r = .528, MAE = 16.26, RMSE = 19.66, Internal 10-Fold CV r = .109 [-0.481, 0.532]* | | | | | |

*Note. Excluded Site indicates clinical centre omitted from the subcohort to test multi-centre parameter robustness. Standardised β and Unstandardised B [95% CI] are derived from linear regression models fitted to the remaining training cohort (N). Model-Fit r, MAE, and RMSE quantify in-sample explanatory fit; Internal 10-Fold CV r [95% CI] represents cross-validated shrinkage within the remaining cohort.*

**Table 11**

**Linear Regression Results Characterising Clinical Prediction - PTA4 Models**

| Predictor | N | Standardised β | t-value | p-value | p_fdr | r coefficient | Δ $R^2$ | p (Δ $R^2$) | MAE | RMSE |
|---|---|---|---|---|---|---|---|---|---|---|
| **Behavioural & Demographic Variables** | | | | | | | | | | |
| Age (Years) | 226 | **.579** | **10.618** | **< .001*** | **-** | .570 | .335 | **< .001*** | 6.684 | 8.920 |
| Sex (Female vs. Male) | 225 | -.106 | -1.594 | .112 | - | .049 | .011 | .112 | 8.490 | 10.885 |
| Tinnitus Distress (THI) | 226 | -.013 | -.192 | .848 | - | -.221 | .000 | .848 | 8.510 | 10.941 |
| Tinnitus Loudness (VAS) | 218 | **.170** | **2.535** | **.012*** | **-** | .108 | .029 | **.012*** | 8.381 | 10.706 |
| **ERP Only** | | | | | | | | | | |
| omission *xxxxY* - standard *xxxxY* (Positive Cluster) (PTP) | 217 | -.030 | -.434 | .665 | .848 | -.234 | .001 | .665 | 8.428 | 10.876 |
| omission *xxxxY* - standard *xxxxY* (Positive Cluster) (AUC) | 217 | -.019 | -.281 | .779 | .848 | -.261 | .000 | .779 | 8.410 | 10.873 |
| omission *xxxxY* - standard *xxxxY* (Negative Cluster) (PTP) | 217 | -.103 | -1.519 | .130 | .479 | -.131 | .011 | .130 | 8.379 | 10.864 |
| omission *xxxxY* - standard *xxxxY* (Negative Cluster) (AUC) | 217 | .087 | 1.281 | .201 | .479 | -.080 | .008 | .201 | 8.409 | 10.838 |
| *xxxxx* deviant - standard (Negative Cluster) (PTP) | 217 | -.085 | -1.255 | .211 | .479 | -.274 | .007 | .211 | 8.401 | 10.886 |
| *xxxxx* deviant - standard (Negative Cluster) (AUC) | 217 | **.150** | **2.226** | **.027*** | **.206** | .077 | .023 | **.027*** | 8.361 | 10.769 |
| **ERP + Age + Sex** | | | | | | | | | | |
| omission *xxxxY* - standard *xxxxY* (Positive Cluster) (PTP) | 217 | **-.114** | **-2.037** | **.043*** | **.172** | .566 | .348 | **< .001*** | 6.611 | 8.891 |
| omission *xxxxY* - standard *xxxxY* (Positive Cluster) (AUC) | 217 | -.024 | -.431 | .667 | .908 | .554 | .336 | **< .001*** | 6.690 | 8.969 |
| omission *xxxxY* - standard *xxxxY* (Negative Cluster) (PTP) | 217 | -.024 | -.426 | .671 | .908 | .550 | .326 | **< .001*** | 6.703 | 8.994 |

| Predictor | N | Standardised β | t-value | p-value | p_fdr | r coefficient | Δ R² | p (Δ R²) | MAE | RMSE |
|---|---|---|---|---|---|---|---|---|---|---|
| omission *xxxxY* - standard *xxxxY* (Negative Cluster) (AUC) | 217 | .031 | .544 | .587 | .908 | .555 | .329 | **< .001*** | 6.689 | 8.957 |
| *xxxxx* deviant - standard (Negative Cluster) (PTP) | 217 | .016 | .280 | .780 | .908 | .552 | .329 | **< .001*** | 6.725 | 8.984 |
| *xxxxx* deviant - standard (Negative Cluster) (AUC) | 217 | .065 | 1.162 | .247 | .754 | .560 | .317 | **< .001*** | 6.639 | 8.924 |
| **ERP + Age + Sex + THI + VAS** | | | | | | | | | | |
| omission *xxxxY* - standard *xxxxY* (Positive Cluster) (PTP) | 217 | -.109 | -1.941 | .054 | .268 | .557 | .008 | .280 | 6.682 | 8.964 |
| omission *xxxxY* - standard *xxxxY* (Positive Cluster) (AUC) | 217 | -.019 | -.329 | .742 | .889 | .544 | .009 | .246 | 6.782 | 9.051 |
| omission *xxxxY* - standard *xxxxY* (Negative Cluster) (PTP) | 217 | -.020 | -.359 | .720 | .889 | .542 | .009 | .243 | 6.793 | 9.064 |
| omission *xxxxY* - standard *xxxxY* (Negative Cluster) (AUC) | 217 | .038 | .668 | .505 | .841 | .548 | .009 | .219 | 6.756 | 9.022 |
| *xxxxx* deviant - standard (Negative Cluster) (PTP) | 217 | .010 | .171 | .865 | .889 | .544 | .009 | .243 | 6.789 | 9.045 |
| *xxxxx* deviant - standard (Negative Cluster) (AUC) | 217 | .050 | .883 | .378 | .841 | .549 | .007 | .313 | 6.722 | 9.007 |
| **Combined Multivariate ERPs (PTP) - ERP Only** | | | | | | | | | | |
| omission *xxxxY* - standard *xxxxY* (Positive Cluster) (PTP) | 217 | .020 | .262 | .794 | .848 | - | - | - | - | - |
| omission *xxxxY* - standard *xxxxY* (Negative Cluster) (PTP) | 217 | -.088 | -1.084 | .280 | .479 | - | - | - | - | - |
| *xxxxx* deviant - standard (Negative Cluster) (PTP) | 217 | -.050 | -.654 | .514 | .771 | - | - | - | - | - |
| *Overall Model Diagnostics* | *217* | - | - | *.433* | - | *-.170* | *.013* | *.433* | *8.427* | *10.932* |
| **Combined Multivariate ERPs (PTP) - ERP + Age + Sex** | | | | | | | | | | |
| omission *xxxxY* - standard *xxxxY* (Positive Cluster) (PTP) | 217 | **-.135** | **-2.147** | **.033*** | **.172** | - | - | - | - | - |
| omission *xxxxY* - standard *xxxxY* (Negative Cluster) (PTP) | 217 | .011 | .159 | .874 | .908 | - | - | - | - | - |

| Predictor | N | Standardised β | t-value | p-value | p_fdr | r coefficient | Δ R² | p (Δ R²) | MAE | RMSE |
|---|---|---|---|---|---|---|---|---|---|---|
| *xxxxx* deviant - standard (Negative Cluster) (PTP) | 217 | .054 | .836 | .404 | .908 | - | - | - | - | - |
| *Overall Model Diagnostics* | *217* | **-** | **-** | ***<.001**** | **-** | *.559* | *.339* | ***<.001**** | *6.690* | *8.943* |
| **Combined Multivariate ERPs (PTP) - ERP + Age + Sex + THI + VAS** | | | | | | | | | | |
| omission *xxxxY* - standard *xxxxY* (Positive Cluster) (PTP) | 217 | **-.129** | **-2.053** | **.041*** | **.268** | - | - | - | - | - |
| omission *xxxxY* - standard *xxxxY* (Negative Cluster) (PTP) | 217 | .016 | .245 | .806 | .889 | - | - | - | - | - |
| *xxxxx* deviant - standard (Negative Cluster) (PTP) | 217 | .044 | .680 | .497 | .841 | - | - | - | - | - |
| *Overall Model Diagnostics* | *217* | - | - | *.312* | - | *.551* | *.007* | *.312* | *6.769* | *9.010* |
| **Combined Multivariate ERPs (AUC) - ERP Only** | | | | | | | | | | |
| omission *xxxxY* - standard *xxxxY* (Positive Cluster) (AUC) | 217 | .013 | .192 | .848 | .848 | - | - | - | - | - |
| omission *xxxxY* - standard *xxxxY* (Negative Cluster) (AUC) | 217 | .077 | 1.130 | .260 | .479 | - | - | - | - | - |
| *xxxxx* deviant - standard (Negative Cluster) (AUC) | 217 | **.146** | **2.131** | **.034*** | **.206** | - | - | - | - | - |
| *Overall Model Diagnostics* | *217* | - | - | *.105* | - | *.052* | *.028* | *.105* | *8.465* | *10.846* |
| **Combined Multivariate ERPs (AUC) - ERP + Age + Sex** | | | | | | | | | | |
| omission *xxxxY* - standard *xxxxY* (Positive Cluster) (AUC) | 217 | -.011 | -.198 | .843 | .908 | - | - | - | - | - |
| omission *xxxxY* - standard *xxxxY* (Negative Cluster) (AUC) | 217 | .026 | .450 | .653 | .908 | - | - | - | - | - |
| *xxxxx* deviant - standard (Negative Cluster) (AUC) | 217 | .062 | 1.077 | .283 | .754 | - | - | - | - | - |
| *Overall Model Diagnostics* | *217* | **-** | **-** | ***<.001**** | **-** | *.548* | *.312* | ***<.001**** | *6.729* | *9.017* |
| **Combined Multivariate ERPs (AUC) - ERP + Age + Sex + THI + VAS** | | | | | | | | | | |

| **Predictor** | **N** | **Standardised β** | **t-value** | **p-value** | **p_fdr** | **r coefficient** | **Δ R²** | **p (Δ R²)** | **MAE** | **RMSE** |
|---|---|---|---|---|---|---|---|---|---|---|
| omission *xxxxY* - standard *xxxxY* (Positive Cluster) (AUC) | 217 | -.008 | -.140 | .889 | .889 | - | - | - | - | - |
| omission *xxxxY* - standard *xxxxY* (Negative Cluster) (AUC) | 217 | .033 | .585 | .559 | .860 | - | - | - | - | - |
| *xxxxx* deviant - standard (Negative Cluster) (AUC) | 217 | .046 | .798 | .426 | .841 | - | - | - | - | - |
| *Overall Model Diagnostics* | *217* | - | - | *.298* | - | *.538* | *.008* | *.298* | *6.824* | *9.100* |

*Note. N = participant count inside the given model step. Standardised β = Standardised regression coefficient. r coefficient = 10-fold cross-validated correlation coefficient r evaluating internal model shrinkage; MAE and RMSE are computed from held-out folds. Δ R² = change in variance explained at the given hierarchical step. p (Δ R²) = statistical significance of the change in R² calculated via hierarchical F-change testing. For Behavioural & Demographic Variables, unadjusted univariable statistics are computed independently. For ERP Only, Δ R² represents the unique variance explained by the ERP alone. For subsequent hierarchical blocks, Δ R² represents the incremental variance explained by the stepwise addition of demographic, hearing-level, and clinical scale covariates.*

** p < .05. Bold fields indicate predictors that reached statistical significance.*

**Table 12**

**Sensitivity Analysis Table - Hearing Thresholds (PTA4)**

**Multi-Site Sensitivity Analysis: Parameter Stability Across Leave-One-Site-Out Folds**

| Predictor | Standardised β | SE | Unstandardised B [95% CI] | t-value | p-value |
|---|---|---|---|---|---|
| **Excluded Site: Dublin (Remaining N = 187)** | | | | | |
| *xxxxx* deviant - standard (Negative Cluster) AUC | .019 | 33.891 | 10.808 [-56.059, 77.674] | .32 | .750 |
| **Age (Years)** | **.570** | **.045** | **0.421 [0.331, 0.510]** | **9.29** | **< .001*** |
| Sex (Female vs. Male) | -.042 | 1.303 | -0.900 [-3.470, 1.671] | -.69 | .491 |
| *Overall Model Diagnostics: Model-Fit r = .581, MAE = 6.52, RMSE = 8.69, Internal 10-Fold CV r = .588 [0.367, 0.794]* | | | | | |
| **Excluded Site: Ghent (Remaining N = 190)** | | | | | |
| *xxxxx* deviant - standard (Negative Cluster) AUC | .093 | 36.229 | 57.759 [-13.713, 129.230] | 1.59 | .113 |
| **Age (Years)** | **.586** | **.045** | **0.452 [0.363, 0.541]** | **10.00** | **< .001*** |
| Sex (Female vs. Male) | -.056 | 1.326 | -1.263 [-3.879, 1.353] | -.95 | .342 |
| *Overall Model Diagnostics: Model-Fit r = .609, MAE = 6.81, RMSE = 8.99, Internal 10-Fold CV r = .783 [0.668, 0.880]* | | | | | |
| **Excluded Site: Illinois (Remaining N = 191)** | | | | | |
| *xxxxx* deviant - standard (Negative Cluster) AUC | .064 | 34.038 | 35.805 [-31.343, 102.953] | 1.05 | .294 |
| **Age (Years)** | **.557** | **.047** | **0.425 [0.332, 0.518]** | **9.03** | **< .001*** |
| Sex (Female vs. Male) | -.013 | 1.327 | -0.275 [-2.892, 2.343] | -.21 | .836 |

| Predictor | Standardised β | SE | Unstandardised B [95% CI] | t-value | p-value |
|---|---|---|---|---|---|
| *Overall Model Diagnostics: Model-Fit r = .572, MAE = 6.49, RMSE = 8.90, Internal 10-Fold CV r = .464 [0.065, 0.724]* | | | | | |
| **Excluded Site: Regensburg (Remaining N = 186)** | | | | | |
| *xxxxx* deviant - standard (Negative Cluster) AUC | .063 | 33.595 | 34.109 [-32.177, 100.396] | 1.02 | .311 |
| **Age (Years)** | **.546** | **.044** | **0.384 [0.297, 0.471]** | **8.72** | **< .001*** |
| Sex (Female vs. Male) | .032 | 1.230 | 0.634 [-1.793, 3.062] | .52 | .607 |
| *Overall Model Diagnostics: Model-Fit r = .559, MAE = 6.13, RMSE = 8.25, Internal 10-Fold CV r = .511 [0.088, 0.777]* | | | | | |
| **Excluded Site: Tuebingen (Remaining N = 185)** | | | | | |
| *xxxxx* deviant - standard (Negative Cluster) AUC | .101 | 36.404 | 61.074 [-10.757, 132.904] | 1.68 | .095 |
| **Age (Years)** | **.568** | **.050** | **0.464 [0.365, 0.562]** | **9.30** | **< .001*** |
| Sex (Female vs. Male) | -.030 | 1.370 | -0.691 [-3.395, 2.012] | -.50 | .614 |
| *Overall Model Diagnostics: Model-Fit r = .600, MAE = 6.88, RMSE = 9.05, Internal 10-Fold CV r = .732 [0.629, 0.868]* | | | | | |
| **Excluded Site: Zuerich (Remaining N = 186)** | | | | | |
| *xxxxx* deviant - standard (Negative Cluster) AUC | .091 | 36.401 | 54.653 [-17.170, 126.475] | 1.50 | .135 |
| **Age (Years)** | **.561** | **.046** | **0.430 [0.338, 0.521]** | **9.25** | **< .001*** |
| Sex (Female vs. Male) | -.056 | 1.310 | -1.216 [-3.801, 1.370] | -.93 | .355 |
| *Overall Model Diagnostics: Model-Fit r = .587, MAE = 6.71, RMSE = 8.76, Internal 10-Fold CV r = .723 [0.594, 0.863]* | | | | | |

*Note.* *Excluded Site indicates clinical centre omitted from the subcohort to test multi-centre parameter robustness. Standardised β and Unstandardised B [95% CI] are derived from linear regression models fitted to the remaining training cohort (N). Model-Fit r, MAE, and RMSE quantify in-sample explanatory fit; Internal 10-Fold CV r [95% CI] represents cross-validated shrinkage within the remaining cohort.*

**Table 13**

**Linear Regression Results Characterising Clinical Prediction - Loudness (HL) Models**

| Predictor | N | Standardised β | t-value | p-value | p_fdr | r coefficient | Δ $R^2$ | p (Δ $R^2$) | MAE | RMSE |
|---|---|---|---|---|---|---|---|---|---|---|
| **Behavioural & Demographic Variables** | | | | | | | | | | |
| Age (Years) | 179 | **.428** | **6.293** | **< .001*** | - | .402 | .183 | **< .001*** | 15.802 | 19.219 |
| Sex (Female vs. Male) | 178 | **-.159** | **-2.138** | **.034*** | - | .095 | .025 | **.034*** | 16.882 | 20.862 |
| Hearing Loss (PTA4) | 179 | **.484** | **7.365** | **< .001*** | - | .462 | .235 | **< .001*** | 15.222 | 18.610 |
| Tinnitus Distress (THI) | 179 | **.153** | **2.059** | **.041*** | - | .061 | .023 | **.041*** | 17.020 | 21.037 |
| Tinnitus Loudness (VAS) | 172 | **.297** | **4.048** | **< .001*** | - | .270 | .088 | **< .001*** | 16.095 | 20.095 |
| **ERP Only** | | | | | | | | | | |
| omission *xxxxY* - standard *xxxxY* (Positive Cluster) (PTP) | 171 | .027 | .350 | .727 | .727 | -.065 | .001 | .727 | 17.271 | 21.009 |
| omission *xxxxY* - standard *xxxxY* (Positive Cluster) (AUC) | 171 | -.030 | -.393 | .695 | .727 | -.216 | .001 | .695 | 17.298 | 21.012 |
| omission *xxxxY* - standard *xxxxY* (Negative Cluster) (PTP) | 171 | -.047 | -.613 | .541 | .727 | -.211 | .002 | .541 | 17.300 | 21.006 |
| omission *xxxxY* - standard *xxxxY* (Negative Cluster) (AUC) | 171 | -.052 | -.681 | .497 | .727 | -.007 | .003 | .497 | 17.354 | 20.999 |
| *xxxxx* deviant - standard (Negative Cluster) (PTP) | 171 | -.046 | -.605 | .546 | .727 | -.260 | .002 | .546 | 17.285 | 21.030 |
| *xxxxx* deviant - standard (Negative Cluster) (AUC) | 171 | .087 | 1.132 | .259 | .727 | -.091 | .008 | .259 | 17.221 | 20.985 |
| **ERP + Age + Sex + PTA4** | | | | | | | | | | |
| omission *xxxxY* - standard *xxxxY* (Positive Cluster) (PTP) | 171 | -.033 | -.478 | .633 | .929 | .471 | .267 | **< .001*** | 14.762 | 18.379 |
| omission *xxxxY* - standard *xxxxY* (Positive Cluster) (AUC) | 171 | -.010 | -.152 | .880 | .929 | .466 | .266 | **< .001*** | 14.801 | 18.423 |
| omission *xxxxY* - standard *xxxxY* (Negative Cluster) (PTP) | 171 | -.006 | -.089 | .929 | .929 | .473 | .265 | **< .001*** | 14.735 | 18.338 |

| Predictor | N | Standardised β | t-value | p-value | p_fdr | r coefficient | Δ R² | p (Δ R²) | MAE | RMSE |
|---|---|---|---|---|---|---|---|---|---|---|
| omission *xxxxY* - standard *xxxxY* (Negative Cluster) (AUC) | 171 | -.080 | -1.200 | .232 | .521 | .480 | .271 | **< .001*** | 14.664 | 18.267 |
| *xxxxx* deviant - standard (Negative Cluster) (PTP) | 171 | -.071 | -1.046 | .297 | .535 | .476 | .270 | **< .001*** | 14.731 | 18.308 |
| *xxxxx* deviant - standard (Negative Cluster) (AUC) | 171 | -.011 | -.169 | .866 | .929 | .464 | .260 | **< .001*** | 14.800 | 18.453 |
| **ERP + Age + Sex + PTA4 + THI + VAS** | | | | | | | | | | |
| omission *xxxxY* - standard *xxxxY* (Positive Cluster) (PTP) | 171 | -.030 | -.453 | .651 | .895 | .504 | .045 | **.005*** | 14.325 | 18.007 |
| omission *xxxxY* - standard *xxxxY* (Positive Cluster) (AUC) | 171 | -.001 | -.019 | .985 | .985 | .501 | .045 | **.005*** | 14.347 | 18.035 |
| omission *xxxxY* - standard *xxxxY* (Negative Cluster) (PTP) | 171 | -.001 | -.021 | .984 | .985 | .506 | .045 | **.005*** | 14.339 | 17.967 |
| omission *xxxxY* - standard *xxxxY* (Negative Cluster) (AUC) | 171 | -.084 | -1.261 | .209 | .404 | .515 | .046 | **.005*** | 14.260 | 17.854 |
| *xxxxx* deviant - standard (Negative Cluster) (PTP) | 171 | -.083 | -1.248 | .214 | .404 | .510 | .047 | **.004*** | 14.270 | 17.913 |
| *xxxxx* deviant - standard (Negative Cluster) (AUC) | 171 | -.026 | -.390 | .697 | .902 | .498 | .046 | **.005*** | 14.327 | 18.075 |
| **Combined Multivariate ERPs (PTP) - ERP Only** | | | | | | | | | | |
| omission *xxxxY* - standard *xxxxY* (Positive Cluster) (PTP) | 171 | .063 | .733 | .464 | .727 | - | - | - | - | - |
| omission *xxxxY* - standard *xxxxY* (Negative Cluster) (PTP) | 171 | -.053 | -.546 | .586 | .727 | - | - | - | - | - |
| *xxxxx* deviant - standard (Negative Cluster) (PTP) | 171 | -.040 | -.430 | .667 | .727 | - | - | - | - | - |
| *Overall Model Diagnostics* | *171* | - | - | *.798* | - | *-.122* | *.006* | *.798* | *17.390* | *21.164* |
| **Combined ERPs (PTP) - ERP + Age + Sex + PTA4** | | | | | | | | | | |
| omission *xxxxY* - standard *xxxxY* (Positive Cluster) (PTP) | 171 | -.022 | -.278 | .781 | .929 | - | - | - | - | - |
| omission *xxxxY* - standard *xxxxY* (Negative Cluster) (PTP) | 171 | .053 | .621 | .535 | .876 | - | - | - | - | - |

| Predictor | N | Standardised β | t-value | p-value | p_fdr | r coefficient | Δ R² | p (Δ R²) | MAE | RMSE |
|---|---|---|---|---|---|---|---|---|---|---|
| *xxxxx* deviant - standard (Negative Cluster) (PTP) | 171 | -.091 | -1.106 | .270 | .535 | - | - | - | - | - |
| *Overall Model Diagnostics* | *171* | - | - | ***< .001**** | - | *.468* | *.268* | ***< .001**** | *14.821* | *18.411* |
| **Combined ERPs (PTP) - ERP + Age + Sex + PTA4 + THI + VAS** | | | | | | | | | | |
| omission *xxxxY* - standard *xxxxY* (Positive Cluster) (PTP) | 171 | -.018 | -.229 | .819 | .948 | - | - | - | - | - |
| omission *xxxxY* - standard *xxxxY* (Negative Cluster) (PTP) | 171 | .067 | .811 | .419 | .695 | - | - | - | - | - |
| *xxxxx* deviant - standard (Negative Cluster) (PTP) | 171 | -.113 | -1.397 | .164 | .404 | - | - | - | - | - |
| *Overall Model Diagnostics* | *171* | - | - | ***.004**** | - | *.505* | *.048* | ***.004**** | *14.312* | *17.989* |
| **Combined Multivariate ERPs (AUC) - ERP Only** | | | | | | | | | | |
| omission *xxxxY* - standard *xxxxY* (Positive Cluster) (AUC) | 171 | -.030 | -.385 | .701 | .727 | - | - | - | - | - |
| omission *xxxxY* - standard *xxxxY* (Negative Cluster) (AUC) | 171 | -.064 | -.819 | .414 | .727 | - | - | - | - | - |
| *xxxxx* deviant - standard (Negative Cluster) (AUC) | 171 | .090 | 1.160 | .248 | .727 | - | - | - | - | - |
| *Overall Model Diagnostics* | *171* | - | - | *.567* | - | *-.048* | *.012* | *.567* | *17.470* | *21.211* |
| **Combined ERPs (AUC) - ERP + Age + Sex + PTA4** | | | | | | | | | | |
| omission *xxxxY* - standard *xxxxY* (Positive Cluster) (AUC) | 171 | -.019 | -.278 | .781 | .929 | - | - | - | - | - |
| omission *xxxxY* - standard *xxxxY* (Negative Cluster) (AUC) | 171 | -.081 | -1.204 | .230 | .521 | - | - | - | - | - |
| *xxxxx* deviant - standard (Negative Cluster) (AUC) | 171 | -.006 | -.092 | .927 | .929 | - | - | - | - | - |
| *Overall Model Diagnostics* | *171* | - | - | ***< .001**** | - | *.458* | *.262* | ***< .001**** | *14.832* | *18.553* |
| **Combined ERPs (AUC) - ERP + Age + Sex + PTA4 + THI + VAS** | | | | | | | | | | |

| **Predictor** | **N** | **Standardised β** | **t-value** | **p-value** | **p_fdr** | **r coefficient** | **Δ R²** | **p (Δ R²)** | **MAE** | **RMSE** |
|---|---|---|---|---|---|---|---|---|---|---|
| omission *xxxxY* - standard *xxxxY* (Positive Cluster) (AUC) | 171 | -.010 | -.157 | .875 | .963 | - | - | - | - | - |
| omission *xxxxY* - standard *xxxxY* (Negative Cluster) (AUC) | 171 | -.083 | -1.230 | .221 | .404 | - | - | - | - | - |
| *xxxxx* deviant - standard (Negative Cluster) (AUC) | 171 | -.019 | -.277 | .782 | .948 | - | - | - | - | - |
| *Overall Model Diagnostics* | *171* | ***-*** | ***-*** | ***.005**** | ***-*** | *.497* | *.046* | ***.005**** | *14.388* | *18.111* |

*Note. N = participant count inside the given model step. Standardised β = Standardised regression coefficient. r coefficient = 10-fold cross-validated correlation coefficient r evaluating internal model shrinkage; MAE and RMSE are computed from held-out folds. Δ $R^2$ = change in variance explained at the given hierarchical step. p (Δ $R^2$) = statistical significance of the change in $R^2$ calculated via hierarchical F-change testing. For Behavioural & Demographic Variables, unadjusted univariable statistics are computed independently. For ERP Only, Δ $R^2$ represents the unique variance explained by the ERP alone. For subsequent hierarchical blocks, Δ $R^2$ represents the incremental variance explained by the stepwise addition of demographic, hearing-level, and clinical scale covariates.*

** p < .05. Bold fields indicate predictors that reached statistical significance.*

**Table 14**

**Sensitivity Analysis Table - Tinnitus Loudness (dB HL)**

**Multi-Site Sensitivity Analysis: Parameter Stability Across Leave-One-Site-Out Folds**

| Predictor | Standardised β | SE | Unstandardised B [95% CI] | t-value | p-value |
|---|---|---|---|---|---|
| **Excluded Site: Dublin (Remaining N = 133)** | | | | | |
| *xxxxx* deviant - standard (Negative Cluster) AUC | .029 | 82.859 | 32.851 [-131.125, 196.827] | .40 | .692 |
| **Age (Years)** | **.189** | **.133** | **0.277 [0.013, 0.540]** | **2.08** | **.040*** |
| Sex (Female vs. Male) | -.045 | 3.114 | -1.884 [-8.047, 4.279] | -.60 | .546 |
| **Hearing Thresholds (PTA4)** | **.388** | **.190** | **0.807 [0.430, 1.183]** | **4.24** | **< .001*** |
| Tinnitus Severity (THI) | .064 | .103 | 0.077 [-0.126, 0.280] | .75 | .455 |
| Subjective Loudness (VAS) | .109 | .078 | 0.100 [-0.056, 0.255] | 1.27 | .207 |
| *Overall Model Diagnostics: Model-Fit r = .572, MAE = 13.90, RMSE = 17.28, Internal 10-Fold CV r = .563 [-0.042, 0.856]* | | | | | |
| **Excluded Site: Ghent (Remaining N = 138)** | | | | | |
| *xxxxx* deviant - standard (Negative Cluster) AUC | -.059 | 85.890 | -68.584 [-238.494, 101.327] | -.80 | .426 |
| **Age (Years)** | **.192** | **.128** | **0.267 [0.014, 0.521]** | **2.09** | **.039*** |
| Sex (Female vs. Male) | -.071 | 2.990 | -2.879 [-8.794, 3.036] | -.96 | .337 |
| **Hearing Thresholds (PTA4)** | **.330** | **.179** | **0.621 [0.267, 0.976]** | **3.46** | **< .001*** |

| Predictor | Standardised β | SE | Unstandardised B [95% CI] | t-value | p-value |
|---|---|---|---|---|---|
| Tinnitus Severity (THI) | .098 | .111 | 0.130 [-0.089, 0.349] | 1.18 | .242 |
| Subjective Loudness (VAS) | .138 | .076 | 0.125 [-0.026, 0.275] | 1.64 | .104 |
| *Overall Model Diagnostics: Model-Fit r = .550, MAE = 13.56, RMSE = 16.97, Internal 10-Fold CV r = .700 [0.436, 0.901]* | | | | | |
| **Excluded Site: Illinois (Remaining N = 144)** | | | | | |
| *xxxxx* deviant - standard (Negative Cluster) AUC | -.066 | 74.437 | -72.802 [-219.996, 74.393] | -.98 | .330 |
| **Age (Years)** | **.255** | **.121** | **0.381 [0.142, 0.620]** | **3.15** | **.002*** |
| **Sex (Female vs. Male)** | **-.175** | **2.792** | **-7.309 [-12.830, -1.788]** | **-2.62** | **.010*** |
| **Hearing Thresholds (PTA4)** | **.323** | **.158** | **0.630 [0.317, 0.944]** | **3.98** | **< .001*** |
| Tinnitus Severity (THI) | .052 | .093 | 0.064 [-0.119, 0.247] | .69 | .490 |
| **Subjective Loudness (VAS)** | **.203** | **.071** | **0.187 [0.047, 0.327]** | **2.63** | **.009*** |
| *Overall Model Diagnostics: Model-Fit r = .634, MAE = 12.80, RMSE = 16.05, Internal 10-Fold CV r = .578 [0.190, 0.841]* | | | | | |
| **Excluded Site: Regensburg (Remaining N = 133)** | | | | | |
| *xxxxx* deviant - standard (Negative Cluster) AUC | .030 | 89.373 | 33.522 [-143.344, 210.388] | .38 | .708 |
| Age (Years) | .182 | .134 | 0.264 [-0.002, 0.530] | 1.96 | .052 |
| Sex (Female vs. Male) | -.105 | 3.142 | -4.291 [-10.508, 1.926] | -1.37 | .174 |

| Predictor | Standardised β | SE | Unstandardised B [95% CI] | t-value | p-value |
|---|---|---|---|---|---|
| **Hearing Thresholds (PTA4)** | **.279** | **.197** | **0.589 [0.199, 0.979]** | **2.99** | **.003*** |
| Tinnitus Severity (THI) | -.034 | .108 | -0.040 [-0.253, 0.173] | -.37 | .712 |
| **Subjective Loudness (VAS)** | **.276** | **.080** | **0.246 [0.087, 0.405]** | **3.06** | **.003*** |
| *Overall Model Diagnostics: Model-Fit r = .522, MAE = 13.93, RMSE = 17.37, Internal 10-Fold CV r = .129 [-0.379, 0.618]* | | | | | |
| **Excluded Site: Tuebingen (Remaining N = 136)** | | | | | |
| *xxxxx* deviant - standard (Negative Cluster) AUC | -.053 | 90.167 | -62.114 [-240.510, 116.283] | -.69 | .492 |
| **Age (Years)** | **.262** | **.143** | **0.411 [0.128, 0.695]** | **2.87** | **.005*** |
| Sex (Female vs. Male) | -.080 | 3.201 | -3.477 [-9.811, 2.857] | -1.09 | .279 |
| **Hearing Thresholds (PTA4)** | **.233** | **.183** | **0.459 [0.096, 0.822]** | **2.50** | **.014*** |
| Tinnitus Severity (THI) | .012 | .107 | 0.015 [-0.196, 0.227] | .14 | .887 |
| **Subjective Loudness (VAS)** | **.254** | **.079** | **0.231 [0.075, 0.387]** | **2.93** | **.004*** |
| *Overall Model Diagnostics: Model-Fit r = .550, MAE = 13.93, RMSE = 17.80, Internal 10-Fold CV r = .539 [-0.069, 0.896]* | | | | | |
| **Excluded Site: Zuerich (Remaining N = 171)** | | | | | |
| *xxxxx* deviant - standard (Negative Cluster) AUC | -.026 | 75.053 | -29.308 [-177.504, 118.887] | -.39 | .697 |
| **Age (Years)** | **.219** | **.117** | **0.323 [0.091, 0.555]** | **2.75** | **.007*** |
| Sex (Female vs. Male) | -.096 | 2.716 | -4.019 [-9.382, 1.345] | -1.48 | .141 |

| Predictor | Standardised β | SE | Unstandardised B [95% CI] | t-value | p-value |
|---|---|---|---|---|---|
| **Hearing Thresholds (PTA4)** | **.310** | **.160** | **0.617 [0.300, 0.933]** | **3.85** | **< .001*** |
| Tinnitus Severity (THI) | .040 | .092 | 0.049 [-0.132, 0.231] | .54 | .593 |
| **Subjective Loudness (VAS)** | **.195** | **.069** | **0.177 [0.041, 0.313]** | **2.57** | **.011*** |
| *Overall Model Diagnostics: Model-Fit r = .560, MAE = 13.72, RMSE = 17.23, Internal 10-Fold CV r = .587 [0.044, 0.843]* | | | | | |

*Note. Excluded Site indicates clinical centre omitted from the subcohort to test multi-centre parameter robustness. Standardised β and Unstandardised B [95% CI] are derived from linear regression models fitted to the remaining training cohort (N). Model-Fit r, MAE, and RMSE quantify in-sample explanatory fit; Internal 10-Fold CV r [95% CI] represents cross-validated shrinkage within the remaining cohort.*
